## Supplementary material for "The Prevalence, Prevention, and Treatment of Cardiovascular Diseases in Twelve African Countries (2014-2019): An Analysis of the World Health Organisation STEPwise Approach to Chronic Disease Risk Factor Surveillance": Codes: 1_Create_dataset_for_analysis.R

```
################################################################################
# Author: Wingston Ng'ambi
# Affiliations: University of Geneva; Kamuzu University of Health Sciences
# Copyright: CC BY-NC-SA 3.0 IGO
################################################################################
rm(list = ls(all = TRUE)) #Remove all data in memory


# Set a variable for the dynamic author in RMD files
################################################################################
set_author <- "Wingston Ng'ambi"
# Set a variable for the dynamic title in RMD files 
set_title <- "STEPS CVD in Africa Data Book"
################################################################################

# INCLUSION OF URBAN/RURAL AND REGION
# Write into the global environment 
.u_r_reg <<- TRUE
# OR
# .GlobalEnv$.u_r_reg <- TRUE

# Check the object
.u_r_reg


################################################################################
# LOADING AND CLEANING DATA
################################################################################

# Identify project's working directory
library(here)

# Load other main packages
library(tidyverse)
library(foreign)
library(readxl)
library(purrr)
library(haven)
library(dplyr)
library(tidyverse)
library(labelled)
library(epikit)
library(sjPlot)
library(gtsummary)
library(writexl)
library(mice)
library(magrittr)
library(visdat)
library(caret)
library(caTools)
library(finalfit)
library(here)


setwd("C:\\Users\\LENOVO\\Documents\\GitHub\\Illicit_alcohol_use_SSA\\data")

################################################################################
# LOAD DATA 
################################################################################
#Load data for each country and select the needed variables
#1.Algeria
#dirData <- "Data/stata_files/"
alg2016 <- read_dta("stata_files/dza2016.dta")

 
## Recode CVD dataset  
table(alg2016$stratum) # check any other codes for rural and urban residence

#Extract required dataset
alg2016 <- with(alg2016, data.frame(country   = "Algeria",
                                reside    = if_else(stratum==1,1,0), #1=urban, 0=Rural
                                survyear  = year,
                                psu       = psu,
                                stratum   = stratum,
                                wstep1   = wstep1,
                                wstep2 = wstep2,
                                wstep3   = wstep3,                                
                                sex       = if_else(sex == "Women" , 0, # "FEMALE"
                                                    1), # "Male",
                                age       = age,
                                occupation = if_else(c8 == 1, 0, # "Government employee"
                                                               if_else(c8 == 2 , 1, # "Non-Govt employee"
                                                                       if_else(c8 ==3 | c8 ==5 , 2, #Self-employed
                                                                               if_else(c8==4 | c8==8 | c8==9 , 3, # "Non-paid"
                                                                                       if_else(c8==6 , 3, # "Retired"
                                                                                       NA))))),
                                edlev =  if_else(c5 == 1, 0, # "None"
                                                   if_else(c5 == 2 | c5 == 3, 1, # "Primary"
                                                           if_else(c5==5 | c5 ==4 , 2, #"Secondary"
                                                                   if_else( c5==8 |c5==6 | c5==7, 3, # "Tertiary"
                                                                                   NA)))),
                                mstatus =if_else(c7 == 1, 0, # "Never married"
                                                 if_else(c7 == 2 | c7 == 6, 1, # "Currently married"
                                                         if_else(c7 == 3 | c7 == 4 | c5 ==5 , 2, #"Formerly married"
                                                                 NA))), 
                                edyears =c4,
                                #agegroup  = agerange, 
                                h12=h12,
                                h13a=h13a,
                                h13b=h13b,
                                b8=b8,
                                b9=b9,
                                d1=d1,
                                m11=m11,
                                m12=m12,
                                h1=h1,
                                d11a=d11a, 
                                d11b=d11b, 
                                d11c=d11c, 
                                d11d=d11d, 
                                d11e=d11e, 
                                d11f=d11f,
                                h6=h6, 
                                h7a=h7a, 
                                m4a=m4a, 
                                m5a=m5a, 
                                m6a=m6a, 
                                t1=t1, 
                                t8=t8, 
                                h3=h3, 
                                h8=h8, 
                                h9=h9, 
                                h14=h14, 
                                h20a=h20a, 
                                h20b=h20b, 
                                h20c=h20c, 
                                h20d=h20d, 
                                h20e=h20e, 
                                h20f=h20f, 
                                b1=b1, 
                                b6=b6, 
                                b8=b8,
                                smoke=t1,
                                a4 =a4,
                                a6= a6,
                                d5=d5,
                                d2=d2,
                                h2a=h2a,
                                b5=b5,
                                d3 = d3,
                                d4=d4,
                                p1=p1,
                                p2=p2,
                                p3a=p3a,
                                p3b=p3b,
                                p4=p4,
                                p5=p5,
                                p6a=p6a,
                                p6b=p6b,
                                p7=p7,
                                p8=p8,
                                p9a=p9a,
                                p9b=p9b,
                                p10=p10,
                                p11=p11,
                                p12a=p12a,
                                p12b=p12b,
                                p13=p13,
                                p14=p14,
                                p15a=p15a,
                                p15b=p15b,
                                p16a=p16a,
                                p16b=p16b,
                                code= code,
                                h18 = h18,
                                h19 = h19, h20=h20,
                                h17  = h17)
                )
Algeria2016 <- writexl::write_xlsx(alg2016,"alg2016.xlsx",col_names = TRUE, format_headers = TRUE,)
alg2016 <-read_excel("alg2016.xlsx")
#2. Benin
ben2015 <- read_dta("stata_files/ben2015.dta")


## Recode CVD dataset
table(ben2015$stratum) # check any other codes for rural and urban residence

#Extract required dataset
ben2015 <- with(ben2015, data.frame(country   = "Benin",
                                    reside    = if_else(stratum==1,1,0), #1=urban, 0=Rural,
                                    survyear  = year,
                                    psu       = psu,
                                    stratum   = stratum,
                                    wstep1   = wstep1,
                                    wstep2 = wstep2,
                                    wstep3   = wstep3,                                
                                    sex       = if_else(sex == "Women" , 0, # "FEMALE"
                                                        1), # "Male"
                                    age       = age,
                                    occupation =  if_else(c8 == 1, 0, # "Government employee"
                                                                      if_else(c8 == 2 , 1, # "Non-Govt employee"
                                                                              if_else(c8 ==3 | c8 ==5 , 2, #Self-employed
                                                                                      if_else(c8==4 | c8==8 | c8==9 , 3, # "Non-paid"
                                                                                              if_else(c8==6 , 3, # "Retired"
                                                                                                      NA))))),
                                    edlev = if_else(c5 == 88 | c5 == 2 | c5 == 1, 0, # "None"
                                                    if_else( c5 == 3, 1, # "Primary"
                                                            if_else(c5 ==4 | c5 == 5, 2, #"Secondary"
                                                                    if_else(c5==6 | c5==7, 3, # "Tertiary"
                                                                            NA)))),
                                    mstatus =if_else(c7 == 1, 0, # "Never married"
                                                     if_else(c7 == 2 | c7 == 6, 1, # "Currently married"
                                                             if_else(c7 == 3 | c7 == 4 | c5 ==5 , 2, #"Formerly married"
                                                                     NA))), 
                                    edyears =c4, 
                                    #agegroup  = agerange,
                                    h12=h12,
                                    h13a=h13a,
                                    h13b=h13b,
                                    b8=b8,
                                    b9=b9,
                                    d1=d1,
                                    m11=m11,
                                    m12=m12,
                                    h1=h1,
                                    d11a=d11a, 
                                    d11b=d11b, 
                                    d11c=d11c, 
                                    d11d=d11d, 
                                    d11e=d11e, 
                                    d11f=d11f,
                                    h6=h6, 
                                    h7a=h7a, 
                                    m4a=m4a, 
                                    m5a=m5a, 
                                    m6a=m6a, 
                                    t1=t1, 
                                    t8=t8, 
                                    h3=h3, 
                                    h8=h8, 
                                    h9=h9, 
                                    h14=h14, 
                                    h20a=h20a, 
                                    h20b=h20b, 
                                    h20c=h20c, 
                                    h20d=h20d, 
                                    h20e=h20e, 
                                    h20f=h20f, 
                                    b1=b1, 
                                    b6=b6, 
                                    b8=b8,
                                    smoke=t1,
                                    a4 =a4,
                                    a6= a6,
                                    d5=d5,
                                    d2=d2,
                                    h2a=h2a,
                                    b5=b5,
                                    d3 = d3,
                                    d4=d4,
                                    p1=p1,
                                    p2=p2,
                                    p3a=p3a,
                                    p3b=p3b,
                                    p4=p4,
                                    p5=p5,
                                    p6a=p6a,
                                    p6b=p6b,
                                    p7=p7,
                                    p8=p8,
                                    p9a=p9a,
                                    p9b=p9b,
                                    p10=p10,
                                    p11=p11,
                                    p12a=p12a,
                                    p12b=p12b,
                                    p13=p13,
                                    p14=p14,
                                    p15a=p15a,
                                    p15b=p15b,
                                    p16a=p16a,
                                    p16b=p16b,
                                    code= code,
                                    h18 = h18,
                                    h19 = h19, h20=h20,
                                    h17  = h17)
)

Benin2015 <- writexl::write_xlsx(ben2015,"C:/Users/LENOVO/Documents/HappyGitwithR/steps_ssa_data/CVD_Paper_2023/stepsKGZ2023/Data/ben2015.xlsx",col_names = TRUE, format_headers = TRUE,)
ben2015 <-read_excel("ben2015.xlsx")

#3.Botswana
bwa2014 <- read_dta("stata_files/bwa2014.dta")

nrows=nrow(bwa2014)

## Recode CVD dataset
table(bwa2014$stratum) # check any other codes for rural and urban residence

#bwa2014$stratum <-binom::binom.sim(nrows, nrows,p=0.51) #1436+1401 (urban (0.51) =1436/(1436+1401))
#Extract required dataset
bwa2014 <- with(bwa2014, data.frame(country   = "Botswana",
                                    reside    = sample(x = 0:1, size = nrows, replace = TRUE, prob = c(0.49,0.51)), #1=urban, 0=Rural: #Simulated residence in Botswana based on distribution of rural and urban clusters from Botswana STEPS report
                                    survyear  = year,
                                    psu       = psu,
                                    stratum   = stratum,
                                    wstep1   = wstep1,
                                    wstep2 = wstep2,
                                    wstep3   = wstep3,                                
                                    sex       = if_else(sex == "Women" , 0, # "FEMALE"
                                                        1), # "Male",
                                    age       = age,
                                    occupation = if_else(c8 == 1, 0, # "Government employee"
                                                                      if_else(c8 == 2 , 1, # "Non-Govt employee"
                                                                              if_else(c8 ==3 | c8 ==5 , 2, #Self-employed
                                                                                      if_else(c8==4 | c8==8 | c8==9 , 3, # "Non-paid"
                                                                                              if_else(c8==6 , 3, # "Retired"
                                                                                                      NA))))),
                                    edlev = if_else(c5 == 88 | c5 == 2 | c5 == 1, 0, # "None"
                                                    if_else( c5 == 3, 1, # "Primary"
                                                             if_else(c5==6 | c5 ==4 | c5 == 5, 2, #"Secondary"
                                                                     if_else( c5==9 | c5==8 | c5==7, 3, # "Tertiary"
                                                                             NA)))),
                                    mstatus =if_else(c7 == 1, 0, # "Never married"
                                                     if_else(c7 == 2 | c7 == 6, 1, # "Currently married"
                                                             if_else(c7 == 3 | c7 == 4 | c5 ==5 , 2, #"Formerly married"
                                                                     NA))), 
                                    edyears =c4,
                                    #agegroup  = agerange,
                                    h12=h12,
                                    h13a=h13a,
                                    h13b=h13b,
                                    b8=b8,
                                    b9=b9,
                                    d1=d1,
                                    m11=m11,
                                    m12=m12,
                                    h1=h1,
                                    # d11a=d11a, 
                                    # d11b=d11b, 
                                    # d11c=d11c, 
                                    # d11d=d11d, 
                                    # d11e=d11e, 
                                    # d11f=d11f,
                                    h6=h6, 
                                    h7a=h7a, 
                                    m4a=m4a, 
                                    m5a=m5a, 
                                    m6a=m6a, 
                                    t1=t1, 
                                    t8=t8, 
                                    h3=h3, 
                                    h8=h8, 
                                    h9=h9, 
                                    h14=h14, 
                                    h20a=h20a, 
                                    h20b=h20b, 
                                    h20c=h20c, 
                                    h20d=h20d, 
                                    h20e=h20e, 
                                    h20f=h20f, 
                                    b1=b1, 
                                    b6=b6, 
                                    b8=b8,
                                    smoke=t1,
                                    a4 =a4,
                                    a6= a6,
                                    d5=d5,
                                    d2=d2,
                                    h2a=h2a,
                                    b5=b5,
                                    d3 = d3,
                                    d4=d4,
                                    p1=p1,
                                    p2=p2,
                                    p3a=p3a,
                                    p3b=p3b,
                                    p4=p4,
                                    p5=p5,
                                    p6a=p6a,
                                    p6b=p6b,
                                    p7=p7,
                                    p8=p8,
                                    p9a=p9a,
                                    p9b=p9b,
                                    p10=p10,
                                    p11=p11,
                                    p12a=p12a,
                                    p12b=p12b,
                                    p13=p13,
                                    p14=p14,
                                    p15a=p15a,
                                    p15b=p15b,
                                    p16a=p16a,
                                    p16b=p16b,
                                    code= code,
                                    h18 = h18,
                                    h19 = h19, h20=h20,
                                    h17  = h17)
)

bwa2014$stratum <-bwa2014$reside+1 #Rework the data on stratum

Botswana2014 <- writexl::write_xlsx(bwa2014,"C:/Users/LENOVO/Documents/HappyGitwithR/steps_ssa_data/CVD_Paper_2023/stepsKGZ2023/Data/bwa2014.xlsx",col_names = TRUE, format_headers = TRUE,)
bwa2014 <-read_excel("bwa2014.xlsx")

#bwa2014[,2] <- sample(x = 0:1, size = nrows, replace = TRUE, prob = c(0.49,0.51)) #Simulated residence in Botswana based on distribution of rural and urban clusters from Botswana STEPS report

#4. Eswatini (simulate the residence like in Botswana: 4,865 total 1,286 Urban 3,579 Rural) from MICS 2014 Report
swz2014 <- read_dta("stata_files/swz2014.dta")


## Recode CVD dataset
table(swz2014$stratum) # check any other codes for rural and urban residence
nrows=nrow(swz2014)

#Extract required dataset
swz2014 <- with(swz2014, data.frame(country   = "Eswatini",
                                    reside    = sample(x = 0:1, size = nrows, replace = TRUE, prob = c(0.2643371,0.7356629)),
                                    survyear  = year,
                                    psu       = psu,
                                    stratum   = stratum,
                                    wstep1   = wstep1,
                                    wstep2 = wstep2,
                                    wstep3   = wstep3,                                
                                    sex       = if_else(sex == "Women" , 0, # "FEMALE"
                                                        1), # "Male"
                                    age       = age,
                                    occupation =  if_else(c8 == 1, 0, # "Government employee"
                                                                      if_else(c8 == 2 , 1, # "Non-Govt employee"
                                                                              if_else(c8 ==3 | c8 ==5 , 2, #Self-employed
                                                                                      if_else(c8==4 | c8==8 | c8==9 , 3, # "Non-paid"
                                                                                              if_else(c8==6 , 3, # "Retired"
                                                                                                      NA))))),
                                    edlev = if_else(c5 == 88 | c5 == 2 | c5 == 1, 0, # "None"
                                                    if_else( c5 == 3, 1, # "Primary"
                                                             if_else(c5 ==4 | c5 == 5, 2, #"Secondary"
                                                                     if_else( c5==6 | c5==7, 3, # "Tertiary"
                                                                              NA)))),  
                                    mstatus =if_else(c7 == 1, 0, # "Never married"
                                                     if_else(c7 == 2 | c7 == 6, 1, # "Currently married"
                                                             if_else(c7 == 3 | c7 == 4 | c5 ==5 , 2, #"Formerly married"
                                                                     NA))), 
                                    edyears =c4,
                                    #agegroup  = agerange,
                                    h12=h12,
                                    h13a=h13a,
                                    h13b=h13b,
                                    b8=b8,
                                    b9=b9,
                                    d1=d1,
                                    m11=m11,
                                    m12=m12,
                                    h1=h1,
                                    d11a=d11a, 
                                    d11b=d11b, 
                                    d11c=d11c, 
                                    d11d=d11d, 
                                    d11e=d11e, 
                                    d11f=d11f,
                                    h6=h6, 
                                    h7a=h7a, 
                                    m4a=m4a, 
                                    m5a=m5a, 
                                    m6a=m6a, 
                                    t1=t1, 
                                    t8=t8, 
                                    h3=h3, 
                                    h8=h8, 
                                    h9=h9, 
                                    h14=h14, 
                                    h20a=h20a, 
                                    h20b=h20b, 
                                    h20c=h20c, 
                                    h20d=h20d, 
                                    h20e=h20e, 
                                    h20f=h20f, 
                                    b1=b1, 
                                    b6=b6, 
                                    b8=b8,
                                    smoke=t1,
                                    a4 =a4,
                                    a6= a6,
                                    d5=d5,
                                    d2=d2,
                                    h2a=h2a,
                                    b5=b5,
                                    d3 = d3,
                                    d4=d4,
                                    p1=p1,
                                    p2=p2,
                                    p3a=p3a,
                                    p3b=p3b,
                                    p4=p4,
                                    p5=p5,
                                    p6a=p6a,
                                    p6b=p6b,
                                    p7=p7,
                                    p8=p8,
                                    p9a=p9a,
                                    p9b=p9b,
                                    p10=p10,
                                    p11=p11,
                                    p12a=p12a,
                                    p12b=p12b,
                                    p13=p13,
                                    p14=p14,
                                    p15a=p15a,
                                    p15b=p15b,
                                    p16a=p16a,
                                    p16b=p16b,
                                    code= code,
                                    h18 = h18,
                                    h19 = h19, h20=h20,
                                    h17  = h17)
)

#swz2014[,2] <- sample(x = 0:1, size = nrows, replace = TRUE, prob = c(0.49,0.51)) #Simulated residence in Botswana based on distribution of rural and urban clusters from Botswana STEPS report

swz2014$stratum <-swz2014$reside+1 #Rework the data on stratum
Eswatini <- writexl::write_xlsx(swz2014,"C:/Users/LENOVO/Documents/HappyGitwithR/steps_ssa_data/CVD_Paper_2023/stepsKGZ2023/Data/swz2014.xlsx",col_names = TRUE, format_headers = TRUE,)

swz2014 <-read_excel("swz2014.xlsx")


#5.Ethiopia
eth2015 <- read_dta("stata_files/eth2015.dta")

## Recode CVD dataset
table(eth2015$urbanrural) # check any other codes for rural and urban residence

#Extract required dataset
eth2015 <- with(eth2015, data.frame(country   = "Ethiopia",
                                    reside    = if_else(urbanrural==1,1,0),
                                    survyear  = year,
                                    psu       = psu,
                                    stratum   = stratum,
                                    wstep1   = wstep1,
                                    wstep2 = wstep2,
                                    wstep3   = wstep3,                                
                                    sex       = if_else(c1 == 2 , 0, # "FEMALE"
                                                           1), # "Male"
                                    age       = c3,
                                    occupation  = if_else(c8 == 1, 0, # "Government employee"
                                                                      if_else(c8 == 2 , 1, # "Non-Govt employee"
                                                                              if_else(c8 ==3 | c8 ==5 , 2, #Self-employed
                                                                                      if_else(c8==4 | c8==8 | c8==9 , 3, # "Non-paid"
                                                                                              if_else(c8==6 , 3, # "Retired"
                                                                                                      NA))))),
                                    edlev = if_else(c5 == 88 | c5 == 2 | c5 == 1, 0, # "None"
                                                    if_else( c5 == 3, 1, # "Primary"
                                                             if_else(c5 ==4, 2, #"Secondary"
                                                                     if_else( c5==6 | c5==5, 3, # "Tertiary"
                                                                              NA)))),  
                                    mstatus =if_else(c7 == 1, 0, # "Never married"
                                                     if_else(c7 == 2 | c7 == 6, 1, # "Currently married"
                                                             if_else(c7 == 3 | c7 == 4 | c5 ==5 , 2, #"Formerly married"
                                                                     NA))), 
                                    edyears =c4,
                                    #agegroup  = agecat,
                                    h12=h12,
                                    h13a=h13a,
                                    h13b=h13b,
                                    b8=b8,
                                    b9=b9,
                                    d1=d1,
                                    m11=m11,
                                    m12=m12,
                                    h1=h1,
                                    d11a=d11a, 
                                    d11b=d11b, 
                                    d11c=d11c, 
                                    d11d=d11d, 
                                    d11e=d11e, 
                                    d11f=d11f,
                                    h6=h6, 
                                    h7a=h7a, 
                                    m4a=m4a, 
                                    m5a=m5a, 
                                    m6a=m6a, 
                                    t1=t1, 
                                    t8=t8, 
                                    h3=h3, 
                                    h8=h8, 
                                    h9=h9, 
                                    h14=h14, 
                                    h20a=h20a, 
                                    h20b=h20b, 
                                    h20c=h20c, 
                                    h20d=h20d, 
                                    h20e=h20e, 
                                    h20f=h20f, 
                                    b1=b1, 
                                    b6=b6, 
                                    b8=b8, 
                                    smoke=t1,
                                    a4 =a4,
                                    a6= a6,
                                    d5=d5,
                                    d2=d2,
                                    h2a=h2a,
                                    b5=b5,
                                    d3 = d3,
                                    d4=d4,
                                    p1=p1,
                                    p2=p2,
                                    p3a=p3a,
                                    p3b=p3b,
                                    p4=p4,
                                    p5=p5,
                                    p6a=p6a,
                                    p6b=p6b,
                                    p7=p7,
                                    p8=p8,
                                    p9a=p9a,
                                    p9b=p9b,
                                    p10=p10,
                                    p11=p11,
                                    p12a=p12a,
                                    p12b=p12b,
                                    p13=p13,
                                    p14=p14,
                                    p15a=p15a,
                                    p15b=p15b,
                                    p16a=p16a,
                                    p16b=p16b,
                                    code= code,
                                    h18 = h18,
                                    h19 = h19, h20=h20,
                                    h17  = h17)
)

eth2015$stratum <-eth2015$reside+1 #Rework the data on stratum
Ethiopia2015 <- writexl::write_xlsx(eth2015,"C:/Users/LENOVO/Documents/HappyGitwithR/steps_ssa_data/CVD_Paper_2023/stepsKGZ2023/Data/eth2015.xlsx",col_names = TRUE, format_headers = TRUE,)

eth2015 <-read_excel("eth2015.xlsx")

#6. Kenya
ken2015 <- read_dta("stata_files/ken2015.dta")

## Recode CVD dataset
table(ken2015$urbrur) # check any other codes for rural and urban residence

#Extract required dataset
ken2015 <- with(ken2015, data.frame(country   = "Kenya",
                                    reside    = if_else(urbrur==1,1,0),
                                    survyear  = year,
                                    psu       = psu,
                                    stratum   = stratum,
                                    wstep1   = wstep1,
                                    wstep2 = wstep2,
                                    wstep3   = wstep3,                                
                                    sex       = if_else(sex == "Women" , 0, # "FEMALE"
                                                        1), # "Male",
                                    age       = age,
                                    occupation = if_else(c8 == 1, 0, # "Government employee"
                                                         if_else(c8 == 2 , 1, # "Non-Govt employee"
                                                                 if_else(c8 ==3 | c8 ==5 , 2, #Self-employed
                                                                         if_else(c8==4 | c8==8 | c8==9 , 3, # "Non-paid"
                                                                                 if_else(c8==6 , 3, # "Retired"
                                                                                         NA))))),
                                    edlev = if_else(c5 == "No formal schooling", 0, # "None"
                                                    if_else( c5 == "Primary school completed" | c5=="Primary school incomplete", 1, # "Primary"
                                                             if_else(c5 =="A-Level completed" | c5 == "Secondary school completed" | c5=="Secondary school incomplete", 2, #"Secondary"
                                                                     if_else( c5=="Post-graduate degree" | c5=="College/University completed", 3, # "Tertiary"
                                                                              NA)))),  
                                    mstatus =if_else(c7 == "Never married", 0, # "Never married"
                                                     if_else(c7 == "Cohabiting" | c7 == "Currently married", 1, # "Currently married"
                                                             if_else(c7 == "Widowed" | c7 == "Separated" | c5 =="Divorced" , 2, #"Formerly married"
                                                                     NA))), 
                                    edyears =c4, 
                                   # agegroup  = agerange,
                                   h12=h12,
                                   h13a=h13a,
                                   h13b=h13b,
                                   b8=b8,
                                   b9=b9,
                                   d1=d1,
                                   m11=m11,
                                   m12=m12,
                                   h1=h1,
                                   d11a=d11a, 
                                   d11b=d11b, 
                                   d11c=d11c, 
                                   d11d=d11d, 
                                   d11e=d11e, 
                                   d11f=d11f,
                                   h6=h6, 
                                   h7a=h7a, 
                                   m4a=m4a, 
                                   m5a=m5a, 
                                   m6a=m6a, 
                                   t1=t1, 
                                   t8=t8, 
                                   h3=h3, 
                                   h8=h8, 
                                   h9=h9, 
                                   h14=h14, 
                                   h20a=h20a, 
                                   h20b=h20b, 
                                   h20c=h20c, 
                                   h20d=h20d, 
                                   h20e=h20e, 
                                   h20f=h20f, 
                                   b1=b1, 
                                   b6=b6, 
                                   b8=b8,
                                    smoke=t1,
                                    a4 =a4,
                                    a6= a6,
                                    d5=d5,
                                    d2=d2,
                                    h2a=h2a,
                                    b5=b5,
                                    d3 = d3,
                                    d4=d4,
                                    p1=p1,
                                    p2=p2,
                                    p3a=p3a,
                                    p3b=p3b,
                                    p4=p4,
                                    p5=p5,
                                    p6a=p6a,
                                    p6b=p6b,
                                    p7=p7,
                                    p8=p8,
                                    p9a=p9a,
                                    p9b=p9b,
                                    p10=p10,
                                    p11=p11,
                                    p12a=p12a,
                                    p12b=p12b,
                                    p13=p13,
                                    p14=p14,
                                    p15a=p15a,
                                    p15b=p15b,
                                    p16a=p16a,
                                    p16b=p16b,
                                    code= code,
                                    h18 = h18,
                                    h19 = h19, h20=h20,
                                    h17  = h17)
)
Kenya2015 <- writexl::write_xlsx(ken2015,"C:/Users/LENOVO/Documents/HappyGitwithR/steps_ssa_data/CVD_Paper_2023/stepsKGZ2023/Data/ken2015.xlsx",col_names = TRUE, format_headers = TRUE,)
ken2015 <-read_excel("ken2015.xlsx")
 
#7. Malawi (check coding for rural and urban and also for the other variables)
mwi2017 <- read_dta("stata_files/mwi2017.dta")

## Recode CVD dataset
table(mwi2017$urbanrural) # check any other codes for rural and urban residence

#Extract required dataset
mwi2017 <- with(mwi2017, data.frame(country   = "Malawi",
                                    reside    = if_else(urbanrural=="Urban",1,0),
                                    survyear  = year,
                                    psu       = psu,
                                    stratum   = if_else(stratum == "North", 1, # "North"
                                                        if_else(stratum == "Central", 2, # "Centre"
                                                                if_else(stratum == "South" , 3, #"South"
                                                                        NA))),
                                    wstep1   = wstep1,
                                    wstep2 = wstep2,
                                    wstep3   = wstep3,                                
                                    sex       = if_else(sex == "Women" , 0, # "FEMALE"
                                                        1), # "Male"
                                    age       = age,
                                    occupation = if_else(c8 == 1, 0, # "Government employee"
                                                         if_else(c8 == 2 , 1, # "Non-Govt employee"
                                                                 if_else(c8 ==3 | c8 ==5 , 2, #Self-employed
                                                                         if_else(c8==4 | c8==8 | c8==9 , 3, # "Non-paid"
                                                                                 if_else(c8==6 , 3, # "Retired"
                                                                                         NA))))),
                                    edlev = if_else(c5 == 88 | c5 == 1, 0, # "None"
                                                    if_else( c5 == 3 | c5==2, 1, # "Primary"
                                                             if_else(c5 ==4, 2, #"Secondary"
                                                                     if_else( c5==5, 3, # "Tertiary"
                                                                              NA)))),  
                                    mstatus =if_else(c7 == 1, 0, # "Never married"
                                                     if_else(c7 == 2 | c7 == 6, 1, # "Currently married"
                                                             if_else(c7 == 3 | c7 == 4 | c5 ==5 , 2, #"Formerly married"
                                                                     NA))), 
                                    edyears =c4,
                                    #agegroup  = agerange,
                                    h12=h12,
                                    h13a=h13a,
                                    h13b=h13b,
                                    b8=b8,
                                    b9=b9,
                                    d1=d1,
                                    m11=m11,
                                    m12=m12,
                                    h1=h1,
                                    d11a=d11a, 
                                    d11b=d11b, 
                                    d11c=d11c, 
                                    d11d=d11d, 
                                    d11e=d11e, 
                                    d11f=d11f,
                                    h6=h6, 
                                    h7a=h7a, 
                                    m4a=m4a, 
                                    m5a=m5a, 
                                    m6a=m6a, 
                                    t1=t1, 
                                    t8=t8, 
                                    h3=h3, 
                                    h8=h8, 
                                    h9=h9, 
                                    h14=h14, 
                                    h20a=h20a, 
                                    h20b=h20b, 
                                    h20c=h20c, 
                                    h20d=h20d, 
                                    h20e=h20e, 
                                    h20f=h20f, 
                                    b1=b1, 
                                    b6=b6, 
                                    b8=b8,
                                    smoke=t1,
                                    a4 =a4,
                                    a6= a6,
                                    d5=d5,
                                    d2=d2,
                                    h2a=h2a,
                                    b5=b5,
                                    d3 = d3,
                                    d4=d4,
                                    p1=p1,
                                    p2=p2,
                                    p3a=p3a,
                                    p3b=p3b,
                                    p4=p4,
                                    p5=p5,
                                    p6a=p6a,
                                    p6b=p6b,
                                    p7=p7,
                                    p8=p8,
                                    p9a=p9a,
                                    p9b=p9b,
                                    p10=p10,
                                    p11=p11,
                                    p12a=p12a,
                                    p12b=p12b,
                                    p13=p13,
                                    p14=p14,
                                    p15a=p15a,
                                    p15b=p15b,
                                    p16a=p16a,
                                    p16b=p16b,
                                    code= code,
                                    h18 = h18,
                                    h19 = h19, h20=h20,
                                    h17  = h17)
)
Malawi2017 <- writexl::write_xlsx(mwi2017,"C:/Users/LENOVO/Documents/HappyGitwithR/steps_ssa_data/CVD_Paper_2023/stepsKGZ2023/Data/mwi2017.xlsx",col_names = TRUE, format_headers = TRUE,)
mwi2017 <-read_excel("mwi2017.xlsx")

#8. Morocco
mar2017 <- read_dta("stata_files/mar2017.dta")

## Recode CVD dataset
table(mar2017$stratum) # check any other codes for rural and urban residence

#Extract required dataset
mar2017 <- with(mar2017, data.frame(country   = "Morocco",
                                    reside    = if_else(stratum==2,1,0),
                                    survyear  = year,
                                    psu       = psu,
                                    stratum   = stratum,
                                    wstep1   = wstep1,
                                    wstep2 = wstep2,
                                    wstep3   = wstep3,                                
                                    sex       = if_else(sex == "Women" , 0, # "FEMALE"
                                                        1), # "Male"
                                    age       = age,
                                    occupation = if_else(c8 == 1, 0, # "Government employee"
                                                         if_else(c8 == 2 , 1, # "Non-Govt employee"
                                                                 if_else(c8 ==3 | c8 ==5 , 2, #Self-employed
                                                                         if_else(c8==4 | c8==8 | c8==9 , 3, # "Non-paid"
                                                                                 if_else(c8==6 , 3, # "Retired"
                                                                                         NA))))),
                                    edlev = if_else(c5 == 88 |  c5 == 1, 0, # "None"
                                                    if_else( c5 == 2, 1, # "Primary"
                                                             if_else(c5 ==3 | c5 == 4, 2, #"Secondary"
                                                                     if_else( c5==5, 3, # "Tertiary"
                                                                              NA)))), 
                                    mstatus =if_else(c7 == 1, 0, # "Never married"
                                                     if_else(c7 == 2 | c7 == 6, 1, # "Currently married"
                                                             if_else(c7 == 3 | c7 == 4 | c5 ==5 , 2, #"Formerly married"
                                                                     NA))), 
                                    edyears =c4,
                                    #agegroup  = agerange,
                                    h12=h12,
                                    h13a=h13a,
                                    h13b=h13b,
                                    b8=b8,
                                    b9=b9,
                                    d1=d1,
                                    m11=m11,
                                    m12=m12,
                                    h1=h1,
                                    d11a=d11a, 
                                    d11b=d11b, 
                                    d11c=d11c, 
                                    d11d=d11d, 
                                    d11e=d11e, 
                                    d11f=d11f,
                                    h6=h6, 
                                    h7a=h7a, 
                                    m4a=m4a, 
                                    m5a=m5a, 
                                    m6a=m6a, 
                                    t1=t1, 
                                    t8=t8, 
                                    h3=h3, 
                                    h8=h8, 
                                    h9=h9, 
                                    h14=h14, 
                                    h20a=h20a, 
                                    h20b=h20b, 
                                    h20c=h20c, 
                                    h20d=h20d, 
                                    h20e=h20e, 
                                    h20f=h20f, 
                                    b1=b1, 
                                    b6=b6, 
                                    b8=b8,
                                    smoke=t1,
                                    a4 =a4,
                                    a6= a6,
                                    d5=d5,
                                    d2=d2,
                                    h2a=h2a,
                                    b5=b5,
                                    d3 = d3,
                                    d4=d4,
                                    p1=p1,
                                    p2=p2,
                                    p3a=p3a,
                                    p3b=p3b,
                                    p4=p4,
                                    p5=p5,
                                    p6a=p6a,
                                    p6b=p6b,
                                    p7=p7,
                                    p8=p8,
                                    p9a=p9a,
                                    p9b=p9b,
                                    p10=p10,
                                    p11=p11,
                                    p12a=p12a,
                                    p12b=p12b,
                                    p13=p13,
                                    p14=p14,
                                    p15a=p15a,
                                    p15b=p15b,
                                    p16a=p16a,
                                    p16b=p16b,
                                    code= code,
                                    h18 = h18,
                                    h19 = h19, h20=h20,
                                    h17  = h17)
)

Morocco2017 <- writexl::write_xlsx(mar2017,"C:/Users/LENOVO/Documents/HappyGitwithR/steps_ssa_data/CVD_Paper_2023/stepsKGZ2023/Data/mar2017.xlsx",col_names = TRUE, format_headers = TRUE,)
mar2017 <-read_excel("mar2017.xlsx")

#9. Sao Tome and Principe
stp2019 <- read_dta("stata_files/stp2019.dta")

## Recode CVD dataset
table(stp2019$urbanrural) # check any other codes for rural and urban residence

#Extract required dataset
stp2019 <- with(stp2019, data.frame(country   = "STP", #"Sao Tome and Principe",
                                    reside    = if_else(urbanrural==2,1,0),
                                    survyear  = year,
                                    psu       = psu,
                                    stratum   = stratum,
                                    wstep1   = wstep1,
                                    wstep2 = wstep2,
                                    wstep3   = wstep3,                                
                                    sex       = if_else(sex == "Women" , 0, # "FEMALE"
                                                        1), # "Male"
                                    age       = age,
                                    occupation = if_else(c8 == 1, 0, # "Government employee"
                                                         if_else(c8 == 2 , 1, # "Non-Govt employee"
                                                                 if_else(c8 ==3 | c8 ==5 , 2, #Self-employed
                                                                         if_else(c8==4 | c8==8 | c8==9 , 3, # "Non-paid"
                                                                                 if_else(c8==6 , 3, # "Retired"
                                                                                         NA))))),
                                    edlev = if_else(c5 == 88 | c5 == 2 | c5 == 1, 0, # "None"
                                                    if_else( c5 == 3, 1, # "Primary"
                                                             if_else(c5 ==4 | c5 == 5, 2, #"Secondary"
                                                                     if_else( c5==6 | c5==7, 3, # "Tertiary"
                                                                              NA)))), 
                                    mstatus =if_else(c7 == 1, 0, # "Never married"
                                                     if_else(c7 == 2 | c7 == 6, 1, # "Currently married"
                                                             if_else(c7 == 3 | c7 == 4 | c5 ==5 , 2, #"Formerly married"
                                                                     NA))),  
                                    edyears =c4,
                                   # agegroup  = agerange,
                                   h12=h12,
                                   h13a=h13a,
                                   h13b=h13b,
                                   b8=b8,
                                   b9=b9,
                                   d1=d1,
                                   m11=m11,
                                   m12=m12,
                                   h1=h1,
                                   d11a=d11a, 
                                   d11b=d11b, 
                                   d11c=d11c, 
                                   d11d=d11d, 
                                   d11e=d11e, 
                                   d11f=d11f,
                                   h6=h6, 
                                   h7a=h7a, 
                                   m4a=m4a, 
                                   m5a=m5a, 
                                   m6a=m6a, 
                                   t1=t1, 
                                   t8=t8, 
                                   h3=h3, 
                                   h8=h8, 
                                   h9=h9, 
                                   h14=h14, 
                                   h20a=h20a, 
                                   h20b=h20b, 
                                   h20c=h20c, 
                                   h20d=h20d, 
                                   h20e=h20e, 
                                   h20f=h20f, 
                                   b1=b1, 
                                   b6=b6, 
                                   b8=b8,
                                    smoke=t1,
                                    a4 =a4,
                                    a6= a6,
                                    d5=d5,
                                    d2=d2,
                                    h2a=h2a,
                                    b5=b5,
                                    d3 = d3,
                                    d4=d4,
                                    p1=p1,
                                    p2=p2,
                                    p3a=p3a,
                                    p3b=p3b,
                                    p4=p4,
                                    p5=p5,
                                    p6a=p6a,
                                    p6b=p6b,
                                    p7=p7,
                                    p8=p8,
                                    p9a=p9a,
                                    p9b=p9b,
                                    p10=p10,
                                    p11=p11,
                                    p12a=p12a,
                                    p12b=p12b,
                                    p13=p13,
                                    p14=p14,
                                    p15a=p15a,
                                    p15b=p15b,
                                    p16a=p16a,
                                    p16b=p16b,
                                    code= code,
                                    h18 = h18,
                                    h19 = h19, h20=h20,
                                    h17  = h17)
)

SaoTome2019 <- writexl::write_xlsx(stp2019,"C:/Users/LENOVO/Documents/HappyGitwithR/steps_ssa_data/CVD_Paper_2023/stepsKGZ2023/Data/stp2019.xlsx",col_names = TRUE, format_headers = TRUE,)
stp2019 <-read_excel("stp2019.xlsx")

#10.Sudan
sdn2016 <- read_dta("stata_files/sdn2016.dta")

## Recode CVD dataset
table(sdn2016$stratum) # check any other codes for rural and urban residence

#Variable for CVD
sdn2016$h17 <- 0
sdn2016$h17[sdn2016$h17a==1 | sdn2016$h17b ==1] <-1


#Extract required dataset
sdn2016 <- with(sdn2016, data.frame(country   = "Sudan",
                                    reside    = if_else(stratum==1,1,0),
                                    survyear  = year,
                                    psu       = psu,
                                    stratum   = stratum,
                                    wstep1   = wstep1,
                                    wstep2 = wstep2,
                                    wstep3   = wstep3,                                
                                    sex       = if_else(sex == "Women" , 0, # "FEMALE"
                                                        1), # "Male"
                                    age       = age,
                                    occupation = if_else(c8 == 1, 0, # "Government employee"
                                                         if_else(c8 == 2 , 1, # "Non-Govt employee"
                                                                 if_else(c8 ==3 | c8 ==5 , 2, #Self-employed
                                                                         if_else(c8==4 | c8==8 | c8==9 , 3, # "Non-paid"
                                                                                 if_else(c8==6 , 3, # "Retired"
                                                                                         NA))))),
                                    edlev = if_else(c5 == 88 |  c5 == 1, 0, # "None"
                                                    if_else( c5 == 3 | c5==2, 1, # "Primary"
                                                             if_else(c5 ==4 | c5 == 5, 2, #"Secondary"
                                                                     if_else( c5==6 | c5==7, 3, # "Tertiary"
                                                                              NA)))), 
                                    mstatus =if_else(c7 == 1, 0, # "Never married"
                                                     if_else(c7 == 2 | c7 == 6, 1, # "Currently married"
                                                             if_else(c7 == 3 | c7 == 4 | c5 ==5 , 2, #"Formerly married"
                                                                     NA))), 
                                    edyears =c4,
                                    #agegroup  = agerange,
                                    b8=b8,
                                    b9=b9,
                                    d1=d1,
                                    h1=h1,
                                    d11a=d11a, 
                                    # d11b=d11b, 
                                    # d11c=d11c, 
                                    # d11d=d11d, 
                                    d11e=d11e, 
                                    d11f=d11f,
                                    h6=h6, 
                                    h7a=h7a, 
                                    m4a=m4a, 
                                    m5a=m5a, 
                                    m6a=m6a, 
                                    t1=t1, 
                                    t8=t8, 
                                    h3=h3, 
                                    h8=h8, 
                                    h9=h9, 
                                    h14=h14, 
                                    h20a=h20a, 
                                    h20b=h20b, 
                                    h20c=h20c, 
                                    h20d=h20d, 
                                    h20e=h20e, 
                                    h20f=h20f, 
                                    b1=b1, 
                                    b6=b6, 
                                    b8=b8,
                                    smoke=t1,
                                    a4 =a5,
                                    a6= a9,
                                    d5=d5,
                                    d2=d2,
                                    h2a=h2a,
                                    b5=b5,
                                    d3 = d3,
                                    d4=d4,
                                    p1=p1,
                                    p2=p2,
                                    p3a=p3a,
                                    p3b=p3b,
                                    p4=p4,
                                    p5=p5,
                                    p6a=p6a,
                                    p6b=p6b,
                                    p7=p7,
                                    p8=p8,
                                    p9a=p9a,
                                    p9b=p9b,
                                    p10=p10,
                                    p11=p11,
                                    p12a=p12a,
                                    p12b=p12b,
                                    p13=p13,
                                    p14=p14,
                                    p15a=p15a,
                                    p15b=p15b,
                                    p16a=p16a,
                                    p16b=p16b,
                                    code= code,
                                    h18 = h18,
                                    h19 = h19, h20=h20,
                                    h17  = h17)
)

Sudan2016 <- writexl::write_xlsx(sdn2016,"C:/Users/LENOVO/Documents/HappyGitwithR/steps_ssa_data/CVD_Paper_2023/stepsKGZ2023/Data/sdn2016.xlsx",col_names = TRUE, format_headers = TRUE,)

sdn2016 <-read_excel("sdn2016.xlsx")

#11. Uganda
uga2014 <- read_dta("stata_files/uga2014.dta")

## Recode CVD dataset
table(uga2014$stratum) # check any other codes for rural and urban residence
 

#Extract required dataset
uga2014 <- with(uga2014, data.frame(country   = "Uganda",
                                    reside    = if_else(stratum==1,1,0),
                                    survyear  = year,
                                    psu       = psu,
                                    stratum   = stratum,
                                    wstep1   = wstep1,
                                    wstep2 = wstep2,
                                    wstep3   = wstep3,                                
                                    sex       = if_else(sex == "Women" , 0, # "FEMALE"
                                                        1), # "Male"
                                    age       = age,
                                    occupation = if_else(c8 == 1, 0, # "Government employee"
                                                         if_else(c8 == 2 , 1, # "Non-Govt employee"
                                                                 if_else(c8 ==3 | c8 ==5 , 2, #Self-employed
                                                                         if_else(c8==4 | c8==8 | c8==9 , 3, # "Non-paid"
                                                                                 if_else(c8==6 , 3, # "Retired"
                                                                                         NA))))),
                                    edlev = if_else(c5 == 8 | c5 == 1, 0, # "None"
                                                    if_else( c5 == 2, 1, # "Primary"
                                                             if_else(c5==3 | c5 ==4 | c5 == 5, 2, #"Secondary"
                                                                     if_else( c5==6 | c5==7, 3, # "Tertiary"
                                                                              NA)))), 
                                    mstatus =if_else(c7 == 1, 0, # "Never married"
                                                     if_else(c7 == 2 | c7 == 6, 1, # "Currently married"
                                                             if_else(c7 == 3 | c7 == 4 | c5 ==5 , 2, #"Formerly married"
                                                                     NA))), 
                                    edyears =c4,
                                    #agegroup  = agerange,
                                    h12=h12,
                                    h13a=h13a,
                                    h13b=h13b,
                                    b8=b8,
                                    b9=b9,
                                    d1=d1,
                                    m11=m11,
                                    m12=m12,
                                    h1=h1,
                                    d11a=d11a, 
                                    d11b=d11b, 
                                    d11c=d11c, 
                                    d11d=d11d, 
                                    d11e=d11e, 
                                    d11f=d11f,
                                    h6=h6, 
                                    h7a=h7a, 
                                    m4a=m4a, 
                                    m5a=m5a, 
                                    m6a=m6a, 
                                    t1=t1, 
                                    t8=t8, 
                                    h3=h3, 
                                    h8=h8, 
                                    h9=h9, 
                                    h14=h14, 
                                    h20a=h20a, 
                                    h20b=h20b, 
                                    h20c=h20c, 
                                    h20d=h20d, 
                                    h20e=h20e, 
                                    h20f=h20f, 
                                    b1=b1, 
                                    b6=b6, 
                                    b8=b8,
                                    smoke=t1,
                                    a4 =a5,
                                    a6= a9,
                                    d5=d5,
                                    d2=d2,
                                    h2a=h2a,
                                    b5=b5,
                                    d3 = d3,
                                    d4=d4,
                                    p1=p1,
                                    p2=p2,
                                    p3a=p3a,
                                    p3b=p3b,
                                    p4=p4,
                                    p5=p5,
                                    p6a=p6a,
                                    p6b=p6b,
                                    p7=p7,
                                    p8=p8,
                                    p9a=p9a,
                                    p9b=p9b,
                                    p10=p10,
                                    p11=p11,
                                    p12a=p12a,
                                    p12b=p12b,
                                    p13=p13,
                                    p14=p14,
                                    p15a=p15a,
                                    p15b=p15b,
                                    p16a=p16a,
                                    p16b=p16b,
                                    code= code,
                                    h18 = h18,
                                    h19 = h19, h20=h20,
                                    h17  = h17)
)
Uganda2014 <- writexl::write_xlsx(uga2014,"C:/Users/LENOVO/Documents/HappyGitwithR/steps_ssa_data/CVD_Paper_2023/stepsKGZ2023/Data/uga2014.xlsx",col_names = TRUE, format_headers = TRUE,)
uga2014 <-read_excel("uga2014.xlsx")

#12. Zambia
zmb2017 <- read_dta("stata_files/zmb2017.dta")

## Recode CVD dataset
table(zmb2017$stratum) # check any other codes for rural and urban residence


#Extract required dataset
zmb2017 <- with(zmb2017, data.frame(country   = "Zambia",
                                    reside    = if_else(stratum==2,1,0),
                                    survyear  = year,
                                    psu       = psu,
                                    stratum   = stratum,
                                    wstep1   = wstep1,
                                    wstep2 = wstep2,
                                    wstep3   = wstep3,                                
                                    sex       = if_else(sex == "Women" , 0, # "FEMALE"
                                                        1), # "Male"
                                    age       = age,
                                    occupation = if_else(c8 == 1, 0, # "Government employee"
                                                         if_else(c8 == 2 , 1, # "Non-Govt employee"
                                                                 if_else(c8 ==3 | c8 ==5 , 2, #Self-employed
                                                                         if_else(c8==4 | c8==8 | c8==9 , 3, # "Non-paid"
                                                                                 if_else(c8==6 , 3, # "Retired"
                                                                                         NA))))),
                                    edlev = if_else(c5 == 88  | c5 == 1, 0, # "None"
                                                    if_else( c5 == 3 | c5 == 2, 1, # "Primary"
                                                             if_else(c5 ==4 | c5 == 5, 2, #"Secondary"
                                                                     if_else( c5==6 | c5==7, 3, # "Tertiary"
                                                                              NA)))), 
                                    mstatus =if_else(c7 == 1, 0, # "Never married"
                                                     if_else(c7 == 2 | c7 == 6, 1, # "Currently married"
                                                             if_else(c7 == 3 | c7 == 4 | c5 ==5 , 2, #"Formerly married"
                                                                     NA))), 
                                    edyears =c4,
                                   # agegroup  = agerange,
                                   h12=h12,
                                   h13a=h13a,
                                   h13b=h13b,
                                   b8=b8,
                                   b9=b9,
                                   d1=d1,
                                   m11=m11,
                                   m12=m12,
                                   h1=h1,
                                   d11a=d11a, 
                                   d11b=d11b, 
                                   d11c=d11c, 
                                   d11d=d11d, 
                                   d11e=d11e, 
                                   d11f=d11f,
                                   h6=h6, 
                                   h7a=h7a, 
                                   m4a=m4a, 
                                   m5a=m5a, 
                                   m6a=m6a, 
                                   t1=t1, 
                                   t8=t8, 
                                   h3=h3, 
                                   h8=h8, 
                                   h9=h9, 
                                   h14=h14, 
                                   h20a=h20a, 
                                   h20b=h20b, 
                                   h20c=h20c, 
                                   h20d=h20d, 
                                   h20e=h20e, 
                                   h20f=h20f, 
                                   b1=b1, 
                                   b6=b6, 
                                   b8=b8,
                                    smoke=t1,
                                    a4 =a5,
                                    a6= a9,
                                    d5=d5,
                                    d2=d2,
                                    h2a=h2a,
                                    b5=b5,
                                    d3 = d3,
                                    d4=d4,
                                    p1=p1,
                                    p2=p2,
                                    p3a=p3a,
                                    p3b=p3b,
                                    p4=p4,
                                    p5=p5,
                                    p6a=p6a,
                                    p6b=p6b,
                                    p7=p7,
                                    p8=p8,
                                    p9a=p9a,
                                    p9b=p9b,
                                    p10=p10, 
                                    p11=p11,
                                    p12a=p12a,
                                    p12b=p12b,
                                    p13=p13,
                                    p14=p14,
                                    p15a=p15a,
                                    p15b=p15b,
                                    p16a=p16a,
                                    p16b=p16b,
                                    code= code,
                                    h18 = h18,
                                    h19 = h19, h20=h20,
                                    h17  = h17)
)

zambia2017 <-writexl::write_xlsx(zmb2017,"C:/Users/LENOVO/Documents/HappyGitwithR/steps_ssa_data/CVD_Paper_2023/stepsKGZ2023/Data/zmb2017.xlsx",col_names = TRUE, format_headers = TRUE,)
zmb2017 <-read_excel("zmb2017.xlsx")

#Append data
datawt <-list.files(pattern = "\\.xlsx$") %>%
  map_dfr(read_excel)
  
#Remove unwanted datasets
remove(Algeria2016,Benin2015,Botswana2014,Ethiopia2015,Kenya2015,Morocco2017,Malawi2017,Sudan2016,SaoTome2019,Eswatini,Uganda2014,zambia2017)
remove(alg2016,bwa2014,ben2015,eth2015,ken2015,mar2017,mwi2017,sdn2016,stp2019,swz2014,uga2014,zmb2017,nrows)


#Merge datasets 
#Load alcohol policy data
alcohol_policy_africa <- read_dta("stata_files/alcohol_policy_africa.dta")
#Load country and region in Africa
country_STEPS_surveys_years <- read_dta("stata_files/country_STEPS_surveys_years.dta")
#Load tobacco policy
tobacco_policy_africa <- read_dta("stata_files/tobacco_policy_africa.dta")

tmerge <- merge(tobacco_policy_africa,country_STEPS_surveys_years,by=c("country"),all.x=T)
tmerge$code <-tmerge$code.x  

tmerge <-subset(tmerge,select = -c(code.x,code.y))


amerge <- merge(tmerge,alcohol_policy_africa,by=c("country"),all.x=T)
amerge$code <-amerge$code.x
amerge <-subset(amerge,select = -c(code.x,code.y))

amerge_2024 <- writexl::write_xlsx(amerge,"C:\\Users\\LENOVO\\Documents\\HappyGitwithR\\steps_ssa_data\\CVD_Analysis_2024\\Data\\amerge_2024.xlsx",col_names = TRUE, format_headers = TRUE,)


datawt <- merge(datawt,amerge, by=c("country"),all.x=T)


################################################################################
# CLEAN LOADED DATA
################################################################################

# Clean column names to lower case
datawt <- datawt %>% rename_all(tolower)

################################################################################
# CHECK LOADED DATA
################################################################################

datawt_names <- c("sex", "c1", "valid", "wstep1", "wstep2", "wstep3", "psu", 
                  "stratum", "agerange", "agerange2", "ur", "region", "tc_policy", "alc_policy")
#stratum=region 

# Check which names are not in the data before proceeding
missing_names <- setdiff(datawt_names, names(datawt))

if (length(missing_names) > 0) {
  warning(sprintf("%s doesn't contain: %s. Creating the missing variables.", 
                  deparse(substitute(datawt)),
                  paste(missing_names, collapse=", ")))
  
  datawt <- datawt %>% 
    mutate(
      # Create correct agerange variable (should be two ranges in KGZ) 
      agerange = factor(ifelse(age>=18 & age<=39, "18–39",
                               ifelse(age>=40 & age<=69, "40–69", NA))),
      # Create sex variable
      sex = factor(sex),
      
      # Create stratum variable to include regional disaggregation
      stratum = factor(stratum),
      
      # Create psu variable to include clusters
      psu = factor(psu),
      
      # Add agerange2 (18-39, 40-69) for Urban/Rural disaggregation
      agerange2 = factor(ifelse(age>=18 & age<=39, "18–39", 
                                ifelse(age>=40 & age<=69, "40–69", NA))),
      
      # Add region, ur variables as factors (specific for SSA countries)
      # Region column based on provided information from the country
      region = factor(country),
      ur = factor(reside),
      #Alcohol and tobacoo control policies
      #tc_policy = factor(tc_policy),
      #alc_policy = factor(restriction),    
      # Recode values in ur with dplyr (specific for KGZ)
      #ur = recode(ur, "1" = "Urban", "2" = "Rural"),
      # Create c1 variable after joining datasets for CardiovascularDiseaseRisk 
      # script as it requires c1 variable
      c1 = sex,
      # Create valid variable = consent signed, ages 18 to 69, and sex present
      valid = ifelse( (age >= 18 & age <= 69) 
                     & (sex == 1 | sex == 0), 1, 2)
    )
    
  } else {
    
    # NOTE: agerange should be changed to a factor because sometimes age ranges are
    # dropped if there are 0 values in data and .drop = FALSE argument in group_by function
    # does not work if it is not a factor.
    # The same applies to variables agerange2, region, sex and ur
    
    datawt <- datawt %>% 
      mutate(
        agerange = factor(agerange),
        agerange2 = factor(agerange2),
        region = factor(region),
        ur = factor(ur),
        tc_policy = factor(tc_policy),
        alc_policy = factor(alc_policy),
        sex = factor(sex)
    )

  }

################################################################################

#Exclude missing data on h17---history of CVD
MISSING <- is.na(datawt$h17) 
# Count the number of rows flagged for deletion
sum(MISSING)

# Use ! to include those that are NOT missing
datawt <- subset(datawt, 
                   subset = !MISSING)

# Subset to valid
datawt <- subset(datawt, valid == 1)

#-------------------------------------------------------------------------------

# Check vars
unique(datawt$agerange)
unique(datawt$agerange2) 
unique(datawt$sex)
unique(datawt$ur)
unique(datawt$region)
unique(datawt$tc_policy)
unique(datawt$alc_policy)
dim(datawt)
table(datawt$valid)

################################################################################
##Further data management
#0. Physical inactivity
datawt <- datawt %>%
  mutate(
    ### Clean Recode P1-P3
    # Mutate "a" (hr) and "b" (mins) at the same time
    p3b = replace(p3b, p3a==15 & (is.na(p3b) | p3b==0 | p3b==15 | p3b==77 | p3b==88 | p3b==99), 15),
    p3a = replace(p3a, p3a==15 & (is.na(p3b) | p3b==0 | p3b==15 | p3b==77 | p3b==88 | p3b==99), 0),
    p3b = replace(p3b, p3a==30 & (is.na(p3b) | p3b==0 | p3b==30 | p3b==77 | p3b==88 | p3b==99), 30),
    p3a = replace(p3a, p3a==30 & (is.na(p3b) | p3b==0 | p3b==30 | p3b==77 | p3b==88 | p3b==99), 0),
    p3b = replace(p3b, p3a==45 & (is.na(p3b) | p3b==0 | p3b==45 | p3b==77 | p3b==88 | p3b==99), 45),
    p3a = replace(p3a, p3a==45 & (is.na(p3b) | p3b==0 | p3b==45 | p3b==77 | p3b==88 | p3b==99), 0),
    p3b = replace(p3b, p3a==60 & (is.na(p3b) | p3b==0 | p3b==60 | p3b==77 | p3b==88 | p3b==99), 0),
    p3a = replace(p3a, p3a==60 & (is.na(p3b) | p3b==0 | p3b==60 | p3b==77 | p3b==88 | p3b==99), 1),
    p3b = replace(p3b, (p3a==7 & p3b==77) | (p3a==8 & p3b==88) | (p3a==9 & p3b==99), 0),
    p3a = replace(p3a, (p3a==7 & p3b==77) | (p3a==8 & p3b==88) | (p3a==9 & p3b==99), 0),
    p3b = replace(p3b, p3b==77 | p3b==88 | p3b==99, 0),
    p3a = replace(p3a, p3a==77 | p3a==88 | p3a==99, 0),
    # Recode variables into minutes only
    p3amin = ifelse(is.na(p3a), 0, p3a*60),
    p3bmin = ifelse(is.na(p3b), 0, p3b),
    p3 = p3amin + p3bmin,
    # Clean P1-P3
    p2 = replace(p2, is.na(p2) | p2==99, 0),
    p2cln = if_else((p1==1 & p2>0 & p2<8) | (p1==2 & p2==0), 1, 2, missing = 2),
    p3cln = if_else(p2cln==1 & p2>0 & p2<8 & p3>9 & p3<961, 1, 2, missing = 2),
    p3cln = replace(p3cln, p2cln==1 & p2==0 & p3==0, 1),
    p1t3cln = if_else(p3cln==1 & valid==1, 1, 2, missing = 2),
    p1t3cln = replace(p1t3cln, is.na(p1) & p2==0 & p3==0 & valid==1, 1),
    ### Clean Recode P4-P6
    p6a = replace(p6a, p6a==15 & (is.na(p6b) | p6b==0 | p6b==15 | p6b==77 | p6b==88 | p6b==99), 0),
    p6b = replace(p6b, p6a==15 & (is.na(p6b) | p6b==0 | p6b==15 | p6b==77 | p6b==88 | p6b==99), 15),
    p6a = replace(p6a, p6a==30 & (is.na(p6b) | p6b==0 | p6b==30 | p6b==77 | p6b==88 | p6b==99), 0),
    p6b = replace(p6b, p6a==30 & (is.na(p6b) | p6b==0 | p6b==30 | p6b==77 | p6b==88 | p6b==99), 30),
    p6a = replace(p6a, p6a==45 & (is.na(p6b) | p6b==0 | p6b==45 | p6b==77 | p6b==88 | p6b==99), 0),
    p6b = replace(p6b, p6a==45 & (is.na(p6b) | p6b==0 | p6b==45 | p6b==77 | p6b==88 | p6b==99), 45),
    p6a = replace(p6a, p6a==60 & (is.na(p6b) | p6b==0 | p6b==60 | p6b==77 | p6b==88 | p6b==99), 1),
    p6b = replace(p6b, p6a==60 & (is.na(p6b) | p6b==0 | p6b==60 | p6b==77 | p6b==88 | p6b==99), 0),
    p6a = replace(p6a, (p6a==7 & p6b==77) | (p6a==8 & p6b==88) | (p6a==9 & p6b==99), 0),
    p6b = replace(p6b, (p6a==7 & p6b==77) | (p6a==8 & p6b==88) | (p6a==9 & p6b==99), 0),
    p6b = replace(p6b, p6b==77 | p6b==88 | p6b==99, 0),
    p6a = replace(p6a, p6a==77 | p6a==88 | p6a==99, 0),
    # Recode variables into minutes only
    p6amin = ifelse(is.na(p6a), 0, p6a*60),
    p6bmin = ifelse(is.na(p6b), 0, p6b),
    p6 = p6amin + p6bmin,
    # Clean P4-P6
    p5 = replace(p5, is.na(p5) | p5==99, 0),
    p5cln = if_else((p4==1 & p5>0 & p5<8) | (p4==2 & p5==0), 1, 2, missing = 2),
    p6cln = if_else(p5cln==1 & p5>0 & p5<8 & p6>9 & p6<961, 1, 2, missing = 2),
    p6cln = replace(p6cln, p5cln==1 & p5==0 & p6==0, 1),
    p4t6cln = if_else(p6cln==1 & valid==1, 1, 2, missing = 2),
    p4t6cln = replace(p4t6cln, is.na(p4) & p5==0 & p6==0 & valid==1, 1),
    ### Clean Recode P7-P9
    p9a = replace(p9a, p9a==15 & (is.na(p9b) | p9b==0 | p9b==15 | p9b==77 | p9b==88 | p9b==99), 0),
    p9b = replace(p9b, p9a==15 & (is.na(p9b) | p9b==0 | p9b==15 | p9b==77 | p9b==88 | p9b==99), 15),
    p9a = replace(p9a, p9a==30 & (is.na(p9b) | p9b==0 | p9b==30 | p9b==77 | p9b==88 | p9b==99), 0),
    p9b = replace(p9b, p9a==30 & (is.na(p9b) | p9b==0 | p9b==30 | p9b==77 | p9b==88 | p9b==99), 30),
    p9a = replace(p9a, p9a==45 & (is.na(p9b) | p9b==0 | p9b==45 | p9b==77 | p9b==88 | p9b==99), 0),
    p9b = replace(p9b, p9a==45 & (is.na(p9b) | p9b==0 | p9b==45 | p9b==77 | p9b==88 | p9b==99), 45),
    p9a = replace(p9a, p9a==60 & (is.na(p9b) | p9b==0 | p9b==60 | p9b==77 | p9b==88 | p9b==99), 1),
    p9b = replace(p9b, p9a==60 & (is.na(p9b) | p9b==0 | p9b==60 | p9b==77 | p9b==88 | p9b==99), 0),
    p9a = replace(p9a, (p9a==7 & p9b==77) | (p9a==8 & p9b==88) | (p9a==9 & p9b==99), 0),
    p9b = replace(p9b, (p9a==7 & p9b==77) | (p9a==8 & p9b==88) | (p9a==9 & p9b==99), 0),
    p9b = replace(p9b, p9b==77 | p9b==88 | p9b==99, 0),
    p9a = replace(p9a, p9a==77 | p9a==88 | p9a==99, 0),
    # Recode variables into minutes only
    p9amin = ifelse(is.na(p9a), 0, p9a*60),
    p9bmin = ifelse(is.na(p9b), 0, p9b),
    p9 = p9amin + p9bmin,
    # Clean p7-p9
    p8 = replace(p8, is.na(p8) | p8==99, 0),
    p8cln = if_else((p7==1 & p8>0 & p8<8) | (p7==2 & p8==0), 1, 2, missing = 2),
    p9cln = if_else(p8cln==1 & p8>0 & p8<8 & p9>9 & p9<961, 1, 2, missing = 2),
    p9cln = replace(p9cln, p8cln==1 & p8==0 & p9==0, 1),
    p7t9cln = if_else(p9cln==1 & valid==1, 1, 2, missing = 2),
    p7t9cln = replace(p7t9cln, is.na(p7) & p8==0 & p9==0 & valid==1, 1),
    ### Clean Recode P10-P12
    p12a = replace(p12a, p12a==15 & (is.na(p12b) | p12b==0 | p12b==15 | p12b==77 | p12b==88 | p12b==99), 0),
    p12b = replace(p12b, p12a==15 & (is.na(p12b) | p12b==0 | p12b==15 | p12b==77 | p12b==88 | p12b==99), 15),
    p12a = replace(p12a, p12a==30 & (is.na(p12b) | p12b==0 | p12b==30 | p12b==77 | p12b==88 | p12b==99), 0),
    p12b = replace(p12b, p12a==30 & (is.na(p12b) | p12b==0 | p12b==30 | p12b==77 | p12b==88 | p12b==99), 30),
    p12a = replace(p12a, p12a==45 & (is.na(p12b) | p12b==0 | p12b==45 | p12b==77 | p12b==88 | p12b==99), 0),
    p12b = replace(p12b, p12a==45 & (is.na(p12b) | p12b==0 | p12b==45 | p12b==77 | p12b==88 | p12b==99), 45),
    p12a = replace(p12a, p12a==60 & (is.na(p12b) | p12b==0 | p12b==60 | p12b==77 | p12b==88 | p12b==99), 1),
    p12b = replace(p12b, p12a==60 & (is.na(p12b) | p12b==0 | p12b==60 | p12b==77 | p12b==88 | p12b==99), 0),
    p12a = replace(p12a, (p12a==7 & p12b==77) | (p12a==8 & p12b==88) | (p12a==9 & p12b==99), 0),
    p12b = replace(p12b, (p12a==7 & p12b==77) | (p12a==8 & p12b==88) | (p12a==9 & p12b==99), 0),
    p12b = replace(p12b, p12b==77 | p12b==88 | p12b==99, 0),
    p12a = replace(p12a, p12a==77 | p12a==88 | p12a==99, 0),
    # Recode variables into minutes only
    p12amin = ifelse(is.na(p12a), 0, p12a*60),
    p12bmin = ifelse(is.na(p12b), 0, p12b),
    p12 = p12amin + p12bmin,
    # Clean p10-p12
    p11 = replace(p11, is.na(p11) | p11==99, 0),
    p11cln = if_else((p10==1 & p11>0 & p11<8) | (p10==2 & p11==0), 1, 2, missing = 2),
    p12cln = if_else(p11cln==1 & p11>0 & p11<8 & p12>9 & p12<961, 1, 2, missing = 2),
    p12cln = replace(p12cln, p11cln==1 & p11==0 & p12==0, 1),
    p10t12cln = if_else(p12cln==1 & valid==1, 1, 2, missing = 2),
    p10t12cln = replace(p10t12cln, is.na(p10) & p11==0 & p12==0 & valid==1, 1),
    ### Clean Recode P13-P15
    p15a = replace(p15a, p15a==15 & (is.na(p15b) | p15b==0 | p15b==15 | p15b==77 | p15b==88 | p15b==99), 0),
    p15b = replace(p15b, p15a==15 & (is.na(p15b) | p15b==0 | p15b==15 | p15b==77 | p15b==88 | p15b==99), 15),
    p15a = replace(p15a, p15a==30 & (is.na(p15b) | p15b==0 | p15b==30 | p15b==77 | p15b==88 | p15b==99), 0),
    p15b = replace(p15b, p15a==30 & (is.na(p15b) | p15b==0 | p15b==30 | p15b==77 | p15b==88 | p15b==99), 30),
    p15a = replace(p15a, p15a==45 & (is.na(p15b) | p15b==0 | p15b==45 | p15b==77 | p15b==88 | p15b==99), 0),
    p15b = replace(p15b, p15a==45 & (is.na(p15b) | p15b==0 | p15b==45 | p15b==77 | p15b==88 | p15b==99), 45),
    p15a = replace(p15a, p15a==60 & (is.na(p15b) | p15b==0 | p15b==60 | p15b==77 | p15b==88 | p15b==99), 1),
    p15b = replace(p15b, p15a==60 & (is.na(p15b) | p15b==0 | p15b==60 | p15b==77 | p15b==88 | p15b==99), 0),
    p15a = replace(p15a, (p15a==7 & p15b==77) | (p15a==8 & p15b==88) | (p15a==9 & p15b==99), 0),
    p15b = replace(p15b, (p15a==7 & p15b==77) | (p15a==8 & p15b==88) | (p15a==9 & p15b==99), 0),
    p15b = replace(p15b, p15b==77 | p15b==88 | p15b==99, 0),
    p15a = replace(p15a, p15a==77 | p15a==88 | p15a==99, 0),
    # Recode variables into minutes only
    p15amin = ifelse(is.na(p15a), 0, p15a*60),
    p15bmin = ifelse(is.na(p15b), 0, p15b),
    p15 = p15amin + p15bmin,
    # Clean p13-p15
    p14 = replace(p14, is.na(p14) | p14==99, 0),
    p14cln = if_else((p13==1 & p14>0 & p14<8) | (p13==2 & p14==0), 1, 2, missing = 2),
    p15cln = if_else(p14cln==1 & p14>0 & p14<8 & p15>9 & p15<961, 1, 2, missing = 2),
    p15cln = replace(p15cln, p14cln==1 & p14==0 & p15==0, 1),
    p13t15cln = if_else(p15cln==1 & valid==1, 1, 2, missing = 2),
    p13t15cln = replace(p13t15cln, is.na(p13) & p14==0 & p15==0 & valid==1, 1)
    
  )


datawt <- datawt %>%
  mutate(
    p16a = replace(p16a, p16a==15 & (is.na(p16b) | p16b==0 | p16b==15 | p16b==77 | p16b==88 | p16b==99), 0),
    p16b = replace(p16b, p16a==15 & (is.na(p16b) | p16b==0 | p16b==15 | p16b==77 | p16b==88 | p16b==99), 15),
    p16a = replace(p16a, p16a==30 & (is.na(p16b) | p16b==0 | p16b==30 | p16b==77 | p16b==88 | p16b==99), 0),
    p16b = replace(p16b, p16a==30 & (is.na(p16b) | p16b==0 | p16b==30 | p16b==77 | p16b==88 | p16b==99), 30),
    p16a = replace(p16a, p16a==45 & (is.na(p16b) | p16b==0 | p16b==45 | p16b==77 | p16b==88 | p16b==99), 0),
    p16b = replace(p16b, p16a==45 & (is.na(p16b) | p16b==0 | p16b==45 | p16b==77 | p16b==88 | p16b==99), 45),
    p16a = replace(p16a, p16a==60 & (is.na(p16b) | p16b==0 | p16b==60 | p16b==77 | p16b==88 | p16b==99), 1),
    p16b = replace(p16b, p16a==60 & (is.na(p16b) | p16b==0 | p16b==60 | p16b==77 | p16b==88 | p16b==99), 0),
    p16a = replace(p16a, (p16a==7 & p16b==77) | (p16a==8 & p16b==88) | (p16a==9 & p16b==99), 0),
    p16b = replace(p16b, (p16a==7 & p16b==77) | (p16a==8 & p16b==88) | (p16a==9 & p16b==99), 0),
    p16b = replace(p16b, p16b==77 | p16b==88 | p16b==99, 0),
    p16a = replace(p16a, p16a==77 | p16a==88 | p16a==99, 0),
    # Recode variables into minutes only
    p16amin = ifelse(p16a>=0 & p16a<=24, p16a*60, NA),
    p16bmin = ifelse(p16b>=0 & p16b<=60, p16b, NA),
    p16 = p16amin + p16bmin,
    # Clean P16
    p16cln = if_else(p16>=0 & p16<1441, 1, 2, missing = 2)
  )

datawt <- datawt %>%
  mutate(
    work = factor(if_else(p1==1 | p4==1, "did work activity", "did no work activity", 
                          missing = "did no work activity"),
                  levels = c("did work activity", "did no work activity")),
    trans = factor(if_else(p7==1, "did transport activity", "did no transport activity", 
                           missing = "did no transport activity"),
                   levels = c("did transport activity", "did no transport activity")),
    rec = factor(if_else(p10==1 | p13==1, "did recreation activity", "did no recreation activity", 
                         missing = "did no recreation activity"),
                 levels = c("did recreation activity", "did no recreation activity"))
  ) 

datawt <- datawt %>%
  mutate(
    p1t3 = ifelse(p1t3cln==1, p2*p3*8, NA),
    p4t6 = ifelse(p4t6cln==1, p5*p6*4, NA),
    p7t9 = ifelse(p7t9cln==1, p8*p9*4, NA),
    p10t12 = ifelse(p10t12cln==1, p11*p12*8, NA),
    p13t15 = ifelse(p13t15cln==1, p14*p15*4, NA),
    ptotal = p1t3+p4t6+p7t9+p10t12+p13t15,
    c = factor(if_else(ptotal>=600, "2) meets recs", "1) doesn't meet recs", 
                       missing = "1) doesn't meet recs"),
               levels = c("1) doesn't meet recs", "2) meets recs")),
    # Calculate additional ≥1200 MET-minutes/week
    mets1200 = factor(if_else(ptotal>=1200, "2) meets recs", "1) doesn't meet recs", 
                              missing = "1) doesn't meet recs"),
                      levels = c("1) doesn't meet recs", "2) meets recs"))
  )

datawt  <- datawt  %>%
  mutate(
    p1t3 = ifelse(p1t3cln==1, p2*p3*8, NA),
    p4t6 = ifelse(p4t6cln==1, p5*p6*4, NA),
    p7t9 = ifelse(p7t9cln==1, p8*p9*4, NA),
    p10t12 = ifelse(p10t12cln==1, p11*p12*8, NA),
    p13t15 = ifelse(p13t15cln==1, p14*p15*4, NA),
    ptotal = p1t3+p4t6+p7t9+p10t12+p13t15,
    c = ifelse((p2+p5+p8+p11+p14)>6 & ptotal>2999, "3) High level", NA),
    c = replace(c, (p2+p11)>2 & ptotal>1499, "3) High level"),
    c = replace(c, is.na(c) & ((p2+p5+p8+p11+p14)>=5) & ptotal>=600, "2) Moderate level"),
    c = replace(c, is.na(c) & ((p2+p11)==3 | (p2+p11)==4) & p12>=20 & p3>=20, "2) Moderate level"),
    c = replace(c, is.na(c) & p2>=3 & p11>=3 & (p12>=20 | p3>=20), "2) Moderate level"),
    c = replace(c, is.na(c) & ((p2>=3 & p11<3 & p3>=20) | (p11>=3 & p2<3 & p12>=20)), "2) Moderate level"),
    c = replace(c, is.na(c) & (p5+p8+p14)>=5 & ((p5*p6)+(p8*p9)+(p14*p15))>=150, "2) Moderate level"),
    c = replace(c, p1t3cln==1 & p4t6cln==1 & p7t9cln==1 & p10t12cln==1 & p13t15cln==1 & is.na(c), "1) Low Level"),
    c = factor(c, levels = c("1) Low Level", "2) Moderate level", "3) High level")),
    cln = if_else(p1t3cln==1 & p4t6cln==1 & p7t9cln==1 & p10t12cln==1 & 
                    p13t15cln==1, 1, 2, missing = 2),
    cln = replace(cln, is.na(p1) & is.na(p4) & is.na(p7) & is.na(p10) & is.na(p13), 2)
  )

datawt$pa <- datawt$c
var_label(datawt$pa) <- "Physical activity"  


#High cholesterol history 
datawt  <- datawt  %>% 
  mutate(
    h12cln = if_else(h12==1 | h12==2, 1, 2, missing = 2),
    h13acln = if_else(h12==1 & (h13a==1 | h13a==2), 1, 2, missing = 2),
    h13acln = replace(h13acln, h12==2 & (h13a==2 | is.na(h13a)), 1),
    h13bcln = if_else(h13a==1 & (h13b==1 | h13b==2), 1, 2, missing = 2),
    h13bcln = replace(h13bcln, h13a==2 & (h13b==2 | is.na(h13b)), 1),
    h13bcln = replace(h13bcln, h12==2 & (h13b==2 | is.na(h13b)), 1),
    cln = if_else(h12cln==1 & h13acln==1 & h13bcln==1 & valid==1, 1, 2, missing = 2),
    h14cln = if_else(h12==1 & h13a==1 & (h14==1 | h14==2) & valid==1, 1, 2, missing = 2),
    c = factor(
      case_when(
        h13b==1 ~ "4) diagnosed within past 12 months",
        h13a==1 & h13b==2 ~ "3) diagnosed, but not within past 12 months", 
        h13a==2 ~ "2) measured, not diagnosed",
        h12==2 ~ "1) never measured",
        TRUE ~ NA_character_
      ), 
      levels = c("1) never measured", "2) measured, not diagnosed",
                 "3) diagnosed, but not within past 12 months",
                 "4) diagnosed within past 12 months")
    ),
    d = factor(ifelse(h14==1, "1) taking meds", 
                      ifelse(h14==2, "2) not taking meds", NA)),
               levels = c("1) taking meds", "2) not taking meds")),
    # update 2023-10-17: added for question h13a only as ever diagnosed
    h13acln2 = if_else(h12==1 & (h13a==1 | h13a==2), 1, 2, missing = 2),
    e = factor(ifelse(h13a==1, "1) was told",
                      ifelse(h13a==2, "2) was not told", NA)),
               levels = c("1) was told", "2) was not told"))
  )
#High blood cholesterol
datawt$b8 <- if_else(datawt$b8<10,datawt$b8,datawt$b8/100)  #Manipulate and make uniform the values for B8
datawt <- datawt %>%
  mutate(
    cln = if_else(b8>=2 & b8<=12, 1, 2, missing = 2),
    c = ifelse(b8<5.0, "total cholesterol < 5.0", NA),
    d = ifelse(b8<5.0, "total cholesterol < 6.2", NA),
    e = ifelse(b8<5.0, "total cholesterol < 5.0", NA),
    f = ifelse(b8<5.0, "total cholesterol < 6.2", NA),
    c = replace(c, b8>=5.0 & b8<6.2, "total cholesterol >= 5.0 or on meds"),
    d = replace(d, b8>=5.0 & b8<6.2, "total cholesterol < 6.2"),
    e = replace(e, b8>=5.0 & b8<6.2, "total cholesterol >= 5.0"),
    f = replace(f, b8>=5.0 & b8<6.2, "total cholesterol < 6.2"),
    c = replace(c, b8>=6.2, "total cholesterol >= 5.0 or on meds"),
    d = replace(d, b8>=6.2, "total cholesterol >= 6.2 or on meds"),
    e = replace(e, b8>=6.2, "total cholesterol >= 5.0"),
    f = replace(f, b8>=6.2, "total cholesterol >= 6.2"),
    c = replace(c, b9==1, "total cholesterol >= 5.0 or on meds"),
    d = replace(d, b9==1, "total cholesterol >= 6.2 or on meds"),
    b8mg = b8*38.67,
    c = factor(c, levels = c("total cholesterol < 5.0", "total cholesterol >= 5.0 or on meds")),
    d = factor(d, levels = c("total cholesterol < 6.2", "total cholesterol >= 6.2 or on meds")),
    e = factor(e, levels = c("total cholesterol < 5.0", "total cholesterol >= 5.0")),
    f = factor(f, levels = c("total cholesterol < 6.2", "total cholesterol >= 6.2"))
  )

#CVD risk
datawt <- datawt %>%
  mutate(
    t10 = NA,
    t11 = 10,
    t11type = "years",
    smokecln = if_else(t1==1 | t1==2, 1, 2, missing = 2),
    smokecln = replace(smokecln, t1==2 & is.na(t8), 2),
    smoker = if_else(t1==1, 1, 2, missing = 2),
    # Calculate T11b (months) AND T11c (weeks) into years
    t11a = ifelse(t11type>="years", t11, NA),
    t11byrs = ifelse(t11type>="months", t11/12, NA),
    t11cyrs = ifelse(t11type>="weeks", t11/52, NA),
    t10cln = if_else(t10>=7 & t10<=69, 1, 2, missing = 2),
    t11acln = if_else(t11a>0 & t11a<=61, 1, 2, missing = 2),
    t11bcln = if_else(is.na(t11byrs), 2, 1, missing = 1),
    t11ccln = if_else(is.na(t11cyrs), 2, 1, missing = 1),
    # Order the priority of responses so that T10 takes precedence over other responses
    stop = ifelse(t11ccln==1, t11cyrs, NA),
    stop = replace(stop, t11bcln==1, t11byrs),
    stop = replace(stop, t11acln==1, t11a),
    stop = replace(stop, t10cln==1, age-t10),
    smokecln = replace(smokecln, t10cln==2 & t11acln==2 & t11bcln==2 & t11ccln==2 & t1==2 & t8==1, 2),
    smokecln = replace(smokecln, t1==2 & t8==1 & (is.na(stop) | stop>61 | stop<0), 2),
    smoker = replace(smoker, stop<=1, 1),
    m4acln = if_else(m4a>=40 & m4a<=300, 1, 2, missing = 2),
    m5acln = if_else(m5a>=40 & m5a<=300, 1, 2, missing = 2),
    m6acln = if_else(m6a>=40 & m6a<=300, 1, 2, missing = 2),
    sbp = ifelse((m4acln==1 | m4acln==2) & m5acln==1 & m6acln==1, (m5a + m6a)/2, NA),
    sbp = ifelse(m4acln==1 & m5acln==2 & m6acln==1, (m4a + m6a)/2, sbp),
    sbp = ifelse(m4acln==1 & m5acln==1 & m6acln==2, (m4a + m5a)/2, sbp),
    sbpcln = if_else((sbp<40 | sbp>300) | is.na(sbp), 2, 1, missing = 1),
    chol = ifelse(b8>=2 & b8<=12, b8, NA),
    cholcln = if_else(b8>=2 & b8<=12, 1, 2, missing = 2),
    diabcln = if_else(b5>=1 & b5<=35 & b1==2, 1, 2, missing = 2),
    diab = if_else(b5>=6.1 | b6==1, 1, 2, missing = 2),
    diabcln = replace(diabcln, h6==1 & h7a==1, 1),
    diab = replace(diab, h6==1 & h7a==1, 1),
    cln = if_else(diabcln==1 & cholcln==1 & sbpcln==1 & smokecln==1 & valid==1, 1, 2, missing = 2),
    highrisk = 2,
    # MALES WITH DIABETES
    highrisk = replace(highrisk, c1==1 & (age>=40 & age<50) & smoker==1 & diab==1 & ((chol>=8 & sbp>=140) | (chol>=7 & sbp>=160) | (chol>=4 & sbp>=180)), 1),
    highrisk = replace(highrisk, c1==1 & (age>=50 & age<60) & smoker==1 & diab==1 & ((chol>=8 & sbp>=140) | (chol>=6 & sbp>=160) | (chol>=4 & sbp>=180)), 1),
    highrisk = replace(highrisk, c1==1 & (age>=60 & age<70) & smoker==1 & diab==1 & ((chol>=8 & sbp>=120) | (chol>=7 & sbp>=140)), 1),
    highrisk = replace(highrisk, c1==1 & (age>=60 & age<70) & smoker==1 & diab==1 & ((chol>=5 & sbp>=160) | (chol>=4 & sbp>=180)), 1),
    highrisk = replace(highrisk, c1==1 & (age>=40 & age<50) & smoker==2 & diab==1 & ((chol>=8 & sbp>=160) | (chol>=6 & sbp>=180)), 1),
    highrisk = replace(highrisk, c1==1 & (age>=50 & age<60) & smoker==2 & diab==1 & ((chol>=8 & sbp>=160) | (chol>=5 & sbp>=180)), 1),
    highrisk = replace(highrisk, c1==1 & (age>=60 & age<70) & smoker==2 & diab==1 & ((chol>=8 & sbp>=140) | (chol>=7 & sbp>=160) | (chol>=4 & sbp>=180)), 1),
    # FEMALES WITH DIABETES
    highrisk = replace(highrisk, c1==2 & (age>=40 & age<60) & smoker==1 & diab==1 & ((chol>=8 & sbp>=140) | (chol>=7 & sbp>=160) | (chol>=4 & sbp>=180)), 1),
    highrisk = replace(highrisk, c1==2 & (age>=60 & age<70) & smoker==1 & diab==1 & ((chol>=8 & sbp>=140) | (chol>=6 & sbp>=160) | (chol>=4 & sbp>=180)), 1),
    highrisk = replace(highrisk, c1==2 & (age>=40 & age<60) & smoker==2 & diab==1 & ((chol>=8 & sbp>=160) | (chol>=5 & sbp>=180)), 1),
    highrisk = replace(highrisk, c1==2 & (age>=60 & age<70) & smoker==2 & diab==1 & ((chol>=8 & sbp>=140) | (chol>=7 & sbp>=160) | (chol>=4 & sbp>=180)), 1),
    # MEN WITHOUT DIABETES
    highrisk = replace(highrisk, c1==1 & (age>=40 & age<50) & smoker==1 & diab==2 & ((chol>=8 & sbp>=160) | (chol>=6 & sbp>=180)), 1),
    highrisk = replace(highrisk, c1==1 & (age>=50 & age<60) & smoker==1 & diab==2 & ((chol>=8 & sbp>=160) | (chol>=5 & sbp>=180)), 1),
    highrisk = replace(highrisk, c1==1 & (age>=60 & age<70) & smoker==1 & diab==2 & ((chol>=8 & sbp>=140) | (chol>=7 & sbp>=160) | (chol>=4 & sbp>=180)), 1),
    highrisk = replace(highrisk, c1==1 & (age>=40 & age<60) & smoker==2 & diab==2 & (chol>=7 & sbp>=180), 1),
    highrisk = replace(highrisk, c1==1 & (age>=60 & age<70) & smoker==2 & diab==2 & ((chol>=8 & sbp>=160) | (chol>=5 & sbp>=180)), 1),
    # WOMEN WITHOUT DIABETES
    highrisk = replace(highrisk, c1==2 & (age>=40 & age<70) & smoker==1 & diab==2 & ((chol>=8 & sbp>=160) | (chol>=5 & sbp>=180)), 1),
    highrisk = replace(highrisk, c1==2 & (age>=40 & age<70) & smoker==2 & diab==2 & (chol>=7 & sbp>=180), 1),
    highrisk = replace(highrisk, h17==1, 1),
    ###
    c = factor(ifelse(highrisk==1, "risk 30% or more or has CVD", 
                      ifelse(highrisk==2, "risk less than 30%", NA)),
               levels = c("risk 30% or more or has CVD", "risk less than 30%")),
    drugs = if_else(h3==1 | h8==1 | h9==1 | h14==1 | h18==1 | h19==1, 1, 2, missing = 2),
    counseling = if_else(h20a==1 | h20b==1 | h20c==1 | h20d==1 | h20e==1 | h20f==1, 1, 2, missing = 2),
    d = factor(if_else(drugs==1 & counseling==1, "1) received drug therapy and counseling", 
                       "2) did not receive drug therapy and counseling", missing = "2) did not receive drug therapy and counseling"),
               levels = c("1) received drug therapy and counseling", "2) did not receive drug therapy and counseling")),
    agerangecvd = factor(ifelse(age>=40 & age<=54, "40–54", 
                                ifelse(age>=55 & age<=69, "55–69", NA)),
                         levels = c("40–54", "55–69")),
    cln = replace(cln, age<40, 2)
  )

#Generate some useful variables
#1. Harmful Alcohol Use
datawt$alcharm =if_else(datawt$a4==1 & datawt$a6 >1 & datawt$sex==0,1,
                        if_else(datawt$a4==1 & datawt$a6 >2 & datawt$sex==1,1,0))


datawt$alcharm[is.na(datawt$alcharm)] <-0

datawt$alcharm <- factor(datawt$alcharm,
                                    levels = c(0,1),
                                    labels = c("No", "Yes"))

var_label(datawt$alcharm) <- "Harmful alcohol use"

#Current tobacco smoking status
datawt$smok <-datawt$t1
datawt$smok[is.na(datawt$smok)] <- 0
datawt$smok[datawt$smok==2] <- 0
datawt$smok <- factor(datawt$smok,
                                    levels = c(0,1),
                                    labels = c("No", "Yes"))

var_label(datawt$smok) <- "Currently smokes tobacco"

#Smoking history
datawt$smokehis <- 0
datawt$smokehis[datawt$t8==1 & datawt$smokehis==0] <- 1
datawt$smokehis[datawt$t1==1 & datawt$smokehis==0] <- 2

datawt$smokehis <- factor(datawt$smokehis,
                    levels = c(0,1,2),
                    labels = c("Never", "Previous","Current"))

var_label(datawt$smokehis) <- "Smoking history"

#Sex
datawt$gender <- datawt$sex
datawt$gender <- factor(datawt$gender,
                          levels = c(0,1),
                          labels = c("Female", "Male"))

var_label(datawt$gender) <- "Gender"
 
# #Creating age groups
datawt <- datawt %>%
  mutate(
    age_cat = age_categories(             # create new column
      age,                            # numeric column to make groups from
      breakers = c(15,        # break points (18+)
                   20, 25, 30, 35, 40,45, 50, 55,60,65)))
var_label(datawt$age_cat) <- "Age groups"


#High salt intake
datawt$highsalt <- if_else(datawt$d5 <3,1,0)
datawt$highsalt <- factor(datawt$highsalt,
                        levels = c(0,1),
                        labels = c("No", "Yes"))

var_label(datawt$highsalt) <- "High salt intake"


#Low fruit intake
datawt$lfi <- if_else(datawt$d1 <5,1,0)
datawt$lfi <- factor(datawt$lfi,
                          levels = c(0,1),
                          labels = c("No", "Yes"))

var_label(datawt$lfi) <- "Low fruit intake" 

#Low vegetable uptake
datawt$lvi <- if_else(datawt$d3 <5,1,0)
datawt$lvi <- factor(datawt$lvi,
                     levels = c(0,1),
                     labels = c("No", "Yes"))

var_label(datawt$lvi) <- "Low vegetable intake" 

#High blood pressure
datawt$bp <- datawt$h2a
datawt$bp[datawt$bp==2 | is.na(datawt$bp)] <-0
datawt$bp <- factor(datawt$bp,
                          levels = c(0,1),
                          labels = c("No", "Yes"))
 
var_label(datawt$bp) <- "Has high BP"


#High blood sugar
datawt$dbm <- datawt$b1 
datawt$dbm[datawt$dbm==2] <-0
datawt$dbm <- factor(datawt$dbm,
                    levels = c(0,1),
                    labels = c("No", "Yes"))

var_label(datawt$dbm) <- "High blood sugar"


#Occupation
datawt$occupation <- factor(datawt$occupation,
                    levels = c(0,1,2,3),
                    labels = c("Government employee", "Non-Govt employee","Self-employed","Non-paid/Retired"))

var_label(datawt$occupation) <- "Occupation"

#Highest education level
datawt$edlev <- factor(datawt$edlev,
                            levels = c(0,1,2,3),
                            labels = c("None", "Primary","Secondary","Tertiary"))

var_label(datawt$edlev) <- "Occupation"


#Marital status
datawt$mstatus <- factor(datawt$mstatus,
                       levels = c(0,1,2),
                       labels = c("Never married", "Currently married","Formerly married"))

var_label(datawt$mstatus) <- "Marital status"

#Area of residence
datawt$residence <- factor(datawt$reside,
                          levels = c(0,1),
                          labels = c("Rural", "Urban"))

var_label(datawt$residence) <- "Area of residence" 


#Body mass index 
datawt$bmi <- datawt$m12/(datawt$m11/100)^2 #Calculating BMI
var_label(datawt$bmi) <- "Body mass index" 


#Had heart attack or stroke
datawt$cvdcase <- datawt$h17 
datawt$cvdcase[datawt$cvdcase==2] <-0
datawt$cvdcase <- factor(datawt$cvdcase,
                     levels = c(0,1),
                     labels = c("No", "Yes"))

var_label(datawt$cvdcase) <- "Had heart attack or stroke"


#On aspirin
datawt$aspirin <- datawt$h18 
datawt$aspirin[datawt$aspirin==2] <-0
datawt$aspirin <- factor(datawt$aspirin,
                         levels = c(0,1),
                         labels = c("No", "Yes"))

var_label(datawt$aspirin) <- "Taking aspirin"


#On statins
datawt$statins <- datawt$h19
datawt$statins[datawt$statins==2] <-0
datawt$statins <- factor(datawt$statins,
                         levels = c(0,1),
                         labels = c("No", "Yes"))
 
var_label(datawt$statins) <- "Taking statins"


var_label(datawt$country) <- "Country"


datawt$tc_policy <-factor(datawt$tc_policy,
                          levels = c(1,2,3),
                          labels = c("Low", "Middle","High"))

var_label(datawt$tc_policy) <- "Tobacco restriction"


datawt$alc_policy <-factor(datawt$restriction,
                           levels = c(1,2,3),
                           labels = c("Low", "Middle","High"))
  
var_label(datawt$alc_policy) <- "Alcohol restriction"

datawt$region <-factor(datawt$subregion,
                           levels = c("Eastern Africa","Middle Africa","Northern Africa","Southern Africa","Western Africa"),
                           labels = c("East", "Middle","North","South","West"))

var_label(datawt$region) <- "Region" 

var_label(datawt$agerange2) <- "Age range" 

var_label(datawt$age) <- "Age" 

dplyr::mutate(datawt, ID = row_number()) #Generate the autonumber of the rows

datawt$highrisk <- if_else(datawt$highrisk==2,0,1)
#datawt$alcharm <- if_else(datawt$alcharm==1,"Yes","No")
datawt$cholesterol <- datawt$b8 
datawt$cholesteroltx <- datawt$b9 

datawt$highriskdes <- datawt$c 
datawt$cvdtherapy <- datawt$d 
datawt$cholbelo6p2 <- datawt$f

#Select the variables to be used in the analysis  
cvdafro <-datawt %>%
  dplyr::mutate(datawt, ID = row_number())  %>% #Generate the autonumber of the rows  
  select(c(ID,psu,statins,aspirin,cvdcase,country,gender,age,age_cat,dbm,bmi,residence,mstatus,edlev,occupation,bp,lvi,highsalt,lfi,smokehis,
           psu,alcharm,highriskdes,cvdtherapy,cholbelo6p2,cholesterol,cholesteroltx,highrisk,tc_policy,alc_policy,ptotal,agerange2,mets1200,pa,work,trans,rec,survyear,stratum,wstep1,wstep2,wstep3,m4a,m5a,m6a)) 

remove(MISSING,datawt_names,missing_names,set_author,tobacco_policy_africa,set_title,alcohol_policy_africa,amerge,tmerge,country_STEPS_surveys_years)

# #Creating age groups
datawt <- datawt %>% 
  mutate(
    age_cat = age_categories(             # create new column
      age,                            # numeric column to make groups from
      breakers = c(15,30,40,50, 60)))

var_label(datawt$age_cat) <- "Age groups" 

c <- 

#BMI categories
# datawt$bmi[is.na(datawt$bmi)] <-23
datawt <- datawt %>%  
  mutate(
    bmi_cat = age_categories(             # create new column
      bmi,                            # numeric column to make groups from
      breakers = c(0,18.5,25.0,30.0)))

var_label(datawt$bmi_cat) <- "BMI categories" 


save(datawt,file = "cvd_risk_africa_2024.RData")

save(cvdafro,file = "datawt.RData")


datawt <- writexl::write_xlsx(cvdafro,"C:\\Users\\LENOVO\\Documents\\HappyGitwithR\\steps_ssa_data\\CVD_Analysis_2024\\Data\\datawt.xlsx",col_names = TRUE, format_headers = TRUE,)
#datawt <-read_excel("datawt.xlsx")

cvd_risk_africa_2024 <- writexl::write_xlsx(cvdafro,"C:\\Users\\LENOVO\\Documents\\HappyGitwithR\\steps_ssa_data\\CVD_Analysis_2024\\Data\\cvd_risk_africa_2024.xlsx",col_names = TRUE, format_headers = TRUE,)
#cvd_risk_africa_2024 <-read_excel("cvd_risk_africa_2024.xlsx")
```
