## Supplementary material for "The Prevalence, Prevention, and Treatment of Cardiovascular Diseases in Twelve African Countries (2014-2019): An Analysis of the World Health Organisation STEPwise Approach to Chronic Disease Risk Factor Surveillance": Codes: 2_who_steps_ncd_data_cleaning_code_2024.R

```
library(writexl)
library(readxl)

#Load functions
rm(list = ls(all = TRUE)) #a\ Remove all data in memory
cvd_africa_2024 <- read_excel("data/cvd_africa_2024_clean.xlsx")
#
cvd_africa_2024$idcode <-100
cvd_africa_2024$idcode[cvd_africa_2024$country=="Benin"] <-200
cvd_africa_2024$idcode[cvd_africa_2024$country=="Botswana"] <-300
cvd_africa_2024$idcode[cvd_africa_2024$country=="Eswatini"] <-400
cvd_africa_2024$idcode[cvd_africa_2024$country=="Ethiopia"] <-500
cvd_africa_2024$idcode[cvd_africa_2024$country=="Kenya"] <-600
cvd_africa_2024$idcode[cvd_africa_2024$country=="Malawi"] <-700
cvd_africa_2024$idcode[cvd_africa_2024$country=="Morocco"] <-800
cvd_africa_2024$idcode[cvd_africa_2024$country=="STP"] <-900
cvd_africa_2024$idcode[cvd_africa_2024$country=="Uganda"] <-1000
cvd_africa_2024$idcode[cvd_africa_2024$country=="Zambia"] <-1100
cvd_africa_2024$idcode[cvd_africa_2024$country=="Sudan"] <-1200
#
#
cvd_africa_2024$alcharm <- factor(cvd_africa_2024$alcharm)
set.seed(123) # #  Set seed for reproducibility
#
#Proportions for each category
proportions_alcharm <- c("No" = 0.97, "Yes" = 0.03)
#
#Get the indices of NA values in cvd_africa_2024$alcharm
na_indices_alcharm <- which(is.na(cvd_africa_2024$alcharm))
#
#Number of NA values
n_na_alcharm <- length(na_indices_alcharm)
#
#Sample replacement values based on the given proportions
replacement_values_alcharm <- sample(
  names(proportions_alcharm),
  size = n_na_alcharm,
  replace = TRUE,
  prob = proportions_alcharm
)
#
#Replace the NA values with the sampled values
cvd_africa_2024$alcharm[na_indices_alcharm] <- replacement_values_alcharm
#
#Check the updated column
table(cvd_africa_2024$alcharm)
#
#
var_label(cvd_africa_2024$alcharm) <- "Harmful alcohol use"
#
#Smoking history
set.seed(123) # #  Set seed for reproducibility
#
#Proportions for each education level
proportions <- c("Current" = 0.1, "Never" = 0.83, "Previous" = 0.07)
#
#Get the indices of NA values in cvd_africa_2024$edlev
na_indices <- which(is.na(cvd_africa_2024$smokehis))
#
#Number of NA values
n_na <- length(na_indices)
#
#Sample replacement values based on the given proportions
replacement_values <- sample(
  names(proportions),         #Possible values ("None", "Primary", etc.)
  size = n_na,                #Number of values to sample
  replace = TRUE,             #Sampling with replacement
  prob = proportions          #The given probabilities
)
#
#Replace the NA values with the sampled values
cvd_africa_2024$smokehis[na_indices] <- replacement_values
#
#Check the updated column
table(cvd_africa_2024$smokehis)
cvd_africa_2024$smokehis <- factor(cvd_africa_2024$smokehis)
var_label(cvd_africa_2024$smokehis) <- "Smoking history"
#
#
#Define the distribution probabilities
set.seed(123) # #  Set seed for reproducibility
pa_probs <- c("1) Low Level" = 0.1643, "2) Moderate level" = 0.1970, "3) High level" = 0.6387)
#
#Check which values of pa are missing
missing_pa_indices <- which(is.na(cvd_africa_2024$pa))
#
#Simulate missing values based on the distribution
set.seed(123)   #Setting seed for reproducibility
#Simulate values for the missing data
cvd_africa_2024$pa[missing_pa_indices] <- sample(
  c("1) Low Level", "2) Moderate level", "3) High level"),
  length(missing_pa_indices),
  replace = TRUE,
  prob = pa_probs
)
#
#Check the results
table(cvd_africa_2024$pa, useNA = "ifany")
#


#
#Sex
set.seed(123) # #  Set seed for reproducibility
#
#Proportions for gender
gender_proportions <- c("Female" = 0.49, "Male" = 0.51)
#
#Get the indices of NA values in cvd_africa_2024$gender
na_gender_indices <- which(is.na(cvd_africa_2024$gender))
#
#Number of NA values
n_na_gender <- length(na_gender_indices)
#
#Sample replacement values based on the given proportions
gender_replacements <- sample(
  names(gender_proportions),   #Possible values ("Female", "Male")
  size = n_na_gender,          #Number of values to sample
  replace = TRUE,              #Sampling with replacement
  prob = gender_proportions    #The given probabilities
)
#
#Replace the NA values with the sampled values
cvd_africa_2024$gender[na_gender_indices] <- gender_replacements
#Check the updated gender column
table(cvd_africa_2024$gender)
#
cvd_africa_2024$gender <- factor(cvd_africa_2024$gender)
#
var_label(cvd_africa_2024$gender) <- "Gender"
#
#Age group
cvd_africa_2024$age_cat <- factor(cvd_africa_2024$age_cat)
var_label(cvd_africa_2024$age_cat) <- "Age groups"
#
#High salt intake
cvd_africa_2024$highsalt <- factor(cvd_africa_2024$highsalt)
#
var_label(cvd_africa_2024$highsalt) <- "High salt intake"
#
#
#Low fruit intake
cvd_africa_2024$lfi <- factor(cvd_africa_2024$lfi)
#
var_label(cvd_africa_2024$lfi) <- "Low fruit intake"
#
#Low vegetable uptake
cvd_africa_2024$lvi <- factor(cvd_africa_2024$lvi)
#
var_label(cvd_africa_2024$lvi) <- "Low vegetable intake"
#
#High blood pressure
cvd_africa_2024$bp <- factor(cvd_africa_2024$bp)
#
var_label(cvd_africa_2024$bp) <- "Has high BP"
#
#High blood sugar
cvd_africa_2024$dbm <- factor(cvd_africa_2024$dbm)
#
var_label(cvd_africa_2024$dbm) <- "High blood sugar"
#
#Occupation
cvd_africa_2024$occupation <- factor(cvd_africa_2024$occupation)
#
var_label(cvd_africa_2024$occupation) <- "Occupation"
#
#Highest education level
cvd_africa_2024$edlev <- factor(cvd_africa_2024$edlev)
#
var_label(cvd_africa_2024$edlev) <- "Occupation"
#
#Marital status
cvd_africa_2024$mstatus <- factor(cvd_africa_2024$mstatus)
#
var_label(cvd_africa_2024$mstatus) <- "Marital status"
#
#Area of residence
cvd_africa_2024$residence <- factor(cvd_africa_2024$residence)
#
var_label(cvd_africa_2024$residence) <- "Area of residence"
#
#Body mass index
var_label(cvd_africa_2024$bmi) <- "Body mass index"
#
#Had heart attack or stroke
cvd_africa_2024$cvdcase <- factor(cvd_africa_2024$cvdcase)
#
cvd_africa_2024$cvd <- if_else(cvd_africa_2024$cvdcase=="Yes",1,0)
#cvd_africa_2024$cvd <- factor(cvd_africa_2024$cvd)
#
var_label(cvd_africa_2024$cvdcase) <- "Had heart attack or stroke"
#
#On aspirin
cvd_africa_2024$aspirin <- factor(cvd_africa_2024$aspirin)
#
var_label(cvd_africa_2024$aspirin) <- "Taking aspirin"
#
#On statins
cvd_africa_2024$statins <- factor(cvd_africa_2024$statins)
#
var_label(cvd_africa_2024$statins) <- "Taking statins"
#
cvd_africa_2024$country <- factor(cvd_africa_2024$country)
#
var_label(cvd_africa_2024$country) <- "Country"
#Replace missing values in statins/aspirin with "No"
cvd_africa_2024$statins[is.na(cvd_africa_2024$statins)] <- "No"
cvd_africa_2024$aspirin[is.na(cvd_africa_2024$aspirin)] <- "No"
#
#Simulating cholesterol levels
##  Set seed for reproducibility
set.seed(123)
# Define the mean and#  Standard deviation
mean_cholesterol <- 2.827
sd_cholesterol <- 1.502
# Find the indices of NA values in cvd_africa_2024$cholesterol
na_indices_cholesterol <- which(is.na(cvd_africa_2024$cholesterol))
#
#Number of NA values
n_na_cholesterol <- length(na_indices_cholesterol)
#Generate random values from the normal distribution
replacement_cholesterol_values <- rnorm(n_na_cholesterol, mean = mean_cholesterol, sd = sd_cholesterol)
#Replace the NA values with the generated values
cvd_africa_2024$cholesterol[na_indices_cholesterol] <- replacement_cholesterol_values
# Replace cholesterol values with their absolute values
cvd_africa_2024$cholesterol <- abs(cvd_africa_2024$cholesterol)
# Check the updated cholesterol column
summary(cvd_africa_2024$cholesterol)
#
#
var_label(cvd_africa_2024$age) <- "Age"
cvd_africa_2024$pa <-factor(cvd_africa_2024$pa)
var_label(cvd_africa_2024$pa) <- "Physical activity"
cvd_africa_2024$work <-factor(cvd_africa_2024$work)
var_label(cvd_africa_2024$work) <- "Does work activity"
cvd_africa_2024$trans <-factor(cvd_africa_2024$trans)
var_label(cvd_africa_2024$trans) <- "Does transport activity"
cvd_africa_2024$rec <-factor(cvd_africa_2024$rec)
var_label(cvd_africa_2024$rec) <- "Does recreation activity"
#
#
#Creating age groups
set.seed(123)
cvd_africa_2024$age[is.na(cvd_africa_2024$age)] <- rnorm(
  sum(is.na(cvd_africa_2024$age)),  #Number of NA values
  mean = 37.93,                     # Mean value
  sd = 13.52                       #  Standard deviation
)
 cvd_africa_2024 <- cvd_africa_2024 %>%
   mutate(
     age_cat = age_categories(             #  create new column
       age,                            #  numeric column to make groups from
       breakers = c(15,30,40,50, 60)))
 var_label(cvd_africa_2024$age_cat) <- "Age groups"
#
 #BMI categories
set.seed(123)
cvd_africa_2024$bmi[is.na(cvd_africa_2024$bmi)] <- rnorm(
  sum(is.na(cvd_africa_2024$bmi)),  #Number of NA values
  mean = 24.47,                     # Mean value
  sd = 11.88                       #  Standard deviation
)
#
 cvd_africa_2024 <- cvd_africa_2024 %>%
   mutate(
    bmi_cat = age_categories(             #  create new column
       bmi,                            #  numeric column to make groups from
       breakers = c(0,18.5,25.0,30.0)))
#
 var_label(cvd_africa_2024$bmi_cat) <- "BMI categories"
#
var_label(cvd_africa_2024$survyear) <- "Survey year"
#
#
#BMI Category
#  Set seed for reproducibility
set.seed(123)
#Proportions for each BMI category
bmi_cat_proportions <- c("0-17.5" = 0.1551,
                         "18.5-24" = 0.5315,
                         "25-29" = 0.1740,
                         "30+" = 0.1395)
# Get the indices of NA values in cvd_africa_2024$bmi_cat
na_indices_bmi <- which(is.na(cvd_africa_2024$bmi_cat))
#
#Number of NA values
n_na_bmi <- length(na_indices_bmi)
#
#Sample replacement values based on the given proportions
replacement_bmi_values <- sample(
  names(bmi_cat_proportions),   #  Possible BMI categories
  size = n_na_bmi,              #Number of values to sample
  replace = TRUE,               #Sampling with replacement
  prob = bmi_cat_proportions    #The given probabilities
)
#
#Replace the NA values with the sampled values
cvd_africa_2024$bmi_cat[na_indices_bmi] <- replacement_bmi_values
# Check the updated bmi_cat column
table(cvd_africa_2024$bmi_cat)
#
#
#Highest education level
set.seed(123) # #  Set seed for reproducibility
#
#Proportions for each education level
proportions <- c("None" = 0.29, "Primary" = 0.35, "Secondary" = 0.26, "Tertiary" = 0.10)
#
#Get the indices of NA values in cvd_africa_2024$edlev
na_indices <- which(is.na(cvd_africa_2024$edlev))
#
#Number of NA values
n_na <- length(na_indices)
#
#Sample replacement values based on the given proportions
replacement_values <- sample(
  names(proportions),         #Possible values ("None", "Primary", etc.)
  size = n_na,                #Number of values to sample
  replace = TRUE,             #Sampling with replacement
  prob = proportions          #The given probabilities
)
#
#Replace the NA values with the sampled values
cvd_africa_2024$edlev[na_indices] <- replacement_values
#
#Check the updated column
table(cvd_africa_2024$edlev)
#
#
#Occupation
#Sample probabilities for each category
probs <- c(0.08, 0.09, 0.38, 0.44)
categories <- c("Government employee", "Non-Govt employee", "Non-paid/Retired", "Self-employed")
# Function to randomly sample a category
sample_category <- function() {
  return(sample(categories, 1, prob = probs))
}
#
cvd_africa_2024 <- mutate(cvd_africa_2024,
                           occupation = replace(occupation, is.na(occupation), sample_category()))
#
#Marital status
#  Set seed for reproducibility
set.seed(123)
#Proportions for "Currently married", "Formerly married", and "Never married"
proportions <- c("Currently married" = 0.68, "Formerly married" = 0.05, "Never married" = 0.26)
# Get the indices of NA values in cvd_africa_2024$mstatus
na_indices <- which(is.na(cvd_africa_2024$mstatus))
#
#Number of NA values
n_na <- length(na_indices)
#
#Sample replacement values based on the given proportions
replacement_values <- sample(
  names(proportions),         #Possible values ("Currently married", "Formerly married", "Never married")
  size = n_na,                #Number of values to sample
  replace = TRUE,             #Sampling with replacement
  prob = proportions          #The given probabilities
)
#
#Replace the NA values with the sampled values
cvd_africa_2024$mstatus[na_indices] <- replacement_values
#
#Check the updated column
table(cvd_africa_2024$mstatus)
#
#
#Diabetes
set.seed(123) # #  Set seed for reproducibility
#
# Proportions for "No" and "Yes"
proportions <- c("No" = 0.97, "Yes" = 0.03)
# Get the indices of NA values in cvd_africa_2024$dbm
na_indices <- which(is.na(cvd_africa_2024$dbm))
#
#Number of NA values
n_na <- length(na_indices)
#
#Sample replacement values based on the given proportions
replacement_values <- sample(
  names(proportions),         #Possible values ("No", "Yes")
  size = n_na,                #Number of values to sample
  replace = TRUE,             #Sampling with replacement
  prob = proportions          #The given probabilities
)
#
#Replace the NA values with the sampled values
cvd_africa_2024$dbm[na_indices] <- replacement_values
#
#Check the updated column
table(cvd_africa_2024$dbm)
#
#High salt uptake
set.seed(123) # #  Set seed for reproducibility
## Proportions for "No" and "Yes"
proportions_highsalt <- c("No" = 0.67, "Yes" = 0.33)
#Get the indices of NA values in cvd_africa_2024$highsalt
na_indices_highsalt <- which(is.na(cvd_africa_2024$highsalt))
#
#Number of NA values
n_na_highsalt <- length(na_indices_highsalt)
#
#Sample replacement values based on the given proportions
replacement_highsalt <- sample(
  names(proportions_highsalt),  #Possible values ("No", "Yes")
  size = n_na_highsalt,         #Number of values to sample
  replace = TRUE,               #Sampling with replacement
  prob = proportions_highsalt   #The given probabilities
)
#
#Replace the NA values with the sampled values
cvd_africa_2024$highsalt[na_indices_highsalt] <- replacement_highsalt
#
#Check the updated column
table(cvd_africa_2024$highsalt)
#
#Low fruit uptake
set.seed(123) # #  Set seed for reproducibility
## Proportions for "No" and "Yes"
proportions_lfi <- c("No" = 0.19, "Yes" = 0.81)
# Get the indices of NA values in cvd_africa_2024$lfi
na_indices_lfi <- which(is.na(cvd_africa_2024$lfi))
#
#Number of NA values
n_na_lfi <- length(na_indices_lfi)
#
#Sample replacement values based on the given proportions
replacement_values_lfi <- sample(
  names(proportions_lfi),     #Possible values ("No", "Yes")
  size = n_na_lfi,            #Number of values to sample
  replace = TRUE,             #Sampling with replacement
  prob = proportions_lfi      #The given probabilities
)
#
#Replace the NA values with the sampled values
cvd_africa_2024$lfi[na_indices_lfi] <- replacement_values_lfi
#
#Check the updated column
table(cvd_africa_2024$lfi)
#
#Low vegetable uptake
set.seed(123) # #  Set seed for reproducibility
## Proportions for "No" and "Yes"
proportions <- c("No" = 0.46, "Yes" = 0.54)
# Get the indices of NA values in cvd_africa_2024$lvi
na_indices <- which(is.na(cvd_africa_2024$lvi))
#
#Number of NA values
n_na <- length(na_indices)
#
#Sample replacement values based on the given proportions
replacement_values <- sample(
  names(proportions),         #Possible values ("No", "Yes")
  size = n_na,                #Number of values to sample
  replace = TRUE,             #Sampling with replacement
  prob = proportions          #The given probabilities
)
#
#Replace the NA values with the sampled values
cvd_africa_2024$lvi[na_indices] <- replacement_values
#
#Check the updated column
table(cvd_africa_2024$lvi)
#
#
#Physical activity level
set.seed(123) # #  Set seed for reproducibility

#Proportions for each physical activity level
pa_proportions <- c("1) Low Level" = 0.16, "2) Moderate level" = 0.20, "3) High level" = 0.64)

#Get the indices of NA values in cvd_africa_2024$pa
na_indices_pa <- which(is.na(cvd_africa_2024$pa))

#Number of NA values
n_na_pa <- length(na_indices_pa)

#Sample replacement values based on the given proportions
replacement_pa_values <- sample(
  names(pa_proportions),      #Possible values ("Low Level", "Moderate Level", "High Level")
  size = n_na_pa,             #Number of values to sample
  replace = TRUE,             #Sampling with replacement
  prob = pa_proportions       #The given probabilities
)

#Replace the NA values with the sampled values
cvd_africa_2024$pa[na_indices_pa] <- replacement_pa_values

#Check the updated column
table(cvd_africa_2024$pa)


#
#
cvd_africa_2024$survyear <-factor(cvd_africa_2024$survyear)
#
#View the first few rows of the subset
head(cvd_africa_2024)
#Classify cholesterol levels based on WHO classification
cvd_africa_2024$cholesterol_class <- ifelse(cvd_africa_2024$cholesterol >= 5.0, "High", "Low")
#
# Check the classification
table(cvd_africa_2024$cholesterol_class)
#
cvd_africa_2024$cholesterol_class <- factor(cvd_africa_2024$cholesterol_class)
#


#Physical activity level
set.seed(123)  # Set seed for reproducibility

# Proportions for each physical activity level
pa_proportions <- c("1) Low Level" = 0.16, "2) Moderate level" = 0.20, "3) High level" = 0.64)

# Get the indices of NA values in cvd_africa_2024$pa
na_indices_pa <- which(is.na(cvd_africa_2024$pa))

# Number of NA values
n_na_pa <- length(na_indices_pa)

# Sample replacement values based on the given proportions
replacement_pa_values <- sample(
  names(pa_proportions),      # Possible values ("Low Level", "Moderate Level", "High Level")
  size = n_na_pa,             # Number of values to sample
  replace = TRUE,             # Sampling with replacement
  prob = pa_proportions       # The given probabilities
)

# Replace the NA values with the sampled values
cvd_africa_2024$pa[na_indices_pa] <- replacement_pa_values

# Check the updated column
table(cvd_africa_2024$pa)


cvd_africa_2024_trt <-cvd_africa_2024


cvd_africa_2024_trt$trtcvd <- 0
cvd_africa_2024_trt$trtcvd[cvd_africa_2024_trt$aspirin=="Yes" & cvd_africa_2024_trt$statins !="Yes"] <-1
cvd_africa_2024_trt$trtcvd[cvd_africa_2024_trt$aspirin !="Yes" & cvd_africa_2024_trt$statins=="Yes"] <-2
cvd_africa_2024_trt$trtcvd[cvd_africa_2024_trt$aspirin=="Yes" & cvd_africa_2024_trt$statins=="Yes"] <-3
cvd_africa_2024_trt$trtcvd[cvd_africa_2024_trt$aspirin=="No" & cvd_africa_2024_trt$statins=="No" & cvd_africa_2024_trt$cvdtherapy=="1) received drug therapy and counseling"] <-4
#
cvd_africa_2024_trt$treatment <- if_else(cvd_africa_2024_trt$trtcvd==0,0,1)
#
cvd_africa_2024_trt$trtcvd <- factor(cvd_africa_2024_trt$trtcvd,
                                                        levels = c(0,1,2,3,4),
                                                        labels = c("None", "Aspirin", "Statins","Both aspirin and statins","Counselling"))
#
var_label(cvd_africa_2024_trt$trtcvd) <- "Treatment status"
#
#
#Population risk (2)
 cvd_africa_2024_trt$risk <- ifelse(cvd_africa_2024_trt$age > 39, 1, 0)
#
cvd_africa_2024_trt$highrisk2 <- ifelse(cvd_africa_2024_trt$risk > cvd_africa_2024_trt$highrisk, cvd_africa_2024_trt$risk, cvd_africa_2024_trt$highrisk)
#
cvd_africa_2024_trt$highrisk <- cvd_africa_2024_trt$highrisk2
#
#Creating datasets for treatment and prevention analyses
#
cvd_africa_2024_prevention <- cvd_africa_2024_trt %>%
  filter(cvdcase =="No") %>%
  filter(highrisk==1)
#
cvd_africa_2024_prevention$trtcvd <- if_else(cvd_africa_2024_prevention$cvdtherapy=="1) received drug therapy and counseling",1,0)
#
cvd_africa_2024_prevention$treatment <-cvd_africa_2024_prevention$trtcvd

#cvd_africa_2024_trt <-cvd_africa_2024
cvd_africa_2024_trt$trtcvd <- 0
cvd_africa_2024_trt$trtcvd[cvd_africa_2024_trt$aspirin=="Yes" & cvd_africa_2024_trt$statins !="Yes"] <-1
cvd_africa_2024_trt$trtcvd[cvd_africa_2024_trt$aspirin !="Yes" & cvd_africa_2024_trt$statins=="Yes"] <-2
cvd_africa_2024_trt$trtcvd[cvd_africa_2024_trt$aspirin=="Yes" & cvd_africa_2024_trt$statins=="Yes"] <-3
cvd_africa_2024_trt$trtcvd[cvd_africa_2024_trt$aspirin=="No" & cvd_africa_2024_trt$statins=="No" & cvd_africa_2024_trt$cvdtherapy=="1) received drug therapy and counseling"] <-4

cvd_africa_2024_trt$treatment <- if_else(cvd_africa_2024_trt$trtcvd==0,0,1)

cvd_africa_2024_trt$trtcvd <- factor(cvd_africa_2024_trt$trtcvd,
                                     levels = c(0,1,2,3,4),
                                     labels = c("None", "Aspirin", "Statins","Both aspirin and statins","Counselling"))

var_label(cvd_africa_2024_trt$trtcvd) <- "Treatment status"


#
cvd_africa_2024_treatment <- cvd_africa_2024_trt %>%
  filter(cvdcase =="Yes")
#
#
#
cvd_africa_2024_prevention$cholesterol_class <- factor(cvd_africa_2024_prevention$cholesterol_class)
#
cvd_africa_2024_treatment$cholesterol_class <- factor(cvd_africa_2024_treatment$cholesterol_class)
#
#Select a few variables for the analysis
cvd_africa_2024 <- cvd_africa_2024 %>%
  dplyr::select(gender, age_cat, age, bmi, residence, edlev, occupation, mstatus, dbm,
         bp, highsalt, lfi, lvi, pa, cholesterol_class,smokehis, alcharm, bmi_cat, survyear,
         cvdcase, aspirin, statins, country, highrisk, idcode, cvd, psu, stratum, wstep1,cholesterol)
#Calculate the mean of wstep1, ignoring missing values (NA)
mean_wstep1 <- mean(cvd_africa_2024$wstep1, na.rm = TRUE)
#Replace NA values in wstep1 with the calculated mean
cvd_africa_2024$wstep1[is.na(cvd_africa_2024$wstep1)] <- mean_wstep1


cvd_africa_2024_prevention <- cvd_africa_2024_trt %>%
  filter(cvdcase =="No") %>%
  filter(highrisk==1)

cvd_africa_2024_prevention$trtcvd <- if_else(cvd_africa_2024_prevention$cvdtherapy=="1) received drug therapy and counseling",1,0)

cvd_africa_2024_prevention$treatment <-cvd_africa_2024_prevention$trtcvd

cvd_africa_2024_treatment <- cvd_africa_2024_trt %>%
  filter(cvdcase =="Yes")

# Calculate the mean of wstep1, ignoring missing values (NA)
mean_wstep1 <- mean(cvd_africa_2024_treatment$wstep1, na.rm = TRUE)

# Replace NA values in wstep1 with the calculated mean
cvd_africa_2024_treatment$wstep1[is.na(cvd_africa_2024_treatment$wstep1)] <- mean_wstep1


# cvd_africa_2024_prevention$cholesterol_class <- factor(cvd_africa_2024_prevention$cholesterol_class)
#
# cvd_africa_2024_treatment$cholesterol_class <- factor(cvd_africa_2024_treatment$cholesterol_class)

#Select a few variables for the analysis
cvd_africa_2024 <- cvd_africa_2024 %>%
  dplyr::select(gender, age_cat, age, bmi, residence, edlev, occupation, mstatus, dbm,
         bp, highsalt, lfi, lvi, pa, cholesterol_class,smokehis, alcharm, bmi_cat, survyear,
         cvdcase, aspirin, statins, country, highrisk, idcode, cvd, psu, stratum, wstep1,cholesterol)

# Calculate the mean of wstep1, ignoring missing values (NA)
mean_wstep1 <- mean(cvd_africa_2024$wstep1, na.rm = TRUE)

# Replace NA values in wstep1 with the calculated mean
cvd_africa_2024$wstep1[is.na(cvd_africa_2024$wstep1)] <- mean_wstep1

# Check the updated column
summary(cvd_africa_2024$wstep1)

myvars = c("age_cat","bmi_cat", "gender", "residence","cvdtherapy", "edlev", "bp", "highsalt", "lfi", "smokehis","dbm","lvi","occupation","survyear","pa","mstatus","country","alcharm","trtcvd", "treatment","psu","stratum","wstep1","cholesterol_class")
cvd_africa_2024_prevention <- cvd_africa_2024_prevention[myvars]


myvars = c("age_cat","bmi_cat", "gender", "residence","cvdtherapy", "edlev", "bp", "highsalt", "lfi", "smokehis","dbm","lvi","occupation","trtcvd","survyear","pa","mstatus","country","alcharm","treatment", "psu","stratum","wstep1","cholesterol_class")
cvd_africa_2024_treatment <- cvd_africa_2024_treatment[myvars]


#Check the updated column
summary(cvd_africa_2024$wstep1)
#
myvars = c("age_cat","bmi_cat", "gender", "residence", "edlev", "bp", "highsalt", "lfi", "smokehis","dbm","lvi","occupation","survyear","pa","mstatus","country","alcharm","trtcvd", "treatment","psu","stratum","wstep1","cholesterol_class")
cvd_africa_2024_prevention <- cvd_africa_2024_prevention[myvars]
#
#
myvars = c("age_cat","bmi_cat", "gender", "residence", "edlev", "bp", "highsalt", "lfi", "smokehis","dbm","lvi","occupation","trtcvd","survyear","pa","mstatus","country","alcharm","treatment", "psu","stratum","wstep1","cholesterol_class")
cvd_africa_2024_treatment <- cvd_africa_2024_treatment[myvars]

#Save R objects created
save(cvd_africa_2024,file="cvd_africa_2024.RData")
save(cvd_africa_2024_prevention,file="cvd_africa_2024_prevention.RData")
save(cvd_africa_2024_treatment,file="cvd_africa_2024_treatment.RData")
 
```
