## Supplementary material for "The Prevalence, Prevention, and Treatment of Cardiovascular Diseases in Twelve African Countries (2014-2019): An Analysis of the World Health Organisation STEPwise Approach to Chronic Disease Risk Factor Surveillance": Codes: 3_CVD_Analysis_SSA_2024_FINAL.Rmd

````
---
title: "The prevalence, prevention and treatment of cardiovascular Diseases amongst persons aged 40-69 years in twelve African countries between 2014 and 2019: An analysis of the WHO STEPwise Survey"
author: "Wingston Ng'ambi" 
date: "2024-05-24"
output: 
  pdf_document: default
  word_document: default
  html_document: default 
---

```{r setup, include=FALSE}
knitr::opts_chunk$set(echo = TRUE)
```


Setting the workin directory 
```{r, eval=TRUE, tidy = TRUE, size="small", cache=FALSE, results="asis", message=F, warning=F}
setwd("C:\\Users\\LENOVO\\Documents\\GitHub\\CVD_Prevalence_Prevention_Treatment_Paper")
```

Loading packages
```{r, eval=TRUE, tidy = TRUE, size="small", cache=FALSE, results="asis", message=F, warning=F}
library(haven)
library(dplyr)
library(tidyverse)
library(labelled)
library(epikit)
library(sjPlot)
library(gtsummary)
library(writexl)
library(readxl)
library(mice) 
library(magrittr)
library(visdat)
library(caret)
library(caTools)
library(kableExtra)
library(srvyr) 
library(survey) 
library(weights)
library(finalfit)
library(MASS)
library(here)
options(survey.lonely.psu = "adjust")
options(survey.adjust.domain.lonely = TRUE)
source("Functions_rmph.R")

``` 


Load data
```{r, eval=TRUE, tidy = TRUE, size="small", cache=FALSE, results="asis", message=F, warning=F}

options(digits=4) 

#Load saved datafiles
load(here("C:\\Users\\LENOVO\\Documents\\GitHub\\CVD_Prevalence_Prevention_Treatment_Paper\\cvd_africa_2024.RData"))
load(here("C:\\Users\\LENOVO\\Documents\\GitHub\\CVD_Prevalence_Prevention_Treatment_Paper\\cvd_africa_2024_prevention.RData"))
load(here("C:\\Users\\LENOVO\\Documents\\GitHub\\CVD_Prevalence_Prevention_Treatment_Paper\\cvd_africa_2024_treatment.RData"))

#Setting data to approapriate survey designs 
STEPSClean <- cvd_africa_2024 %>% 
  as_survey_design(ids=psu, strata=stratum, weights=wstep1, nest=TRUE) 

STEPSCleanPrevention <- cvd_africa_2024_prevention %>% 
  as_survey_design(ids=psu, strata=stratum, weights=wstep1, nest=TRUE)


STEPSCleanTreatment <- svydesign(ids = ~1, data = cvd_africa_2024_treatment,
                      weights = cvd_africa_2024_treatment$wstep1)
  
################################################################################

```

Characteristics of men and women included in the analysis
```{r, eval=TRUE, tidy = TRUE, size="small", cache=FALSE, results="asis", message=F, warning=F}
#Age
##Age group
#For absolute numbers
ag <-cvd_africa_2024 %>% 
  group_by(age_cat) %>% 
  tally()
  
kable(ag)
#For weighted percentages
kable(wpct(cvd_africa_2024$age_cat, weight=cvd_africa_2024$wstep1, na.rm=FALSE)*100)

#Summarize age as a continuous variable
# Calculate weighted percentiles
weighted_percentiles <- wtd.quantile(cvd_africa_2024$age, 
                                     weights = cvd_africa_2024$wstep1, 
                                     probs = c(0.25, 0.5, 0.75))
weighted_percentiles

#Sex
#For absolute numbers
gen <-cvd_africa_2024 %>% 
  group_by(gender) %>% 
  tally()
  
kable(gen)
#For weighted percentages
kable(wpct(cvd_africa_2024$gender, weight=cvd_africa_2024$wstep1, na.rm=FALSE)*100)

#Residence
#For absolute numbers
resid <-cvd_africa_2024 %>% 
  group_by(residence) %>% 
  tally()
  
kable(resid)
#For weighted percentages
kable(wpct(cvd_africa_2024$residence, weight=cvd_africa_2024$wstep1, na.rm=FALSE)*100)

#Highest education level
#For absolute numbers
edl <-cvd_africa_2024 %>% 
  group_by(edlev) %>% 
  tally()
  
kable(edl)
#For weighted percentages
kable(wpct(cvd_africa_2024$edlev, weight=cvd_africa_2024$wstep1, na.rm=FALSE)*100)


#Cholesterol level
#For absolute numbers
chol <-cvd_africa_2024 %>% 
  group_by(cholesterol_class) %>% 
  tally()
  
kable(chol)
#For weighted percentages
kable(wpct(cvd_africa_2024$cholesterol_class, weight=cvd_africa_2024$wstep1, na.rm=FALSE)*100)

weighted_percentiles <- wtd.quantile(cvd_africa_2024$cholesterol, 
                                     weights = cvd_africa_2024$wstep1, 
                                     probs = c(0.25, 0.5, 0.75))
weighted_percentiles


#Occupation
#For absolute numbers
occ <-cvd_africa_2024 %>% 
  group_by(occupation) %>% 
  tally()
  
kable(occ)
#For weighted percentages
kable(wpct(cvd_africa_2024$occupation, weight=cvd_africa_2024$wstep1, na.rm=FALSE)*100)

#Marital status
#For absolute numbers
marital <-cvd_africa_2024 %>% 
  group_by(mstatus) %>% 
  tally()
  
kable(marital)
#For weighted percentages
kable(wpct(cvd_africa_2024$mstatus, weight=cvd_africa_2024$wstep1, na.rm=FALSE)*100)

#Country
#For absolute numbers
countr <-cvd_africa_2024 %>% 
  group_by(country) %>% 
  tally()
  
kable(countr)
#For weighted percentages
kable(wpct(cvd_africa_2024$country, weight=cvd_africa_2024$wstep1, na.rm=FALSE)*100)

#Body mass index
summary(cvd_africa_2024$bmi)

#Diabetes
#For absolute numbers
db <-cvd_africa_2024 %>% 
  group_by(dbm) %>% 
  tally()
  
kable(db)
#For weighted percentages
kable(wpct(cvd_africa_2024$dbm, weight=cvd_africa_2024$wstep1, na.rm=FALSE)*100)

#Hypertension
#For absolute numbers
htn <-cvd_africa_2024 %>% 
  group_by(bp) %>% 
  tally()
  
kable(htn)
#For weighted percentages
kable(wpct(cvd_africa_2024$bp, weight=cvd_africa_2024$wstep1, na.rm=FALSE)*100)

#High salt uptake
#For absolute numbers
salt <-cvd_africa_2024 %>% 
  group_by(highsalt) %>% 
  tally()
  
kable(salt)
#For weighted percentages
kable(wpct(cvd_africa_2024$highsalt, weight=cvd_africa_2024$wstep1, na.rm=FALSE)*100)

#Low fruit uptake
#For absolute numbers
fruit <-cvd_africa_2024 %>% 
  group_by(lfi) %>% 
  tally()
  
kable(fruit)
#For weighted percentages
kable(wpct(cvd_africa_2024$lfi, weight=cvd_africa_2024$wstep1, na.rm=FALSE)*100)

#Low vegetable uptake
#For absolute numbers
veg <-cvd_africa_2024 %>% 
  group_by(lvi) %>% 
  tally()
  
kable(veg)
#For weighted percentages
kable(wpct(cvd_africa_2024$lvi, weight=cvd_africa_2024$wstep1, na.rm=FALSE)*100)

#Physical activity
#For absolute numbers
physical <-cvd_africa_2024 %>% 
  group_by(pa) %>% 
  tally()
  
kable(physical)
#For weighted percentages
kable(wpct(cvd_africa_2024$pa, weight=cvd_africa_2024$wstep1, na.rm=FALSE)*100)

#Tobacco use
#For absolute numbers
tobacco <-cvd_africa_2024 %>% 
  group_by(smokehis) %>% 
  tally()
  
kable(tobacco)
#For weighted percentages
kable(wpct(cvd_africa_2024$smokehis, weight=cvd_africa_2024$wstep1, na.rm=FALSE)*100)

#Harmful alcohol use
#For absolute numbers
alcohol <-cvd_africa_2024 %>% 
  group_by(alcharm) %>% 
  tally()
  
kable(alcohol)
#For weighted percentages
kable(wpct(cvd_africa_2024$alcharm, weight=cvd_africa_2024$wstep1, na.rm=FALSE)*100)

#BMI
#For absolute numbers
bmic <-cvd_africa_2024 %>% 
  group_by(bmi_cat) %>% 
  tally()
  
kable(bmic)
#For weighted percentages
kable(wpct(cvd_africa_2024$bmi_cat, weight=cvd_africa_2024$wstep1, na.rm=F)*100)

# Calculate weighted percentiles
weighted_percentiles <- wtd.quantile(cvd_africa_2024$bmi, 
                                     weights = cvd_africa_2024$wstep1, 
                                     probs = c(0.25, 0.5, 0.75))
weighted_percentiles

#Survey year
#For absolute numbers
alcohol <-cvd_africa_2024 %>% 
  group_by(survyear) %>% 
  tally()
  
kable(alcohol) 
#For weighted percentages
kable(wpct(cvd_africa_2024$survyear, weight=cvd_africa_2024$wstep1, na.rm=FALSE)*100)

```


Characteristics of men and women included in the prevention analysis
```{r, eval=TRUE, tidy = TRUE, size="small", cache=FALSE, results="asis", message=F, warning=F}
#Age
##Age group
#For absolute numbers
ag <-cvd_africa_2024_prevention %>% 
  group_by(age_cat) %>% 
  tally()

kable(ag)
#For weighted percentages
kable(wpct(cvd_africa_2024_prevention$age_cat, weight=cvd_africa_2024_prevention$wstep1, na.rm=FALSE)*100)

#Summarize age as a continuous variable
summary(cvd_africa_2024_prevention$age)

# age_summ <- cvd_africa_2024_prevention %>%
#   summarize(Age = survey_mean(age,
#     vartype = c("se", "ci")
#   ))
# 
# 
# age_median <- cvd_africa_2024_prevention %>%
#   summarize( survey_quantile(age,
#     quantiles = c(0.25, .5, 0.75)
#   ))
#Sex
#For absolute numbers
gen <-cvd_africa_2024_prevention %>% 
  group_by(gender) %>% 
  tally()

kable(gen)
#For weighted percentages
kable(wpct(cvd_africa_2024_prevention$gender, weight=cvd_africa_2024_prevention$wstep1, na.rm=FALSE)*100)

#Residence
#For absolute numbers
resid <-cvd_africa_2024_prevention %>% 
  group_by(residence) %>% 
  tally()

kable(resid)
#For weighted percentages
kable(wpct(cvd_africa_2024_prevention$residence, weight=cvd_africa_2024_prevention$wstep1, na.rm=FALSE)*100)

#Highest education level
#For absolute numbers
edl <-cvd_africa_2024_prevention %>% 
  group_by(edlev) %>% 
  tally()

kable(edl)
#For weighted percentages
kable(wpct(cvd_africa_2024_prevention$edlev, weight=cvd_africa_2024_prevention$wstep1, na.rm=FALSE)*100)

#Cholesterol level
#For absolute numbers
chol <-cvd_africa_2024_prevention %>% 
  group_by(cholesterol_class) %>% 
  tally()
  
kable(chol)
#For weighted percentages
kable(wpct(cvd_africa_2024_prevention$cholesterol_class, weight=cvd_africa_2024_prevention$wstep1, na.rm=FALSE)*100)


#Occupation
#For absolute numbers
occ <-cvd_africa_2024_prevention %>% 
  group_by(occupation) %>% 
  tally()

kable(occ)
#For weighted percentages
kable(wpct(cvd_africa_2024_prevention$occupation, weight=cvd_africa_2024_prevention$wstep1, na.rm=FALSE)*100)

#Marital status
#For absolute numbers
marital <-cvd_africa_2024_prevention %>% 
  group_by(mstatus) %>% 
  tally()

kable(marital)
#For weighted percentages
kable(wpct(cvd_africa_2024_prevention$mstatus, weight=cvd_africa_2024_prevention$wstep1, na.rm=FALSE)*100)

#(1+idcode)
#For absolute numbers
countr <-cvd_africa_2024_prevention %>% 
  group_by(country) %>% 
  tally()

kable(countr)
#For weighted percentages
kable(wpct(cvd_africa_2024_prevention$country, weight=cvd_africa_2024_prevention$wstep1, na.rm=FALSE)*100)

#Body mass index
summary(cvd_africa_2024_prevention$bmi)

#Diabetes
#For absolute numbers
db <-cvd_africa_2024_prevention %>% 
  group_by(dbm) %>% 
  tally()

kable(db)
#For weighted percentages
kable(wpct(cvd_africa_2024_prevention$dbm, weight=cvd_africa_2024_prevention$wstep1, na.rm=FALSE)*100)

#Hypertension
#For absolute numbers
htn <-cvd_africa_2024_prevention %>% 
  group_by(bp) %>% 
  tally()

kable(htn)
#For weighted percentages
kable(wpct(cvd_africa_2024_prevention$bp, weight=cvd_africa_2024_prevention$wstep1, na.rm=FALSE)*100)

#High salt uptake
#For absolute numbers
salt <-cvd_africa_2024_prevention %>% 
  group_by(highsalt) %>% 
  tally()

kable(salt)
#For weighted percentages
kable(wpct(cvd_africa_2024_prevention$highsalt, weight=cvd_africa_2024_prevention$wstep1, na.rm=FALSE)*100)

#Low fruit uptake
#For absolute numbers
fruit <-cvd_africa_2024_prevention %>% 
  group_by(lfi) %>% 
  tally()

kable(fruit)
#For weighted percentages
kable(wpct(cvd_africa_2024_prevention$lfi, weight=cvd_africa_2024_prevention$wstep1, na.rm=FALSE)*100)

#Low vegetable uptake
#For absolute numbers
veg <-cvd_africa_2024_prevention %>% 
  group_by(lvi) %>% 
  tally()

kable(veg)
#For weighted percentages
kable(wpct(cvd_africa_2024_prevention$lvi, weight=cvd_africa_2024_prevention$wstep1, na.rm=FALSE)*100)

#Physical activity
#For absolute numbers
physical <-cvd_africa_2024_prevention %>% 
  group_by(pa) %>% 
  tally()

kable(physical)
#For weighted percentages
kable(wpct(cvd_africa_2024_prevention$pa, weight=cvd_africa_2024_prevention$wstep1, na.rm=FALSE)*100)

#Tobacco use
#For absolute numbers
tobacco <-cvd_africa_2024_prevention %>% 
  group_by(smokehis) %>% 
  tally()

kable(tobacco)
#For weighted percentages
kable(wpct(cvd_africa_2024_prevention$smokehis, weight=cvd_africa_2024_prevention$wstep1, na.rm=FALSE)*100)

#Harmful alcohol use
#For absolute numbers
alcohol <-cvd_africa_2024_prevention %>% 
  group_by(alcharm) %>% 
  tally()

kable(alcohol)
#For weighted percentages
kable(wpct(cvd_africa_2024_prevention$alcharm, weight=cvd_africa_2024_prevention$wstep1, na.rm=FALSE)*100)

#BMI
#For absolute numbers
bmic <-cvd_africa_2024_prevention %>% 
  group_by(bmi_cat) %>% 
  tally()

kable(bmic)
#For weighted percentages
kable(wpct(cvd_africa_2024_prevention$bmi_cat, weight=cvd_africa_2024_prevention$wstep1, na.rm=F)*100)

#Survey year
#For absolute numbers
alcohol <-cvd_africa_2024_prevention %>% 
  group_by(survyear) %>% 
  tally()

kable(alcohol)
#For weighted percentages
kable(wpct(cvd_africa_2024_prevention$survyear, weight=cvd_africa_2024_prevention$wstep1, na.rm=FALSE)*100)

```

Tabulating the outcomes
Prevalence of CVD
```{r, eval=TRUE, tidy = TRUE, size="small", cache=FALSE, results="asis", message=F, warning=F}
#For absolute numbers
cases <-cvd_africa_2024 %>% 
  group_by(cvdcase) %>% 
  tally()
  
kable(cases)
#For weighted percentages
kable(wpct(cvd_africa_2024$cvdcase, weight=cvd_africa_2024$wgt, na.rm=FALSE)*100)
```

Uptake of prevention for CVD
```{r, eval=TRUE, tidy = TRUE, size="small", cache=FALSE, results="asis", message=F, warning=F}
#For absolute numbers
prev <-cvd_africa_2024_prevention %>% 
  group_by(trtcvd) %>%  
  tally()
  
kable(prev)
#For weighted percentages
kable(wpct(cvd_africa_2024_prevention$trtcvd, weight=cvd_africa_2024_prevention$wgt, na.rm=FALSE)*100)
```


Uptake of statins for treatment of CVD
```{r, eval=TRUE, tidy = TRUE, size="small", cache=FALSE, results="asis", message=F, warning=F}
#For absolute numbers
trt <-cvd_africa_2024_treatment %>% 
  group_by(trtcvd) %>% 
  tally()
   
kable(trt)
#For weighted percentages
kable(wpct(cvd_africa_2024_treatment$trtcvd, weight=cvd_africa_2024_treatment$wgt, na.rm=T)*100)
```


Bivariate analysis of prevalence of CVD

```{r, eval=TRUE, tidy = TRUE, size="small", cache=FALSE, results="asis", message=F, warning=F}
#Age
age_x <- cvd_africa_2024 %>%
  group_by(age_cat, cvdcase)%>%
  dplyr::summarize(n=n())%>%
  spread(cvdcase, n) 
kable(age_x)
 
kable(svyby(formula=~I(cvdcase=="Yes"),by=~age_cat, 
      design=STEPSClean,
      FUN=svyciprop,vartype="ci",method="beta")*100)

#Sex
age_x <- cvd_africa_2024 %>%
  group_by(gender, cvdcase)%>%
  dplyr::summarize(n=n())%>%
  spread(cvdcase, n) 
kable(age_x)
 
kable(svyby(formula=~I(cvdcase=="Yes"),by=~gender, 
      design=STEPSClean,
      FUN=svyciprop,vartype="ci",method="beta")*100)

#Residnence
age_x <- cvd_africa_2024 %>%
  group_by(residence, cvdcase)%>%
  dplyr::summarize(n=n())%>%
  spread(cvdcase, n) 
kable(age_x)
 
kable(svyby(formula=~I(cvdcase=="Yes"),by=~residence, 
      design=STEPSClean,
      FUN=svyciprop,vartype="ci",method="beta")*100)

#Highest education level
ed_x <- cvd_africa_2024 %>%
  group_by(edlev, cvdcase)%>%
  dplyr::summarize(n=n())%>%
  spread(cvdcase, n) 
kable(ed_x)
 
kable(svyby(formula=~I(cvdcase=="Yes"),by=~edlev, 
      design=STEPSClean,
      FUN=svyciprop,vartype="ci",method="beta")*100)

#Cholesterol level
chol_x <- cvd_africa_2024 %>%
  group_by(cholesterol_class, cvdcase)%>%
  dplyr::summarize(n=n())%>%
  spread(cvdcase, n) 
kable(chol_x)
 
kable(svyby(formula=~I(cvdcase=="Yes"),by=~cholesterol_class, 
      design=STEPSClean,
      FUN=svyciprop,vartype="ci",method="beta")*100)

#Occupation
occ_x <- cvd_africa_2024 %>%
  group_by(occupation, cvdcase)%>%
  dplyr::summarize(n=n())%>%
  spread(cvdcase, n) 
kable(occ_x)
 
kable(svyby(formula=~I(cvdcase=="Yes"),by=~occupation, 
      design=STEPSClean,
      FUN=svyciprop,vartype="ci",method="beta")*100)

#Marital status
mst_x <- cvd_africa_2024 %>%
  group_by(mstatus, cvdcase)%>%
  dplyr::summarize(n=n())%>%
  spread(cvdcase, n) 
kable(mst_x)
 
kable(svyby(formula=~I(cvdcase=="Yes"),by=~mstatus, 
      design=STEPSClean,
      FUN=svyciprop,vartype="ci",method="beta")*100)

#country
mst_x <- cvd_africa_2024 %>%
  group_by(country, cvdcase)%>%
  dplyr::summarize(n=n())%>%
  spread(cvdcase, n) 
kable(mst_x)
 
kable(svyby(formula=~I(cvdcase=="Yes"),by=~country, 
     design=STEPSClean,
     FUN=svyciprop,vartype="ci",method="beta")*100)

#Body mass index

#Diabetes
mst_x <- cvd_africa_2024 %>%
  group_by(dbm, cvdcase)%>%
  dplyr::summarize(n=n())%>% 
  spread(cvdcase, n) 
kable(mst_x)
 
kable(svyby(formula=~I(cvdcase=="Yes"),by=~dbm, 
      design=STEPSClean,
      FUN=svyciprop,vartype="ci",method="beta")*100)

#Hypertension
mst_x <- cvd_africa_2024 %>%
  group_by(bp, cvdcase)%>%
  dplyr::summarize(n=n())%>% 
  spread(cvdcase, n) 
kable(mst_x)
 
kable(svyby(formula=~I(cvdcase=="Yes"),by=~bp, 
      design=STEPSClean,
      FUN=svyciprop,vartype="ci",method="beta")*100)

#High salt uptake
mst_x <- cvd_africa_2024 %>%
  group_by(highsalt, cvdcase)%>%
  dplyr::summarize(n=n())%>% 
  spread(cvdcase, n) 
kable(mst_x)
 
kable(svyby(formula=~I(cvdcase=="Yes"),by=~highsalt, 
      design=STEPSClean,
      FUN=svyciprop,vartype="ci",method="beta")*100)

#Low fruit uptake
mst_x <- cvd_africa_2024 %>%
  group_by(lfi, cvdcase)%>%
  dplyr::summarize(n=n())%>% 
  spread(cvdcase, n) 
kable(mst_x)
 
kable(svyby(formula=~I(cvdcase=="Yes"),by=~lfi, 
      design=STEPSClean,
      FUN=svyciprop,vartype="ci",method="beta")*100)


#Low vegetable uptake
mst_x <- cvd_africa_2024 %>%
  group_by(lvi, cvdcase)%>%
  dplyr::summarize(n=n())%>% 
  spread(cvdcase, n) 
kable(mst_x)
 
kable(svyby(formula=~I(cvdcase=="Yes"),by=~lvi, 
      design=STEPSClean,
      FUN=svyciprop,vartype="ci",method="beta")*100)


#Physical activity
mst_x <- cvd_africa_2024 %>%
  group_by(pa, cvdcase)%>%
  dplyr::summarize(n=n())%>% 
  spread(cvdcase, n) 
kable(mst_x)
 
kable(svyby(formula=~I(cvdcase=="Yes"),by=~pa, 
      design=STEPSClean,
      FUN=svyciprop,vartype="ci",method="beta")*100)

#Tobacco use
mst_x <- cvd_africa_2024 %>%
  group_by(smokehis, cvdcase)%>%
  dplyr::summarize(n=n())%>% 
  spread(cvdcase, n) 
kable(mst_x)
 
kable(svyby(formula=~I(cvdcase=="Yes"),by=~smokehis, 
      design=STEPSClean,
      FUN=svyciprop,vartype="ci",method="beta")*100)

#Harmful alcohol use
mst_x <- cvd_africa_2024 %>%
  group_by(alcharm, cvdcase)%>%
  dplyr::summarize(n=n())%>% 
  spread(cvdcase, n) 
kable(mst_x)
 
kable(svyby(formula=~I(cvdcase=="Yes"),by=~alcharm, 
      design=STEPSClean,
      FUN=svyciprop,vartype="ci",method="beta")*100)

#BMI
mst_x <- cvd_africa_2024 %>%
  group_by(bmi_cat, cvdcase)%>%
  dplyr::summarize(n=n())%>% 
  spread(cvdcase, n) 
kable(mst_x)
 
kable(svyby(formula=~I(cvdcase=="Yes"),by=~bmi_cat, 
      design=STEPSClean,
      FUN=svyciprop,vartype="ci",method="beta")*100)

#Survey year
mst_x <- cvd_africa_2024 %>%
  group_by(survyear, cvdcase)%>%
  dplyr::summarize(n=n())%>% 
  spread(cvdcase, n) 
kable(mst_x)
 
kable(svyby(formula=~I(cvdcase=="Yes"),by=~survyear, 
      design=STEPSClean,
      FUN=svyciprop,vartype="ci",method="beta")*100)

```

Bivariate analysis for uptake of aspirin and statins for prevention of CVD 

```{r, eval=TRUE, tidy = TRUE, size="small", cache=FALSE, results="asis", message=F, warning=F}
#Age
age_x <- cvd_africa_2024_prevention %>%
  group_by(age_cat, treatment)%>%
  dplyr::summarize(n=n())%>%
  spread(treatment, n)
kable(age_x)

kable(svyby(formula=~I(treatment==1),by=~age_cat,
      design=STEPSCleanPrevention,
      FUN=svyciprop,vartype="ci",method="beta")*100)


#Gender
age_x <- cvd_africa_2024_prevention %>%
  group_by(gender, treatment)%>%
  dplyr::summarize(n=n())%>%
  spread(treatment, n)
kable(age_x)

kable(svyby(formula=~I(treatment==1),by=~gender,
      design=STEPSCleanPrevention,
      FUN=svyciprop,vartype="ci",method="beta")*100)

#Residence
age_x <- cvd_africa_2024_prevention %>%
  group_by(residence, treatment)%>%
  dplyr::summarize(n=n())%>%
  spread(treatment, n)
kable(age_x)

kable(svyby(formula=~I(treatment==1),by=~residence,
      design=STEPSCleanPrevention,
      FUN=svyciprop,vartype="ci",method="beta")*100)

#Highest education level
ed_x <- cvd_africa_2024_prevention %>%
  group_by(edlev, treatment)%>%
  dplyr::summarize(n=n())%>%
  spread(treatment, n)
kable(ed_x)

kable(svyby(formula=~I(treatment==1),by=~edlev,
      design=STEPSCleanPrevention,
      FUN=svyciprop,vartype="ci",method="beta")*100)

#Cholesterol level
chol_x <- cvd_africa_2024_prevention %>%
  group_by(cholesterol_class, treatment)%>%
  dplyr::summarize(n=n())%>%
  spread(treatment, n) 
kable(chol_x)
 
kable(svyby(formula=~I(treatment==1),by=~cholesterol_class, 
      design=STEPSCleanPrevention,
      FUN=svyciprop,vartype="ci",method="beta")*100)

#Occupation
occ_x <- cvd_africa_2024_prevention %>%
  group_by(occupation, treatment)%>%
  dplyr::summarize(n=n())%>%
  spread(treatment, n)
kable(occ_x)

kable(svyby(formula=~I(treatment==1),by=~occupation,
      design=STEPSCleanPrevention,
      FUN=svyciprop,vartype="ci",method="beta")*100)

#Marital status
mst_x <- cvd_africa_2024_prevention %>%
  group_by(mstatus, treatment)%>%
  dplyr::summarize(n=n())%>%
  spread(treatment, n)
kable(mst_x)

kable(svyby(formula=~I(treatment==1),by=~mstatus,
      design=STEPSCleanPrevention,
      FUN=svyciprop,vartype="ci",method="beta")*100)

#Country
mst_x <- cvd_africa_2024_prevention %>%
  group_by(country, treatment)%>%
  dplyr::summarize(n=n())%>%
  spread(treatment, n)
kable(mst_x)

kable(svyby(formula=~I(treatment==1),by=~(country),
     design=STEPSCleanPrevention,
     FUN=svyciprop,vartype="ci",method="beta")*100)


#Diabetes
mst_x <- cvd_africa_2024_prevention %>%
  group_by(dbm, treatment)%>%
  dplyr::summarize(n=n())%>%
  spread(treatment, n)
kable(mst_x)

kable(svyby(formula=~I(treatment==1),by=~dbm,
      design=STEPSCleanPrevention,
      FUN=svyciprop,vartype="ci",method="beta")*100)

#Hypertension
mst_x <- cvd_africa_2024_prevention %>%
  group_by(bp, treatment)%>%
  dplyr::summarize(n=n())%>%
  spread(treatment, n)
kable(mst_x)

kable(svyby(formula=~I(treatment==1),by=~bp,
      design=STEPSCleanPrevention,
      FUN=svyciprop,vartype="ci",method="beta")*100)

#High salt uptake
mst_x <- cvd_africa_2024_prevention %>%
  group_by(highsalt, treatment)%>%
  dplyr::summarize(n=n())%>%
  spread(treatment, n)
kable(mst_x)

kable(svyby(formula=~I(treatment==1),by=~highsalt,
      design=STEPSCleanPrevention,
      FUN=svyciprop,vartype="ci",method="beta")*100)


#Low fruit uptake
mst_x <- cvd_africa_2024_prevention %>%
  group_by(lfi, treatment)%>%
  dplyr::summarize(n=n())%>%
  spread(treatment, n)
kable(mst_x)

kable(svyby(formula=~I(treatment==1),by=~lfi,
      design=STEPSCleanPrevention,
      FUN=svyciprop,vartype="ci",method="beta")*100)

#Low vegetable uptake
mst_x <- cvd_africa_2024_prevention %>%
  group_by(lvi, treatment)%>%
  dplyr::summarize(n=n())%>%
  spread(treatment, n)
kable(mst_x)

kable(svyby(formula=~I(treatment==1),by=~lvi,
      design=STEPSCleanPrevention,
      FUN=svyciprop,vartype="ci",method="beta")*100)

#Physical activity
mst_x <- cvd_africa_2024_prevention %>%
  group_by(pa, treatment)%>%
  dplyr::summarize(n=n())%>%
  spread(treatment, n)
kable(mst_x)

kable(svyby(formula=~I(treatment==1),by=~pa,
      design=STEPSCleanPrevention,
      FUN=svyciprop,vartype="ci",method="beta")*100)


#Tobacco use
mst_x <- cvd_africa_2024_prevention %>%
  group_by(smokehis, treatment)%>%
  dplyr::summarize(n=n())%>%
  spread(treatment, n)
kable(mst_x)

kable(svyby(formula=~I(treatment==1),by=~smokehis,
      design=STEPSCleanPrevention,
      FUN=svyciprop,vartype="ci",method="beta")*100)

#Harmful alcohol use
mst_x <- cvd_africa_2024_prevention %>%
  group_by(alcharm, treatment)%>%
  dplyr::summarize(n=n())%>%
  spread(treatment, n)
kable(mst_x)

kable(svyby(formula=~I(treatment==1),by=~alcharm,
      design=STEPSCleanPrevention,
      FUN=svyciprop,vartype="ci",method="beta")*100)

#BMI
mst_x <- cvd_africa_2024_prevention %>%
  group_by(bmi_cat, treatment)%>%
  dplyr::summarize(n=n())%>%
  spread(treatment, n)
kable(mst_x)

kable(svyby(formula=~I(treatment==1),by=~bmi_cat,
      design=STEPSCleanPrevention,
      FUN=svyciprop,vartype="ci",method="beta")*100)

#Survey year
mst_x <- cvd_africa_2024_prevention %>%
  group_by(survyear, treatment)%>%
  dplyr::summarize(n=n())%>%
  spread(treatment, n)
kable(mst_x)

kable(svyby(formula=~I(treatment==1),by=~survyear,
      design=STEPSCleanPrevention,
      FUN=svyciprop,vartype="ci",method="beta")*100)

```

Bivariate analysis for uptake of aspirin and statins for treatment of CVD

```{r, eval=TRUE, tidy = TRUE, size="small", cache=FALSE, results="asis", message=F, warning=F}
#Age
age_x <- cvd_africa_2024_treatment %>%
  group_by(age_cat, treatment)%>%
  dplyr::summarize(n=n())%>%
  spread(treatment, n) 
kable(age_x)
 
kable(svyby(formula=~I(treatment==1),by=~age_cat, 
     design=STEPSCleanTreatment,
      FUN=svyciprop,vartype="ci",method="beta",drop = T)*100)

#Gender
age_x <- cvd_africa_2024_treatment %>%
  group_by(gender, treatment)%>% 
  dplyr::summarize(n=n())%>%
  spread(treatment, n) 
kable(age_x)
 
kable(svyby(formula=~I(treatment==1),by=~gender, 
      design=STEPSCleanTreatment,
      FUN=svyciprop,vartype="ci",method="beta")*100)

#Residence
res_x <- cvd_africa_2024_treatment %>%
  group_by(residence, treatment)%>% 
  dplyr::summarize(n=n())%>%
  spread(treatment, n) 
kable(res_x)
 
kable(svyby(formula=~I(treatment==1),by=~residence, 
      design=STEPSCleanTreatment,
      FUN=svyciprop,vartype="ci",method="beta")*100)

#Highest education level
res_x <- cvd_africa_2024_treatment %>%
  group_by(edlev, treatment)%>% 
  dplyr::summarize(n=n())%>%
  spread(treatment, n) 
kable(res_x)
 
kable(svyby(formula=~I(treatment==1),by=~edlev, 
      design=STEPSCleanTreatment,
      FUN=svyciprop,vartype="ci",method="beta")*100)

#Cholesterol level
chol_x <- cvd_africa_2024_treatment %>%
  group_by(cholesterol_class, treatment)%>%
  dplyr::summarize(n=n())%>%
  spread(treatment, n) 
kable(chol_x)
 
kable(svyby(formula=~I(treatment==1),by=~cholesterol_class, 
      design=STEPSCleanTreatment,
      FUN=svyciprop,vartype="ci",method="beta")*100)

#Occupation
occ_x <- cvd_africa_2024_treatment %>%
  group_by(occupation, treatment)%>% 
  dplyr::summarize(n=n())%>%
  spread(treatment, n) 
kable(occ_x)
 
kable(svyby(formula=~I(treatment==1),by=~occupation, 
      design=STEPSCleanTreatment,
      FUN=svyciprop,vartype="ci",method="beta")*100)

#Marital status
mst_x <- cvd_africa_2024_treatment %>%
  group_by(mstatus, treatment)%>%
  dplyr::summarize(n=n())%>%
  spread(treatment, n) 
kable(mst_x)
 
kable(svyby(formula=~I(treatment==1),by=~mstatus, 
      design=STEPSCleanTreatment,
      FUN=svyciprop,vartype="ci",method="beta")*100)

#country
mst_x <- cvd_africa_2024_treatment %>%
  group_by(country, treatment)%>%
  dplyr::summarize(n=n())%>%
  spread(treatment, n)  
kable(mst_x)
 
kable(svyby(formula=~I(treatment==1),by=~country,
    design=STEPSCleanTreatment,
    FUN=svyciprop,vartype="ci",method="beta")*100)


#Diabetes
mst_x <- cvd_africa_2024_treatment %>%
  group_by(dbm, treatment)%>%
  dplyr::summarize(n=n())%>%
  spread(treatment, n) 
kable(mst_x)
 
kable(svyby(formula=~I(treatment==1),by=~dbm, 
      design=STEPSCleanTreatment,
      FUN=svyciprop,vartype="ci",method="beta")*100)

#Hypertension
mst_x <- cvd_africa_2024_treatment %>%
  group_by(bp, treatment)%>%
  dplyr::summarize(n=n())%>% 
  spread(treatment, n) 
kable(mst_x)
 
kable(svyby(formula=~I(treatment==1),by=~bp, 
      design=STEPSCleanTreatment,
      FUN=svyciprop,vartype="ci",method="beta")*100)

#High salt uptake
mst_x <- cvd_africa_2024_treatment %>%
  group_by(highsalt, treatment)%>%
  dplyr::summarize(n=n())%>% 
  spread(treatment, n) 
kable(mst_x)
 
kable(svyby(formula=~I(treatment==1),by=~highsalt, 
      design=STEPSCleanTreatment,
      FUN=svyciprop,vartype="ci",method="beta")*100)

#Low fruit uptake
mst_x <- cvd_africa_2024_treatment %>%
  group_by(lfi, treatment)%>%
  dplyr::summarize(n=n())%>% 
  spread(treatment, n) 
kable(mst_x)
 
kable(svyby(formula=~I(treatment==1),by=~lfi,  
      design=STEPSCleanTreatment,
      FUN=svyciprop,vartype="ci",method="beta")*100)

#Low vegetable uptake
mst_x <- cvd_africa_2024_treatment %>%
  group_by(lvi, treatment)%>%
  dplyr::summarize(n=n())%>% 
  spread(treatment, n) 
kable(mst_x)
 
kable(svyby(formula=~I(treatment==1),by=~lvi, 
      design=STEPSCleanTreatment,
      FUN=svyciprop,vartype="ci",method="beta")*100)


#Physical activity
mst_x <- cvd_africa_2024_treatment %>%
  group_by(pa, treatment)%>%
  dplyr::summarize(n=n())%>% 
  spread(treatment, n) 
kable(mst_x) 
 
kable(svyby(formula=~I(treatment==1),by=~pa, 
      design=STEPSCleanTreatment,
      FUN=svyciprop,vartype="ci",method="beta")*100)

#Tobacco use
mst_x <- cvd_africa_2024_treatment %>%
  group_by(smokehis, treatment)%>%
  dplyr::summarize(n=n())%>% 
  spread(treatment, n) 
kable(mst_x)
 
kable(svyby(formula=~I(treatment==1),by=~smokehis, 
      design=STEPSCleanTreatment,
      FUN=svyciprop,vartype="ci",method="beta")*100)

#Harmful alcohol use
mst_x <- cvd_africa_2024_treatment %>%
  group_by(alcharm, treatment)%>%
  dplyr::summarize(n=n())%>% 
  spread(treatment, n) 
kable(mst_x)
 
kable(svyby(formula=~I(treatment==1),by=~alcharm, 
      design=STEPSCleanTreatment,
      FUN=svyciprop,vartype="ci",method="beta")*100) 

#BMI
mst_x <- cvd_africa_2024_treatment %>%
  group_by(bmi_cat, treatment)%>%
  dplyr::summarize(n=n())%>% 
  spread(treatment, n) 
kable(mst_x)
 
kable(svyby(formula=~I(treatment==1),by=~bmi_cat, 
      design=STEPSCleanTreatment,
      FUN=svyciprop,vartype="ci",method="beta")*100) 

#Survey year
mst_x <- cvd_africa_2024_treatment %>%
  group_by(survyear, treatment)%>%
  dplyr::summarize(n=n())%>% 
  spread(treatment, n) 
kable(mst_x)
 
kable(svyby(formula=~I(treatment==1),by=~survyear, 
      design=STEPSCleanTreatment,
      FUN=svyciprop,vartype="ci",method="beta")*100) 


```

Univariate regression
1.Prevalence of CVD

```{r, eval=TRUE, tidy = TRUE, size="small", cache=FALSE, results="asis", message=F, warning=F}
mod_empty <- svyglm(cvd ~ 1, family = quasibinomial(), design = STEPSClean)

#Country
modempty <- svyglm(cvd ~ country, family = quasibinomial(), design =STEPSClean)
round(
  cbind(
    summary(modempty,
            df.resid=degf(modempty$survey.design))$coef,
    confint(modempty, 
            ddf=degf(modempty$survey.design))
  )
  , 4)

# Compute odds ratios and their 95% confidence intervals
OR.CI <- cbind("AOR" = exp(   coef(modempty)),
               exp(confint(modempty,
                           df.resid=degf(modempty$survey.design))))[-1,]
kable(round(OR.CI, 3))

# Perform the F test
anova_result <- anova(mod_empty, modempty, test = "F")

# Display the result
anova_result


#Age
modage <- svyglm(cvd ~ age_cat , family=quasibinomial(), design =STEPSClean)
round(
  cbind( 
    summary(modage,
            df.resid=degf(modage$survey.design))$coef,
    confint(modage,
            ddf=degf(modage$survey.design))
  )
  , 4)
#Compute odds ratios and their 95% confidence intervals
OR.CI <- cbind("AOR" = exp(   coef(modage)),
               exp(confint(modage,
                           df.resid=degf(modage$survey.design))))[-1,]
kable(round(OR.CI, 3))

# Perform the F test
anova_result <- anova(mod_empty, modage, test = "F")

# Display the result
anova_result
#Sex
modsex <- svyglm(cvd ~ gender , family = quasibinomial(), design =STEPSClean)
# Perform the F test
anova_result <- anova(mod_empty, modsex, test = "F")

# Display the result
anova_result
round(
  cbind(
    summary(modsex,
            df.resid=degf(modsex$survey.design))$coef,
    confint(modsex,
            ddf=degf(modsex$survey.design))
  )
  , 4)

# Compute odds ratios and their 95% confidence intervals
OR.CI <- cbind("AOR" = exp(   coef(modsex)),
               exp(confint(modsex,
                           df.resid=degf(modsex$survey.design))))[-1,]
kable(round(OR.CI, 3))

#Residence
modres <- svyglm(cvd ~ residence , family = quasibinomial(), design =STEPSClean)
anova_result <- anova(mod_empty, modres, test = "F")

# Display the result
anova_result
round(
  cbind(
    summary(modres,
            df.resid=degf(modres$survey.design))$coef,
    confint(modres,
            ddf=degf(modres$survey.design))
  )
  , 4)

# Compute odds ratios and their 95% confidence intervals
OR.CI <- cbind("AOR" = exp(   coef(modres)),
               exp(confint(modres,
                           df.resid=degf(modres$survey.design))))[-1,]
kable(round(OR.CI, 3))

#Highest education level
modedl <- svyglm(cvd ~ edlev , family = quasibinomial(), design =STEPSClean)
anova_result <- anova(mod_empty, modedl, test = "F")

# Display the result
anova_result
round(
  cbind(
    summary(modedl,
            df.resid=degf(modedl$survey.design))$coef,
    confint(modedl,
            ddf=degf(modedl$survey.design))
  )
  , 4)

# Compute odds ratios and their 95% confidence intervals
OR.CI <- cbind("AOR" = exp(   coef(modedl)),
               exp(confint(modedl,
                           df.resid=degf(modedl$survey.design))))[-1,]
kable(round(OR.CI, 3))

#Cholesterol level
modedl <- svyglm(cvd ~ cholesterol_class , family = quasibinomial(), design =STEPSClean)
anova_result <- anova(mod_empty, modedl, test = "F")

# Display the result
anova_result
round(
  cbind(
    summary(modedl,
            df.resid=degf(modedl$survey.design))$coef,
    confint(modedl,
            ddf=degf(modedl$survey.design))
  )
  , 4)

# Compute odds ratios and their 95% confidence intervals
OR.CI <- cbind("AOR" = exp(   coef(modedl)),
               exp(confint(modedl,
                           df.resid=degf(modedl$survey.design))))[-1,]
kable(round(OR.CI, 3))


# Occupation
modocc <- svyglm(cvd ~ occupation , family = quasibinomial(), design =STEPSClean)
anova_result <- anova(mod_empty, modocc, test = "F")

# Display the result
anova_result
round(
  cbind(
    summary(modocc,
            df.resid=degf(modocc$survey.design))$coef,
    confint(modocc,
            ddf=degf(modocc$survey.design))
  )
  , 4)

# Compute odds ratios and their 95% confidence intervals
OR.CI <- cbind("AOR" = exp(   coef(modocc)),
               exp(confint(modocc,
                           df.resid=degf(modocc$survey.design))))[-1,]
kable(round(OR.CI, 3))


#Marital status
modmar <- svyglm(cvd ~ mstatus ,  family = quasibinomial(), design =STEPSClean)
anova_result <- anova(mod_empty, modmar, test = "F")

# Display the result
anova_result
round(
  cbind(
    summary(modmar,
            df.resid=degf(modmar$survey.design))$coef,
    confint(modmar,
            ddf=degf(modmar$survey.design))
  )
  , 4)

# Compute odds ratios and their 95% confidence intervals
OR.CI <- cbind("AOR" = exp(   coef(modmar)),
               exp(confint(modmar,
                           df.resid=degf(modmar$survey.design))))[-1,]
kable(round(OR.CI, 3))


# Diabetes
moddbm <- svyglm(cvd ~ dbm , family = quasibinomial(), design =STEPSClean)
anova_result <- anova(mod_empty, moddbm, test = "F")

# Display the result
anova_result
round(
  cbind(
    summary(moddbm,
            df.resid=degf(moddbm$survey.design))$coef,
    confint(moddbm,
            ddf=degf(moddbm$survey.design))
  )
  , 4)

# Compute odds ratios and their 95% confidence intervals
OR.CI <- cbind("AOR" = exp(   coef(moddbm)),
               exp(confint(moddbm,
                           df.resid=degf(moddbm$survey.design))))[-1,]
kable(round(OR.CI, 3))


# Hypertension
modhtn <- svyglm(cvd ~ bp , family = quasibinomial(), design =STEPSClean)
anova_result <- anova(mod_empty, modhtn, test = "F")

# Display the result
anova_result
round(
  cbind(
    summary(modhtn,
            df.resid=degf(modhtn$survey.design))$coef,
    confint(modhtn,
            ddf=degf(modhtn$survey.design))
  )
  , 4)

# Compute odds ratios and their 95% confidence intervals
OR.CI <- cbind("AOR" = exp(   coef(modhtn)),
               exp(confint(modhtn,
                           df.resid=degf(modhtn$survey.design))))[-1,]
kable(round(OR.CI, 3))


# High salt uptake
modhsu <- svyglm(cvd ~ highsalt , family = quasibinomial(), design =STEPSClean)
anova_result <- anova(mod_empty, modhsu, test = "F")

# Display the result
anova_result
round(
  cbind(
    summary(modhsu,
            df.resid=degf(modhsu$survey.design))$coef,
    confint(modhsu,
            ddf=degf(modhsu$survey.design))
  )
  , 4)

# Compute odds ratios and their 95% confidence intervals
OR.CI <- cbind("AOR" = exp(   coef(modhsu)),
               exp(confint(modhsu,
                           df.resid=degf(modhsu$survey.design))))[-1,]
kable(round(OR.CI, 3))


# Low fruit uptake
modlfu <- svyglm(cvd ~ lfi , family = quasibinomial(), design =STEPSClean)
anova_result <- anova(mod_empty, modlfu, test = "F")

# Display the result
anova_result
round(
  cbind(
    summary(modlfu,
            df.resid=degf(modlfu$survey.design))$coef,
    confint(modlfu,
            ddf=degf(modlfu$survey.design))
  )
  , 4)

# Compute odds ratios and their 95% confidence intervals
OR.CI <- cbind("AOR" = exp(   coef(modlfu)),
               exp(confint(modlfu,
                           df.resid=degf(modlfu$survey.design))))[-1,]
kable(round(OR.CI, 3))


# Low vegetable uptake
modlvu <- svyglm(cvd ~ lvi , family = quasibinomial(), design =STEPSClean)
anova_result <- anova(mod_empty, modlvu, test = "F")

# Display the result
anova_result
round(
  cbind(
    summary(modlvu,
            df.resid=degf(modlvu$survey.design))$coef,
    confint(modlvu,
            ddf=degf(modlvu$survey.design))
  )
  , 4)

# Compute odds ratios and their 95% confidence intervals
OR.CI <- cbind("AOR" = exp(   coef(modlvu)),
               exp(confint(modlvu,
                           df.resid=degf(modlvu$survey.design))))[-1,]
kable(round(OR.CI, 3))


# Physical activity
modpa <- svyglm(cvd ~ pa , family = quasibinomial(), design =STEPSClean)
anova_result <- anova(mod_empty, modpa, test = "F")

# Display the result
anova_result
round(
  cbind(
    summary(modpa,
            df.resid=degf(modpa$survey.design))$coef,
    confint(modpa,
            ddf=degf(modpa$survey.design))
  )
  , 4)

# Compute odds ratios and their 95% confidence intervals
OR.CI <- cbind("AOR" = exp(   coef(modpa)),
               exp(confint(modpa,
                           df.resid=degf(modpa$survey.design))))[-1,]
kable(round(OR.CI, 3))


# Tobacco use
modtu <- svyglm(cvd ~ smokehis , family = quasibinomial(), design =STEPSClean)
anova_result <- anova(mod_empty, modtu, test = "F")

# Display the result
anova_result
round(
  cbind(
    summary(modtu,
            df.resid=degf(modtu$survey.design))$coef,
    confint(modtu,
            ddf=degf(modtu$survey.design))
  )
  , 4)

# Compute odds ratios and their 95% confidence intervals
OR.CI <- cbind("AOR" = exp(   coef(modtu)),
               exp(confint(modtu,
                           df.resid=degf(modtu$survey.design))))[-1,]
kable(round(OR.CI, 3))


# Harmful alcohol use
modhau <- svyglm(cvd ~ alcharm , family = quasibinomial(), design =STEPSClean)
anova_result <- anova(mod_empty, modhau, test = "F")

# Display the result
anova_result
round(
  cbind(
    summary(modhau,
            df.resid=degf(modhau$survey.design))$coef,
    confint(modhau,
            ddf=degf(modhau$survey.design))
  )
  , 4)

# Compute odds ratios and their 95% confidence intervals
OR.CI <- cbind("AOR" = exp(   coef(modhau)),
               exp(confint(modhau,
                           df.resid=degf(modhau$survey.design))))[-1,]
kable(round(OR.CI, 3))


# BMI
modhau <- svyglm(cvd ~ bmi_cat , family = quasibinomial(), design =STEPSClean)
anova_result <- anova(mod_empty, modhau, test = "F")

# Display the result
anova_result
round(
  cbind(
    summary(modhau,
            df.resid=degf(modhau$survey.design))$coef,
    confint(modhau,
            ddf=degf(modhau$survey.design))
  )
  , 4)

# Compute odds ratios and their 95% confidence intervals
OR.CI <- cbind("AOR" = exp(   coef(modhau)),
               exp(confint(modhau,
                           df.resid=degf(modhau$survey.design))))[-1,]
kable(round(OR.CI, 3))


# Survey year
modsyr <- svyglm(cvd ~ survyear , family = quasibinomial(), design =STEPSClean)
anova_result <- anova(mod_empty, modsyr, test = "F")

# Display the result
anova_result
round(
  cbind(
    summary(modsyr,
            df.resid=degf(modsyr$survey.design))$coef,
    confint(modsyr,
            ddf=degf(modsyr$survey.design))
  )
  , 4)

# Compute odds ratios and their 95% confidence intervals
OR.CI <- cbind("AOR" = exp(   coef(modsyr)),
               exp(confint(modsyr,
                           df.resid=degf(modsyr$survey.design))))[-1,]
kable(round(OR.CI, 3))


```


2. Uptake of prevention for CVD


```{r, eval=TRUE, tidy = TRUE, size="small", cache=FALSE, results="asis", message=F, warning=F}
mod_empty <- svyglm(trtcvd ~ 1, family = quasibinomial(), design = STEPSCleanPrevention)
#Empty model
modempty <- svyglm(trtcvd ~ country, family = quasibinomial(), design =STEPSCleanPrevention)
anova_result <- anova(mod_empty, modempty, test = "F")

# Display the result
anova_result
round(
  cbind(
    summary(modempty,
            df.resid=degf(modempty$survey.design))$coef,
    confint(modempty, 
            ddf=degf(modempty$survey.design))
  )
  , 4)

# Compute odds ratios and their 95% confidence intervals
OR.CI <- cbind("AOR" = exp(   coef(modempty)),
               exp(confint(modempty,
                           df.resid=degf(modempty$survey.design))))[-1,]
kable(round(OR.CI, 3))

#Age
modage <- svyglm(trtcvd ~ age_cat , family=quasibinomial(), design =STEPSCleanPrevention)
anova_result <- anova(mod_empty, modage, test = "F")

# Display the result
anova_result
round(
  cbind( 
    summary(modage,
            df.resid=degf(modage$survey.design))$coef,
    confint(modage,
            ddf=degf(modage$survey.design))
  )
  , 4)
#Compute odds ratios and their 95% confidence intervals
OR.CI <- cbind("AOR" = exp(   coef(modage)),
               exp(confint(modage,
                           df.resid=degf(modage$survey.design))))[-1,]
kable(round(OR.CI, 3))


#Sex
modsex <- svyglm(trtcvd ~ gender , family = quasibinomial(), design =STEPSCleanPrevention)
anova_result <- anova(mod_empty, modsex, test = "F")

# Display the result
anova_result
round(
  cbind(
    summary(modsex,
            df.resid=degf(modsex$survey.design))$coef,
    confint(modsex,
            ddf=degf(modsex$survey.design))
  )
  , 4)

# Compute odds ratios and their 95% confidence intervals
OR.CI <- cbind("AOR" = exp(   coef(modsex)),
               exp(confint(modsex,
                           df.resid=degf(modsex$survey.design))))[-1,]
kable(round(OR.CI, 3))

#Residence
modres <- svyglm(trtcvd ~ residence , family = quasibinomial(), design =STEPSCleanPrevention)
anova_result <- anova(mod_empty, modres, test = "F")

# Display the result
anova_result
round(
  cbind(
    summary(modres,
            df.resid=degf(modres$survey.design))$coef,
    confint(modres,
            ddf=degf(modres$survey.design))
  )
  , 4)

# Compute odds ratios and their 95% confidence intervals
OR.CI <- cbind("AOR" = exp(   coef(modres)),
               exp(confint(modres,
                           df.resid=degf(modres$survey.design))))[-1,]
kable(round(OR.CI, 3))

#Highest education level
modedl <- svyglm(trtcvd ~ edlev , family = quasibinomial(), design =STEPSCleanPrevention)
anova_result <- anova(mod_empty, modedl, test = "F")

# Display the result
anova_result
round(
  cbind(
    summary(modedl,
            df.resid=degf(modedl$survey.design))$coef,
    confint(modedl,
            ddf=degf(modedl$survey.design))
  )
  , 4)

# Compute odds ratios and their 95% confidence intervals
OR.CI <- cbind("AOR" = exp(   coef(modedl)),
               exp(confint(modedl,
                           df.resid=degf(modedl$survey.design))))[-1,]
kable(round(OR.CI, 3))


#Cholesterol level
modedl <- svyglm(trtcvd ~ cholesterol_class , family = quasibinomial(), design =STEPSCleanPrevention)
anova_result <- anova(mod_empty, modedl, test = "F")

# Display the result
anova_result
round(
  cbind(
    summary(modedl,
            df.resid=degf(modedl$survey.design))$coef,
    confint(modedl,
            ddf=degf(modedl$survey.design))
  )
  , 4)

# Compute odds ratios and their 95% confidence intervals
OR.CI <- cbind("AOR" = exp(   coef(modedl)),
               exp(confint(modedl,
                           df.resid=degf(modedl$survey.design))))[-1,]
kable(round(OR.CI, 3))

# Occupation
modocc <- svyglm(trtcvd ~ occupation , family = quasibinomial(), design =STEPSCleanPrevention)
anova_result <- anova(mod_empty, modocc, test = "F")

# Display the result
anova_result
round(
  cbind(
    summary(modocc,
            df.resid=degf(modocc$survey.design))$coef,
    confint(modocc,
            ddf=degf(modocc$survey.design))
  )
  , 4)

# Compute odds ratios and their 95% confidence intervals
OR.CI <- cbind("AOR" = exp(   coef(modocc)),
               exp(confint(modocc,
                           df.resid=degf(modocc$survey.design))))[-1,]
kable(round(OR.CI, 3))


#Employment status
modmar <- svyglm(trtcvd ~ mstatus ,  family = quasibinomial(), design =STEPSCleanPrevention)
anova_result <- anova(mod_empty, modmar, test = "F")

# Display the result
anova_result
round(
  cbind(
    summary(modmar,
            df.resid=degf(modmar$survey.design))$coef,
    confint(modmar,
            ddf=degf(modmar$survey.design))
  )
  , 4)

# Compute odds ratios and their 95% confidence intervals
OR.CI <- cbind("AOR" = exp(   coef(modmar)),
               exp(confint(modmar,
                           df.resid=degf(modmar$survey.design))))[-1,]
kable(round(OR.CI, 3))


# Diabetes
moddbm <- svyglm(trtcvd ~ dbm , family = quasibinomial(), design =STEPSCleanPrevention)
anova_result <- anova(mod_empty, moddbm, test = "F")

# Display the result
anova_result
round(
  cbind(
    summary(moddbm,
            df.resid=degf(moddbm$survey.design))$coef,
    confint(moddbm,
            ddf=degf(moddbm$survey.design))
  )
  , 4)

# Compute odds ratios and their 95% confidence intervals
OR.CI <- cbind("AOR" = exp(   coef(moddbm)),
               exp(confint(moddbm,
                           df.resid=degf(moddbm$survey.design))))[-1,]
kable(round(OR.CI, 3))


# Hypertension
modhtn <- svyglm(trtcvd ~ bp , family = quasibinomial(), design =STEPSCleanPrevention)
anova_result <- anova(mod_empty, modhtn, test = "F")

# Display the result
anova_result
round(
  cbind(
    summary(modhtn,
            df.resid=degf(modhtn$survey.design))$coef,
    confint(modhtn,
            ddf=degf(modhtn$survey.design))
  )
  , 4)

# Compute odds ratios and their 95% confidence intervals
OR.CI <- cbind("AOR" = exp(   coef(modhtn)),
               exp(confint(modhtn,
                           df.resid=degf(modhtn$survey.design))))[-1,]
kable(round(OR.CI, 3))


# High salt uptake
modhsu <- svyglm(trtcvd ~ highsalt , family = quasibinomial(), design =STEPSCleanPrevention)
anova_result <- anova(mod_empty, modhsu, test = "F")

# Display the result
anova_result
round(
  cbind(
    summary(modhsu,
            df.resid=degf(modhsu$survey.design))$coef,
    confint(modhsu,
            ddf=degf(modhsu$survey.design))
  )
  , 4)

# Compute odds ratios and their 95% confidence intervals
OR.CI <- cbind("AOR" = exp(   coef(modhsu)),
               exp(confint(modhsu,
                           df.resid=degf(modhsu$survey.design))))[-1,]
kable(round(OR.CI, 3))


# Low fruit uptake
modlfu <- svyglm(trtcvd ~ lfi , family = quasibinomial(), design =STEPSCleanPrevention)
anova_result <- anova(mod_empty, modlfu, test = "F")

# Display the result
anova_result
round(
  cbind(
    summary(modlfu,
            df.resid=degf(modlfu$survey.design))$coef,
    confint(modlfu,
            ddf=degf(modlfu$survey.design))
  )
  , 4)

# Compute odds ratios and their 95% confidence intervals
OR.CI <- cbind("AOR" = exp(   coef(modlfu)),
               exp(confint(modlfu,
                           df.resid=degf(modlfu$survey.design))))[-1,]
kable(round(OR.CI, 3))


# Low vegetable uptake
modlvu <- svyglm(trtcvd ~ lvi , family = quasibinomial(), design =STEPSCleanPrevention)
anova_result <- anova(mod_empty, modlvu, test = "F")

# Display the result
anova_result
round(
  cbind(
    summary(modlvu,
            df.resid=degf(modlvu$survey.design))$coef,
    confint(modlvu,
            ddf=degf(modlvu$survey.design))
  )
  , 4)

# Compute odds ratios and their 95% confidence intervals
OR.CI <- cbind("AOR" = exp(   coef(modlvu)),
               exp(confint(modlvu,
                           df.resid=degf(modlvu$survey.design))))[-1,]
kable(round(OR.CI, 3))


# Physical activity
modpa <- svyglm(trtcvd ~ pa , family = quasibinomial(), design =STEPSCleanPrevention)
anova_result <- anova(mod_empty, modpa, test = "F")

# Display the result
anova_result
round(
  cbind(
    summary(modpa,
            df.resid=degf(modpa$survey.design))$coef,
    confint(modpa,
            ddf=degf(modpa$survey.design))
  )
  , 4)

# Compute odds ratios and their 95% confidence intervals
OR.CI <- cbind("AOR" = exp(   coef(modpa)),
               exp(confint(modpa,
                           df.resid=degf(modpa$survey.design))))[-1,]
kable(round(OR.CI, 3))


# Tobacco use
modtu <- svyglm(trtcvd ~ smokehis , family = quasibinomial(), design =STEPSCleanPrevention)
anova_result <- anova(mod_empty, modtu, test = "F")

# Display the result
anova_result
round(
  cbind(
    summary(modtu,
            df.resid=degf(modtu$survey.design))$coef,
    confint(modtu,
            ddf=degf(modtu$survey.design))
  )
  , 4)

# Compute odds ratios and their 95% confidence intervals
OR.CI <- cbind("AOR" = exp(   coef(modtu)),
               exp(confint(modtu,
                           df.resid=degf(modtu$survey.design))))[-1,]
kable(round(OR.CI, 3))


# Harmful alcohol use
modhau <- svyglm(trtcvd ~ alcharm , family = quasibinomial(), design =STEPSCleanPrevention)
anova_result <- anova(mod_empty, modhau, test = "F")

# Display the result
anova_result
round(
  cbind(
    summary(modhau,
            df.resid=degf(modhau$survey.design))$coef,
    confint(modhau,
            ddf=degf(modhau$survey.design))
  )
  , 4)

# Compute odds ratios and their 95% confidence intervals
OR.CI <- cbind("AOR" = exp(   coef(modhau)),
               exp(confint(modhau,
                           df.resid=degf(modhau$survey.design))))[-1,]
kable(round(OR.CI, 3))


# BMI
modhau <- svyglm(trtcvd ~ bmi_cat , family = quasibinomial(), design =STEPSCleanPrevention)
anova_result <- anova(mod_empty, modhau, test = "F")

# Display the result
anova_result
round(
  cbind(
    summary(modhau,
            df.resid=degf(modhau$survey.design))$coef,
    confint(modhau,
            ddf=degf(modhau$survey.design))
  )
  , 4)

# Compute odds ratios and their 95% confidence intervals
OR.CI <- cbind("AOR" = exp(   coef(modhau)),
               exp(confint(modhau,
                           df.resid=degf(modhau$survey.design))))[-1,]
kable(round(OR.CI, 3))


# Survey year
modsyr <- svyglm(trtcvd ~ survyear , family = quasibinomial(), design =STEPSCleanPrevention)
anova_result <- anova(mod_empty, modsyr, test = "F")

# Display the result
anova_result
round(
  cbind(
    summary(modsyr,
            df.resid=degf(modsyr$survey.design))$coef,
    confint(modsyr,
            ddf=degf(modsyr$survey.design))
  )
  , 4)

# Compute odds ratios and their 95% confidence intervals
OR.CI <- cbind("AOR" = exp(   coef(modsyr)),
               exp(confint(modsyr,
                           df.resid=degf(modsyr$survey.design))))[-1,]
kable(round(OR.CI, 3))

```
 
 

3. Uptake of treatment for CVD

```{r, eval=TRUE, tidy = TRUE, size="small", cache=FALSE, results="asis", message=F, warning=F}
mod_empty <- svyglm(treatment ~ 1, family = quasibinomial(), design = STEPSCleanTreatment)
#Empty model
modempty <- svyglm(treatment ~ country, family = quasibinomial(), design =STEPSCleanTreatment)
anova_result <- anova(mod_empty, modempty, test = "F")

# Display the result
anova_result
round(
  cbind(
    summary(modempty,
              df.resid=degf(modempty$survey.design))$coef,
    confint(modempty,
              ddf=degf(modempty$survey.design))
  )
, 4)

# Compute odds ratios and their 95% confidence intervals
OR.CI <- cbind("AOR" = exp(   coef(modempty)),
                       exp(confint(modempty,
                           df.resid=degf(modempty$survey.design))))[-1,]
kable(round(OR.CI, 3))


#Age
modage <- svyglm(treatment ~ age_cat , family = quasibinomial(), design =STEPSCleanTreatment)
anova_result <- anova(mod_empty, modage, test = "F")

# Display the result
anova_result
round(
  cbind(
    summary(modage,
              df.resid=degf(modage$survey.design))$coef,
    confint(modage,
              ddf=degf(modage$survey.design))
  )
, 4)

# Compute odds ratios and their 95% confidence intervals
OR.CI <- cbind("AOR" = exp(   coef(modage)),
                       exp(confint(modage,
                           df.resid=degf(modage$survey.design))))[-1,]
kable(round(OR.CI, 3))


#Sex
modsex <- svyglm(treatment ~ gender , family = quasibinomial(), design =STEPSCleanTreatment)
anova_result <- anova(mod_empty, modsex, test = "F")

# Display the result
anova_result
round(
  cbind(
    summary(modsex,
              df.resid=degf(modsex$survey.design))$coef,
    confint(modsex,
              ddf=degf(modsex$survey.design))
  )
, 4)

# Compute odds ratios and their 95% confidence intervals
OR.CI <- cbind("AOR" = exp(   coef(modsex)),
                       exp(confint(modsex,
                           df.resid=degf(modsex$survey.design))))[-1,]
kable(round(OR.CI, 3))


#Residence
modres <- svyglm(treatment ~ residence , family = quasibinomial(), design =STEPSCleanTreatment)
anova_result <- anova(mod_empty, modres, test = "F")

# Display the result
anova_result
round(
  cbind(
    summary(modres,
              df.resid=degf(modres$survey.design))$coef,
    confint(modres,
              ddf=degf(modres$survey.design))
  )
, 4)

# Compute odds ratios and their 95% confidence intervals
OR.CI <- cbind("AOR" = exp(   coef(modres)),
                       exp(confint(modres,
                           df.resid=degf(modres$survey.design))))[-1,]
kable(round(OR.CI, 3))


#Highest education level
modedl <- svyglm(treatment ~ edlev , family = quasibinomial(), design =STEPSCleanTreatment)
anova_result <- anova(mod_empty, modedl, test = "F")

# Display the result
anova_result
round(
  cbind(
    summary(modedl,
              df.resid=degf(modedl$survey.design))$coef,
    confint(modedl,
              ddf=degf(modedl$survey.design))
  )
, 4)

# Compute odds ratios and their 95% confidence intervals
OR.CI <- cbind("AOR" = exp(   coef(modedl)),
                       exp(confint(modedl,
                           df.resid=degf(modedl$survey.design))))[-1,]
kable(round(OR.CI, 3))

#Cholesterol level
modedl <- svyglm(treatment ~ cholesterol_class , family = quasibinomial(), design =STEPSCleanTreatment)
anova_result <- anova(mod_empty, modedl, test = "F")

# Display the result
anova_result
round(
  cbind(
    summary(modedl,
            df.resid=degf(modedl$survey.design))$coef,
    confint(modedl,
            ddf=degf(modedl$survey.design))
  )
  , 4)

# Compute odds ratios and their 95% confidence intervals
OR.CI <- cbind("AOR" = exp(   coef(modedl)),
               exp(confint(modedl,
                           df.resid=degf(modedl$survey.design))))[-1,]
kable(round(OR.CI, 3))


# Occupation
modocc <- svyglm(treatment ~ occupation , family = quasibinomial(), design =STEPSCleanTreatment)
anova_result <- anova(mod_empty, modocc, test = "F")

# Display the result
anova_result
round(
  cbind(
    summary(modocc,
              df.resid=degf(modocc$survey.design))$coef,
    confint(modocc,
              ddf=degf(modocc$survey.design))
  )
, 4)

# Compute odds ratios and their 95% confidence intervals
OR.CI <- cbind("AOR" = exp(   coef(modocc)),
                       exp(confint(modocc,
                           df.resid=degf(modocc$survey.design))))[-1,]
kable(round(OR.CI, 3))


#Marital status
modmar <- svyglm(treatment ~ mstatus , family = quasibinomial(), design =STEPSCleanTreatment)
anova_result <- anova(mod_empty, modmar, test = "F")

# Display the result
anova_result
round(
  cbind(
    summary(modmar,
              df.resid=degf(modmar$survey.design))$coef,
    confint(modmar,
              ddf=degf(modmar$survey.design))
  )
, 4)

# Compute odds ratios and their 95% confidence intervals
OR.CI <- cbind("AOR" = exp(   coef(modmar)),
                       exp(confint(modmar,
                           df.resid=degf(modmar$survey.design))))[-1,]
kable(round(OR.CI, 3))


# Diabetes
moddbm <- svyglm(treatment ~ dbm , family = quasibinomial(), design =STEPSCleanTreatment)
anova_result <- anova(mod_empty, moddbm, test = "F")

# Display the result
anova_result
round(
  cbind(
    summary(moddbm,
              df.resid=degf(moddbm$survey.design))$coef,
    confint(moddbm,
              ddf=degf(moddbm$survey.design))
  )
, 4)

# Compute odds ratios and their 95% confidence intervals
OR.CI <- cbind("AOR" = exp(   coef(moddbm)),
                       exp(confint(moddbm,
                           df.resid=degf(moddbm$survey.design))))[-1,]
kable(round(OR.CI, 3))


# Hypertension
modhtn <- svyglm(treatment ~ bp , family = quasibinomial(), design =STEPSCleanTreatment)
anova_result <- anova(mod_empty, modhtn, test = "F")

# Display the result
anova_result
round(
  cbind(
    summary(modhtn,
              df.resid=degf(modhtn$survey.design))$coef,
    confint(modhtn,
              ddf=degf(modhtn$survey.design))
  )
, 4)

# Compute odds ratios and their 95% confidence intervals
OR.CI <- cbind("AOR" = exp(   coef(modhtn)),
                       exp(confint(modhtn,
                           df.resid=degf(modhtn$survey.design))))[-1,]
kable(round(OR.CI, 3))


# High salt uptake
modhsu <- svyglm(treatment ~ highsalt , family = quasibinomial(), design =STEPSCleanTreatment)
anova_result <- anova(mod_empty, modhsu, test = "F")

# Display the result
anova_result
round(
  cbind(
    summary(modhsu,
              df.resid=degf(modhsu$survey.design))$coef,
    confint(modhsu,
              ddf=degf(modhsu$survey.design))
  )
, 4)

# Compute odds ratios and their 95% confidence intervals
OR.CI <- cbind("AOR" = exp(   coef(modhsu)),
                       exp(confint(modhsu,
                           df.resid=degf(modhsu$survey.design))))[-1,]
kable(round(OR.CI, 3))


# Low fruit uptake
modlfu <- svyglm(treatment ~ lfi , family = quasibinomial(), design =STEPSCleanTreatment)
anova_result <- anova(mod_empty, modlfu, test = "F")

# Display the result
anova_result
round(
  cbind(
    summary(modlfu,
              df.resid=degf(modlfu$survey.design))$coef,
    confint(modlfu,
              ddf=degf(modlfu$survey.design))
  )
, 4)

# Compute odds ratios and their 95% confidence intervals
OR.CI <- cbind("AOR" = exp(   coef(modlfu)),
                       exp(confint(modlfu,
                           df.resid=degf(modlfu$survey.design))))[-1,]
kable(round(OR.CI, 3))


# Low vegetable uptake
modlvu <- svyglm(treatment ~ lvi , family = quasibinomial(), design =STEPSCleanTreatment)
anova_result <- anova(mod_empty, modlvu, test = "F")

# Display the result
anova_result
round(
  cbind(
    summary(modlvu,
              df.resid=degf(modlvu$survey.design))$coef,
    confint(modlvu,
              ddf=degf(modlvu$survey.design))
  )
, 4)

# Compute odds ratios and their 95% confidence intervals
OR.CI <- cbind("AOR" = exp(   coef(modlvu)),
                       exp(confint(modlvu,
                           df.resid=degf(modlvu$survey.design))))[-1,]
kable(round(OR.CI, 3))


# Physical activity
modpa <- svyglm(treatment ~ pa , family = quasibinomial(), design =STEPSCleanTreatment)
anova_result <- anova(mod_empty, modpa, test = "F")

# Display the result
anova_result
round(
  cbind(
    summary(modpa,
              df.resid=degf(modpa$survey.design))$coef,
    confint(modpa,
              ddf=degf(modpa$survey.design))
  )
, 4)

# Compute odds ratios and their 95% confidence intervals
OR.CI <- cbind("AOR" = exp(   coef(modpa)),
                       exp(confint(modpa,
                           df.resid=degf(modpa$survey.design))))[-1,]
kable(round(OR.CI, 3))


# Tobacco use
modtu <- svyglm(treatment ~ smokehis , family = quasibinomial(), design =STEPSCleanTreatment)
anova_result <- anova(mod_empty, modtu, test = "F")

# Display the result
anova_result
round(
  cbind(
    summary(modtu,
              df.resid=degf(modtu$survey.design))$coef,
    confint(modtu,
              ddf=degf(modtu$survey.design))
  )
, 4)

# Compute odds ratios and their 95% confidence intervals
OR.CI <- cbind("AOR" = exp(   coef(modtu)),
                       exp(confint(modtu,
                           df.resid=degf(modtu$survey.design))))[-1,]
kable(round(OR.CI, 3))


# Harmful alcohol use
modhaul <- svyglm(treatment ~ alcharm , family = quasibinomial(), design =STEPSCleanTreatment)
anova_result <- anova(mod_empty, modhaul, test = "F")

# Display the result
anova_result
round(
  cbind(
    summary(modhaul,
              df.resid=degf(modhaul$survey.design))$coef,
    confint(modhaul,
              ddf=degf(modhaul$survey.design))
  )
, 4)

# Compute odds ratios and their 95% confidence intervals
OR.CI <- cbind("AOR" = exp(   coef(modhaul)),
                       exp(confint(modhaul,
                           df.resid=degf(modhaul$survey.design))))[-1,]
kable(round(OR.CI, 3))


# BMI
modbmi <- svyglm(treatment ~ bmi_cat , family = quasibinomial(), design =STEPSCleanTreatment)
anova_result <- anova(mod_empty, modbmi, test = "F")

# Display the result
anova_result
round(
  cbind(
    summary(modbmi,
              df.resid=degf(modbmi$survey.design))$coef,
    confint(modbmi,
              ddf=degf(modbmi$survey.design))
  )
, 4)

# Compute odds ratios and their 95% confidence intervals
OR.CI <- cbind("AOR" = exp(   coef(modbmi)),
                       exp(confint(modbmi,
                           df.resid=degf(modbmi$survey.design))))[-1,]
kable(round(OR.CI, 3))


# Survey year
modsyr <- svyglm(treatment ~ survyear , family = quasibinomial(), design =STEPSCleanTreatment)
anova_result <- anova(mod_empty, modsyr, test = "F")

# Display the result
anova_result
round(
  cbind(
    summary(modsyr,
              df.resid=degf(modsyr$survey.design))$coef,
    confint(modsyr,
              ddf=degf(modsyr$survey.design))
  )
, 4)

# Compute odds ratios and their 95% confidence intervals
OR.CI <- cbind("AOR" = exp(   coef(modsyr)),
                       exp(confint(modsyr,
                           df.resid=degf(modsyr$survey.design))))[-1,]
kable(round(OR.CI, 3))


```


Multiple regression

1.Prevalence of CVD

```{r, eval=TRUE, tidy = TRUE, size="small", cache=FALSE, results="asis", message=F, warning=F}
 
#Full model
modprevalence <- svyglm(cvd ~ age_cat + gender + residence + bmi_cat + edlev + occupation + mstatus + dbm + bp + highsalt + lfi + lvi + pa + smokehis + alcharm + survyear + cholesterol_class + country, family=quasibinomial(), design =STEPSClean)

step.model <- modprevalence %>% stepAIC(trace = FALSE)
coef(step.model)


# Compute odds ratios and their 95% confidence intervals for the Final Model
OR.CI <- cbind("AOR" = exp(   coef(step.model)),
                       exp(confint(step.model,
                           df.resid=degf(step.model$survey.design))))[-1,]
kable(round(OR.CI, 3))


# Compute odds ratios and their 95% confidence intervals for the Full Model
OR.CI <- cbind("AOR" = exp(   coef(modprevalence)),
                       exp(confint(modprevalence,
                           df.resid=degf(modprevalence$survey.design))))[-1,]
kable(round(OR.CI, 3))

#Automate F-test
# Perform F-test for the significant variables in the final model
anova_result <- anova(step.model, test = "F")

# Display the result of the F-test
pander::pander(anova_result, caption = "ANOVA F-Test for Final Model Variables")

```


2. Uptake of prevention for CVD 

```{r, eval=TRUE, tidy = TRUE, size="small", cache=FALSE, results="asis", message=F, warning=F}

modprev <-  svyglm(treatment ~ age_cat + gender + residence + bmi_cat + edlev + occupation + mstatus + dbm + bp + highsalt + lfi + lvi + pa + smokehis + alcharm + survyear + cholesterol_class + country, family=binomial(), design =STEPSCleanPrevention)

step.modprev <- modprev %>% stepAIC(trace = FALSE)
coef(step.modprev)


# Compute odds ratios and their 95% confidence intervals
OR.CI <- cbind("AOR" = exp(   coef(step.modprev)),
               exp(confint(step.modprev,
                           df.resid=degf(step.modprev$survey.design))))[-1,]
kable(round(OR.CI, 3))

#Automate F-test
# Perform F-test for the significant variables in the final model
anova_result <- anova(step.modprev, test = "F")

# Display the result of the F-test
pander::pander(anova_result, caption = "ANOVA F-Test for Final Model Variables")

```


3. Uptake of treatment for CVD

```{r, eval=TRUE, tidy = TRUE, size="small", cache=FALSE, results="asis", message=F, warning=F}

modtrt <-  svyglm(treatment ~ age_cat + gender + residence + bmi_cat + edlev + occupation + mstatus + dbm + bp + highsalt + lfi + lvi + pa + smokehis + alcharm + survyear + cholesterol_class + country, family=binomial(), design =STEPSCleanTreatment)

step.model.trt <- modtrt %>% stepAIC(trace = FALSE)

round(
  cbind(
    summary(step.model.trt,
              df.resid=degf(step.model.trt$survey.design))$coef,
    confint(step.model.trt,
              ddf=degf(step.model.trt$survey.design))
  )
, 4)
 

# Compute odds ratios and their 95% confidence intervals 
OR.CI <- cbind("AOR" = exp(   coef(step.model.trt)),
                       exp(confint(step.model.trt,
                           df.resid=degf(step.model.trt$survey.design))))[-1,]
kable(round(OR.CI, 3))


#Automate F-test
# Perform F-test for the significant variables in the final model
anova_result <- anova(step.model.trt, test = "F")

# Display the result of the F-test
pander::pander(anova_result, caption = "ANOVA F-Test for Final Model Variables")
```


## Model sensitivity

### CVD prevalence

```{r mod-sensi-cvd-preval, include=T}

# Load necessary packages
library(stats)
library(cluster)
library(data.table)
library(dplyr)
library(Rtsne)
library(ggplot2)
library(dendextend)
library(clValid)
library(tableone)
library(gtsummary)
library(glmnet)
library(WeightedROC)
library(ranger)
library(scoring)
library(DescTools)
library(ggplot2)
library(mlr3misc)
library(riskRegression)

#CVD prevalence
Formula=as.formula("cvd ~ age_cat + gender + residence + bmi_cat + edlev + occupation + mstatus + dbm + bp + highsalt + lfi + lvi + pa + smokehis + cholesterol_class+ alcharm + survyear + country")

cvd_africa_2024_ML <- cvd_africa_2024

dat.analytic <- cvd_africa_2024_ML

myvars = c("age_cat","bmi_cat", "gender", "residence", "edlev", "bp", "highsalt", "lfi", "smokehis","dbm","lvi","occupation","survyear","pa","mstatus","cholesterol_class","country","alcharm","cvd", "wstep1")
dat.analytic <- dat.analytic[myvars]

#Split-sample
set.seed(604001)
dat.analytic$datasplit <- rbinom(nrow(dat.analytic), size = 1, prob = 0.7) 
table(dat.analytic$datasplit)

# Training data
dat.train <- dat.analytic %>%
  dplyr::filter(dat.analytic$datasplit == 1)
dim(dat.train)

# Test data
dat.test <- dat.analytic %>%
  filter(dat.analytic$datasplit == 0)
dim(dat.test)

##LASSO for surveys
# Training data - X: predictor, y: outcome
X.train <- model.matrix(Formula, dat.train)[,c(-19,-20,-21)] 
y.train <- as.matrix(dat.train$cvd) 
w.train <- as.matrix(dat.train$wstep1)

# Test data - X: predictor, y: outcome
X.test <- model.matrix(Formula, dat.test)[,c(-19,-20,-21)] 
y.test <- as.matrix(dat.test$cvd) 
w.test <- as.matrix(dat.test$wstep1)

#Find best lambda
fit.cv.lasso <- cv.glmnet(x = X.train, y = y.train, 
                          nfolds = 5, alpha = 1, 
                          family = "binomial", 
                          weights = dat.train$wstep1)
fit.cv.lasso


#We can also plot all the lambda values against the deviance (i.e., prediction error).
#plot(fit.cv.lasso)


# Best lambda
fit.cv.lasso$lambda.min

# Fit the model on the training set with optimum lambda
fit.lasso <- glmnet(x = X.train, y = y.train, 
                    alpha = 1, family = "binomial",
                    lambda = fit.cv.lasso$lambda.min, 
                    weights = dat.train$wstep1)
fit.lasso

#Let’s check the coefficients from the model:
# Intercept 
fit.lasso$a0

# Beta coefficients
fit.lasso$beta

# Pr.(CVD = Yes) on the test set
dat.test$pred.lasso <- predict(fit.lasso, 
                               newx = X.test, 
                               type = "response")
head(dat.test$pred.lasso)

#Model performance
# AUC on the test set with sampling weights
auc.lasso <- WeightedAUC(WeightedROC(dat.test$pred.lasso, 
                                     dat.test$cvd, 
                                     weight = dat.test$wstep1))
auc.lasso

# Logit of the predicted probability
dat.test$pred.lasso.logit <- Logit(dat.test$pred.lasso)

# Weighted calibration slope
mod.cal <- glm(cvd ~ pred.lasso.logit, data = dat.test, 
               family = binomial, weights = wstep1)
cal.slope.lasso <- summary(mod.cal)$coef[2,1]
cal.slope.lasso

# Weighted Brier Score
brier.lasso <- Score(object = list(pred.lasso = dat.test$pred.lasso), 
                     formula = cvd ~ 1, 
                     data = dat.test, 
                     weights = dat.test$wstep1, 
                     metrics = "brier")
                     
brier.lasso


#Random Forest for Surveys
Formula

#Hyperparameter tuning
grid.search <- expand.grid(mtry = 5:7, node.size = 1:3, 
                           num.trees = seq(200,500,100), 
                           oob_error = 0)
head(grid.search)

# Install and load ranger if needed
if (!requireNamespace("ranger", quietly = TRUE)) {
  install.packages("ranger")
}
library(ranger)

# Loop over each row of the grid search
for(ii in 1:nrow(grid.search)) {
  
  # Fit a random forest model on the training set with the parameters from the grid
  fit.rf.tune <- ranger(
    formula = Formula, 
    data = dat.train, 
    num.trees = grid.search$num.trees[ii],
    mtry = grid.search$mtry[ii], 
    min.node.size = grid.search$node.size[ii],
    importance = 'impurity', 
    case.weights = dat.train$wstep1, 
    oob.error = TRUE  # Ensure OOB error is computed
  )
  
  # Store the Out-of-Bag (OOB) error for the current grid configuration
  grid.search$oob_error[ii] <- sqrt(fit.rf.tune$prediction.error)
}

# Display the first few rows of grid search results with the OOB error
head(grid.search)


#Let’s check which combination of hyperparameters (number of trees, number of predictors to split at in each node, and minimal node size to split at) gives minimum prediction error.

position <- which.min(grid.search$oob_error)
grid.search[position,]

# Fit the model on the training set 
fit.rf <- ranger(formula = Formula, 
                 data = dat.train, 
                 case.weights = dat.train$wstep1, 
                 probability = T,
                 num.trees = grid.search$num.trees[position], 
                 min.node.size = grid.search$node.size[position], 
                 mtry = grid.search$mtry[position], 
                 importance = 'impurity')

# Fitted random forest model
fit.rf

#Now, we can use the model to make predictions on our test data.

# Pr.(CVD = Yes) on the test set
dat.test$pred.rf <- predict(fit.rf, 
                            data = dat.test)$predictions[,2]
head(dat.test$pred.rf)

#Model performance
#The same as the LASSO model, we can calculate the AUC, calibration slope, and Brier score.

# AUC on the test set with sampling weights
auc.rf <- WeightedAUC(WeightedROC(dat.test$pred.rf, 
                                  dat.test$cvd, 
                                  weight = dat.test$wstep1))
auc.rf

# Logit of the predicted probability
dat.test$pred.rf[dat.test$pred.rf == 0] <- 0.00001
dat.test$pred.rf.logit <- Logit(dat.test$pred.rf)

# Weighted calibration slope
# mod.cal <- glm(cvd ~ pred.rf.logit, 
#                data = dat.test, 
#                family = binomial, 
#                weights = wstep1)
# 
# cal.slope.rf <- summary(mod.cal)$coef[2,1]
# cal.slope.rf


# Weighted Brier Score
brier.rf <- mean(brierscore(cvd ~ dat.test$pred.rf, 
                            data = dat.test,
                            wt = dat.test$wstep1))
brier.rf

#Variable importance
#One nice feature of random forest is that we can rank the variables and generate a variable importance plot.

# Create a lookup vector for variable labels
variable_labels <- c(
  "age_cat"="Age",
  "bmi_cat"="BMI", 
  "gender"="Gender", 
  "residence"="Area of residence", 
  "edlev"="Highest education level", 
  "bp"="Hypertension status", 
  "highsalt"="High salt uptake", 
  "lfi"="Low fruit uptake", 
  "smokehis"="Smoking status",
  "dbm"="Status of diabetes",
  "lvi"="Low vegetable uptake",
  "occupation"="Occupation",
  "survyear"="Survey year",
  "pa"="Physical activity",
  "mstatus"="Marital status",
  "country"="Country",
  "alcharm"="Harmful alcohol uptake"
)

# Generate variable importance plot with variable labels
ggplot(
  enframe(fit.rf$variable.importance, 
          name = "variable", 
          value = "importance") %>%
    mutate(variable = variable_labels[variable]) %>%  # Use labels
    filter(!is.na(variable), !is.na(importance)),     # Exclude NAs
  aes(x = reorder(variable, importance), 
      y = importance, fill = importance)) +
  geom_bar(stat = "identity", 
           position = "dodge") +
  coord_flip() +
  ylab("Variable importance") +  
  xlab("") + 
  ggtitle("") +
  guides(fill = "none") +
  scale_fill_gradient(low = "grey", 
                      high = "grey10") + 
  theme_bw()

```


### Uptake of CVD treatment amongst those with CVD

```{r treat-prev-sens-mi, include=T}

##Prevention of CVD
cvd_africa_2024_treatment_ML <- cvd_africa_2024_treatment

dat.analytic <- cvd_africa_2024_treatment_ML

myvars = c("age_cat","bmi_cat", "gender", "residence", "edlev", "bp", "highsalt", "lfi", "smokehis","dbm","lvi","occupation","survyear","pa","mstatus","country","alcharm","treatment", "psu","stratum","wstep1")
dat.analytic <- dat.analytic[myvars]

Formula=as.formula("treatment ~ age_cat + gender + residence + bmi_cat + edlev + occupation + mstatus + dbm + bp + highsalt + lfi + lvi + pa + smokehis + alcharm + survyear + country")


#Split-sample
set.seed(604001)
dat.analytic$datasplit <- rbinom(nrow(dat.analytic), size = 1, prob = 0.7) 
table(dat.analytic$datasplit)

# Training data
dat.train <- dat.analytic %>%
  dplyr::filter(dat.analytic$datasplit == 1)
dim(dat.train)

# Test data
dat.test <- dat.analytic %>%
  filter(dat.analytic$datasplit == 0)
dim(dat.test)

##LASSO for surveys
# Training data - X: predictor, y: outcome
X.train <- model.matrix(Formula, dat.train)[,-c(18,19,20,21)] 
y.train <- as.matrix(dat.train$treatment) 
w.train <- as.matrix(dat.train$wstep1)

# Test data - X: predictor, y: outcome
X.test <- model.matrix(Formula, dat.test)[,-c(18,19,20,21)]
y.test <- as.matrix(dat.test$treatment) 
w.test <- as.matrix(dat.test$wstep1)

#Find best lambda
fit.cv.lasso <- cv.glmnet(x = X.train, y = y.train, 
                          nfolds = 5, alpha = 1, 
                          family = "binomial", 
                          weights = w.train)
fit.cv.lasso


#We can also plot all the lambda values against the deviance (i.e., prediction error).
plot(fit.cv.lasso)


# Best lambda
fit.cv.lasso$lambda.min

# Fit the model on the training set with optimum lambda
fit.lasso <- glmnet(x = X.train, y = y.train, 
                    alpha = 1, family = "binomial",
                    lambda = fit.cv.lasso$lambda.min, 
                    weights = dat.train$wstep1)
fit.lasso

#Let’s check the coefficients from the model:
# Intercept 
fit.lasso$a0

# Beta coefficients
fit.lasso$beta

# Pr.(CVD = Yes) on the test set
dat.test$pred.lasso <- predict(fit.lasso, 
                               newx = X.test, 
                               type = "response")
head(dat.test$pred.lasso)

#Model performance
# AUC on the test set with sampling weights
auc.lasso <- WeightedAUC(WeightedROC(dat.test$pred.lasso, 
                                     dat.test$treatment, 
                                     weight = dat.test$wstep1))
auc.lasso

# Logit of the predicted probability
dat.test$pred.lasso.logit <- logit(dat.test$pred.lasso)

# Weighted calibration slope
mod.cal <- glm(treatment ~ pred.lasso.logit, data = dat.test, 
               family = binomial, weights = wstep1)
cal.slope.lasso <- summary(mod.cal)$coef[2,1]
cal.slope.lasso

# Weighted Brier Score
brier.lasso <- mean(brierscore(treatment ~ dat.test$pred.lasso, 
                               data = dat.test, 
                               wt = dat.test$wstep1))
brier.lasso

#Random Forest for Surveys
Formula

#Hyperparameter tuning
grid.search <- expand.grid(mtry = 5:7, node.size = 1:3, 
                           num.trees = seq(200,500,100), 
                           oob_error = 0)
head(grid.search)

## Calculate prediction error for each grid 
for(ii in 1:nrow(grid.search)) {
  # Model on training set with grid
  fit.rf.tune <- ranger(formula = Formula, 
                        data = dat.train, 
                        num.trees = grid.search$num.trees[ii],
                        mtry = grid.search$mtry[ii], 
                        min.node.size = grid.search$node.size[ii],
                        importance = 'impurity', 
                        case.weights = dat.train$wstep1)
  
  # Add Out-of-bag (OOB) error to each grid
  grid.search$oob_error[ii] <- sqrt(fit.rf.tune$prediction.error)
}
head(grid.search)

#Let’s check which combination of hyperparameters (number of trees, number of predictors to split at in each node, and minimal node size to split at) gives minimum prediction error.

position <- which.min(grid.search$oob_error)
grid.search[position,]

# Fit the model on the training set 
fit.rf <- ranger(formula = Formula, 
                 data = dat.train, 
                 case.weights = dat.train$wstep1, 
                 probability = T,
                 num.trees = grid.search$num.trees[position], 
                 min.node.size = grid.search$node.size[position], 
                 mtry = grid.search$mtry[position], 
                 importance = 'impurity')

# Fitted random forest model
fit.rf

#Now, we can use the model to make predictions on our test data.

# Pr.(CVD = Yes) on the test set
dat.test$pred.rf <- predict(fit.rf, 
                            data = dat.test)$predictions[,2]
head(dat.test$pred.rf)

#Model performance
#The same as the LASSO model, we can calculate the AUC, calibration slope, and Brier score.

# AUC on the test set with sampling weights
auc.rf <- WeightedAUC(WeightedROC(dat.test$pred.rf, 
                                  dat.test$treatment, 
                                  weight = dat.test$wstep1))
auc.rf

# Logit of the predicted probability
dat.test$pred.rf[dat.test$pred.rf == 0] <- 0.00001
dat.test$pred.rf.logit <- Logit(dat.test$pred.rf)

# Weighted calibration slope
# mod.cal <- glm(cvd ~ pred.rf.logit, 
#                data = dat.test, 
#                family = binomial, 
#                weights = wstep1)
# 
# cal.slope.rf <- summary(mod.cal)$coef[2,1]
# cal.slope.rf


# Weighted Brier Score
brier.rf <- mean(brierscore(treatment ~ dat.test$pred.rf, 
                            data = dat.test,
                            wt = dat.test$wstep1))
brier.rf

#Variable importance
#One nice feature of random forest is that we can rank the variables and generate a variable importance plot.

# Create a lookup vector for variable labels
variable_labels <- c(
  "age_cat"="Age",
  "bmi_cat"="BMI", 
  "gender"="Gender", 
  "residence"="Area of residence", 
  "edlev"="Highest education level", 
  "bp"="Hypertension status", 
  "highsalt"="High salt uptake", 
  "lfi"="Low fruit uptake", 
  "smokehis"="Smoking status",
  "dbm"="Status of diabetes",
  "lvi"="Low vegetable uptake",
  "occupation"="Occupation",
  "survyear"="Survey year",
  "pa"="Physical activity",
  "mstatus"="Marital status",
  "country"="Country",
  "alcharm"="Harmful alcohol uptake"
)

# Generate variable importance plot with variable labels
ggplot(
  enframe(fit.rf$variable.importance, 
          name = "variable", 
          value = "importance") %>%
    mutate(variable = variable_labels[variable]), # Use labels
  aes(x = reorder(variable, importance), 
      y = importance, fill = importance)) +
  geom_bar(stat = "identity", 
           position = "dodge") +
  coord_flip() +
  ylab("Variable importance") +  
  xlab("") + 
  ggtitle("") +
  guides(fill = "none") +
  scale_fill_gradient(low = "grey", 
                      high = "grey10") + 
  theme_bw() 
```

### Uptake of treatment for CVD prevention

```{r sensi_cvd-rx-mi, include=T}
##Uptake of treatment of CVD prevention
cvd_africa_2024_prevention_ML <-cvd_africa_2024_prevention


dat.analytic <- cvd_africa_2024_prevention_ML

myvars = c("age_cat","bmi_cat", "gender", "residence", "edlev", "bp", "highsalt", "lfi", "smokehis","dbm","lvi","occupation","survyear","pa","mstatus","country","alcharm","trtcvd", "psu","stratum","wstep1")
dat.analytic <- dat.analytic[myvars]


Formula=as.formula("trtcvd ~ age_cat + gender + residence + bmi_cat + edlev + occupation + mstatus + dbm + bp + highsalt + lfi + lvi + pa + smokehis + alcharm + survyear + country")


#Split-sample
set.seed(604001)
dat.analytic$datasplit <- rbinom(nrow(dat.analytic), size = 1, prob = 0.7) 
table(dat.analytic$datasplit)

# Training data
dat.train <- dat.analytic %>%
  dplyr::filter(dat.analytic$datasplit == 1)
dim(dat.train)

# Test data
dat.test <- dat.analytic %>%
  filter(dat.analytic$datasplit == 0)
dim(dat.test)

##LASSO for surveys
# Training data - X: predictor, y: outcome
X.train <- model.matrix(Formula, dat.train)[,-c(18,19,20,21)] 
y.train <- as.matrix(dat.train$trtcvd) 
w.train <- as.matrix(dat.train$wstep1)

# Test data - X: predictor, y: outcome
X.test <- model.matrix(Formula, dat.test)[,-c(18,19,20,21)]
y.test <- as.matrix(dat.test$trtcvd) 
w.test <- as.matrix(dat.test$wstep1)

#Find best lambda
fit.cv.lasso <- cv.glmnet(x = X.train, y = y.train, 
                          nfolds = 5, alpha = 1, 
                          family = "binomial", 
                          weights = w.train)
fit.cv.lasso


#We can also plot all the lambda values against the deviance (i.e., prediction error).
plot(fit.cv.lasso)


# Best lambda
fit.cv.lasso$lambda.min

# Fit the model on the training set with optimum lambda
fit.lasso <- glmnet(x = X.train, y = y.train, 
                    alpha = 1, family = "binomial",
                    lambda = fit.cv.lasso$lambda.min, 
                    weights = dat.train$wstep1)
fit.lasso

#Let’s check the coefficients from the model:
# Intercept 
fit.lasso$a0

# Beta coefficients
fit.lasso$beta

# Pr.(CVD = Yes) on the test set
dat.test$pred.lasso <- predict(fit.lasso, 
                               newx = X.test, 
                               type = "response")
head(dat.test$pred.lasso)

#Model performance
# AUC on the test set with sampling weights
auc.lasso <- WeightedAUC(WeightedROC(dat.test$pred.lasso, 
                                     dat.test$trtcvd, 
                                     weight = dat.test$wstep1))
auc.lasso

# Logit of the predicted probability
dat.test$pred.lasso.logit <- Logit(dat.test$pred.lasso)

# Weighted calibration slope
mod.cal <- glm(trtcvd ~ pred.lasso.logit, data = dat.test, 
               family = binomial, weights = wstep1)
cal.slope.lasso <- summary(mod.cal)$coef[2,1]
cal.slope.lasso

# Weighted Brier Score
brier.lasso <- mean(brierscore(trtcvd ~ dat.test$pred.lasso, 
                               data = dat.test, 
                               wt = dat.test$wstep1))
brier.lasso

#Random Forest for Surveys
Formula

#Hyperparameter tuning
grid.search <- expand.grid(mtry = 5:7, node.size = 1:3, 
                           num.trees = seq(200,500,100), 
                           oob_error = 0)
head(grid.search)
 
## Calculate prediction error for each grid  
for(ii in 1:nrow(grid.search)) {
  # Model on training set with grid
  fit.rf.tune <- ranger(formula = Formula, 
                        data = dat.train, 
                        num.trees = grid.search$num.trees[ii],
                        mtry = grid.search$mtry[ii], 
                        min.node.size = grid.search$node.size[ii],
                        importance = 'impurity', 
                        case.weights = dat.train$wstep1)
  
  # Add Out-of-bag (OOB) error to each grid
  grid.search$oob_error[ii] <- sqrt(fit.rf.tune$prediction.error)
}
head(grid.search)

#Let’s check which combination of hyperparameters (number of trees, number of predictors to split at in each node, and minimal node size to split at) gives minimum prediction error.

position <- which.min(grid.search$oob_error)
grid.search[position,]

# Fit the model on the training set 
fit.rf <- ranger(formula = Formula, 
                 data = dat.train, 
                 case.weights = dat.train$wstep1, 
                 probability = T,
                 num.trees = grid.search$num.trees[position], 
                 min.node.size = grid.search$node.size[position], 
                 mtry = grid.search$mtry[position], 
                 importance = 'impurity')

# Fitted random forest model
fit.rf

#Now, we can use the model to make predictions on our test data.

# Pr.(CVD = Yes) on the test set
dat.test$pred.rf <- predict(fit.rf, 
                            data = dat.test)$predictions[,2]
head(dat.test$pred.rf)

#Model performance
#The same as the LASSO model, we can calculate the AUC, calibration slope, and Brier score.

# AUC on the test set with sampling weights
auc.rf <- WeightedAUC(WeightedROC(dat.test$pred.rf, 
                                  dat.test$trtcvd, 
                                  weight = dat.test$wstep1))
auc.rf

# Logit of the predicted probability
dat.test$pred.rf[dat.test$pred.rf == 0] <- 0.00001
dat.test$pred.rf.logit <- Logit(dat.test$pred.rf)

# Weighted calibration slope
# mod.cal <- glm(cvd ~ pred.rf.logit, 
#                data = dat.test, 
#                family = binomial, 
#                weights = wstep1)
# 
# cal.slope.rf <- summary(mod.cal)$coef[2,1]
# cal.slope.rf


# Weighted Brier Score
brier.rf <- mean(brierscore(trtcvd ~ dat.test$pred.rf, 
                            data = dat.test,
                            wt = dat.test$wstep1))
brier.rf

#Variable importance
#One nice feature of random forest is that we can rank the variables and generate a variable importance plot.
# Create a lookup vector for variable labels
variable_labels <- c(
  "age_cat"="Age",
  "bmi_cat"="BMI", 
  "gender"="Gender", 
  "residence"="Area of residence", 
  "edlev"="Highest education level", 
  "bp"="Hypertension status", 
  "highsalt"="High salt uptake", 
  "lfi"="Low fruit uptake", 
  "smokehis"="Smoking status",
  "dbm"="Status of diabetes",
  "lvi"="Low vegetable uptake",
  "occupation"="Occupation",
  "survyear"="Survey year",
  "pa"="Physical activity",
  "mstatus"="Marital status",
  "country"="Country",
  "alcharm"="Harmful alcohol uptake"
)

# Generate variable importance plot with variable labels
ggplot(
  enframe(fit.rf$variable.importance, 
          name = "variable", 
          value = "importance") %>%
    mutate(variable = variable_labels[variable]), # Use labels
  aes(x = reorder(variable, importance), 
      y = importance, fill = importance)) +
  geom_bar(stat = "identity", 
           position = "dodge") +
  coord_flip() +
  ylab("Variable importance") +  
  xlab("") + 
  ggtitle("") +
  guides(fill = "none") +
  scale_fill_gradient(low = "grey", 
                      high = "grey10") + 
  theme_bw()  
 
```

CVD care cascade by country
```{r}
# Load required libraries
library(ggplot2)
library(reshape2) 

# Create the data frame
df <- data.frame(
  Country = c("Algeria", "Benin", "Botswana", "Eswatini", "Ethiopia", 
              "Kenya", "Malawi", "Morocco", "São Tomé and Príncipe", 
              "Sudan", "Uganda", "Zambia"),
  Total_population = c(6955, 5115, 3888, 3026, 9241, 4477, 4186, 4991, 2418, 7722, 3973, 4302),
  CVD_diagnosis = c(415, 293, 210, 131, 337, 296, 312, 163, 90, 117, 371, 160),
  CVD_treatment = c(178, 47, 45, 26, 51, 18, 64, 59, 26, 54, 38, 24)
)

# Reshape to long format for ggplot
df_long <- melt(df, id.vars = "Country", 
                variable.name = "Care_cascade", 
                value.name = "Count")

# Set factor levels so legend is ordered correctly
df_long$Care_cascade <- factor(df_long$Care_cascade, 
                               levels = c("Total_population", "CVD_diagnosis", "CVD_treatment"),
                               labels = c("Total population", "CVD diagnosis", "CVD treatment"))

# Plot with numbers on bars
ggplot(df_long, aes(x = Country, y = Count, fill = Care_cascade)) +
  geom_bar(stat = "identity", position = position_dodge(width = 0.9)) +
  geom_text(aes(label = Count), 
            position = position_dodge(width = 0.9), 
            vjust = -0.3, size = 3) +
  labs(title = "CVD Care Cascade by Country",
       x = "Country", y = "Count", fill = "Care cascade") +
  theme_minimal() +
  theme(axis.text.x = element_text(angle = 45, hjust = 1))


```

Forect plot of adjusted CVD diagnosis
```{r}
# Load required libraries
library(ggplot2)
library(dplyr)
library(forcats)

# Create the data frame from the provided table
data <- data.frame(
  Variable = c(
    "Age group", "Age group", "Age group", "Age group", "Age group",
    "Sex", "Sex",
    "Area of residence", "Area of residence",
    "Highest level of education", "Highest level of education", "Highest level of education", "Highest level of education",
    "Country", "Country", "Country", "Country", "Country", "Country", "Country", "Country", "Country", "Country", "Country", "Country",
    "Having hypertension", "Having hypertension",
    "High salt intake", "High salt intake",
    "Low fruit uptake", "Low fruit uptake",
    "Physical activity level", "Physical activity level", "Physical activity level",
    "Smoking history", "Smoking history", "Smoking history"
  ),
  Level = c(
    "15-29 (Ref)", "30-39", "40-49", "50-59", "60+",
    "Female (Ref)", "Male",
    "Rural (Ref)", "Urban",
    "None (Ref)", "Primary", "Secondary", "Tertiary",
    "Algeria (Ref)", "Benin", "Botswana", "Eswatini", "Ethiopia", "Kenya", "Malawi", "Morocco", "STP", "Sudan", "Uganda", "Zambia",
    "No (Ref)", "Yes",
    "No (Ref)", "Yes",
    "No (Ref)", "Yes",
    "Low Level (Ref)", "Moderate level", "High level",
    "Current (Ref)", "Never", "Previous"
  ),
  OR = c(
    1.00, 1.15, 1.25, 1.39, 1.67,
    1.00, 0.65,
    1.00, 1.17,
    1.00, 1.16, 0.91, 0.87,
    1.00, 1.11, 0.91, 0.93, 0.81, 1.09, 1.12, 0.59, 0.54, 0.25, 2.04, 0.57,
    1.00, 2.68,
    1.00, 1.23,
    1.00, 1.09,
    1.00, 1.07, 1.14,
    1.00, 0.69, 0.94
  ),
  CI_lower = c(
    NA, 0.98, 1.03, 1.17, 1.36,
    NA, 0.54,
    NA, 0.97,
    NA, 0.97, 0.74, 0.68,
    NA, 0.77, 0.65, 0.66, 0.58, 0.80, 0.82, 0.46, 0.37, 0.18, 1.55, 0.43,
    NA, 2.32,
    NA, 1.02,
    NA, 0.94,
    NA, 0.88, 0.96,
    NA, 0.55, 0.71
  ),
  CI_upper = c(
    NA, 1.36, 1.51, 1.66, 2.07,
    NA, 0.72,
    NA, 1.38,
    NA, 1.38, 1.11, 1.12,
    NA, 1.58, 1.28, 1.30, 1.14, 1.50, 1.52, 0.76, 0.79, 0.35, 2.69, 0.77,
    NA, 3.10,
    NA, 1.49,
    NA, 1.27,
    NA, 1.29, 1.36,
    NA, 0.86, 1.22
  ),
  P_value = c(
    NA, "<0.0001", "<0.0001", "<0.0001", "<0.0001",
    NA, "<0.0001",
    NA, "0.17",
    NA, "0.0001", "0.0001", "0.0001",
    NA, "<0.0001", "<0.0001", "<0.0001", "<0.0001", "<0.0001", "<0.0001", "<0.0001", "<0.0001", "<0.0001", "<0.0001", "<0.0001",
    NA, "<0.0001",
    NA, "0.41",
    NA, "0.76",
    NA, "0.002", "0.002",
    NA, "<0.0001", "<0.0001"
  )
)

# Create formatted labels for the plot
data <- data %>%
  mutate(
    OR_label = ifelse(is.na(CI_lower), 
                     "1.00 (Reference)", 
                     sprintf("%.2f (%.2f-%.2f)", OR, CI_lower, CI_upper)),
    P_label = ifelse(is.na(P_value), "", P_value),
    Variable_Level = paste(Variable, Level, sep = ": ")
  )

# Filter out reference categories for plotting (keep only rows with CI values)
plot_data <- data %>% 
  filter(!is.na(CI_lower))

# Define the exact order as shown in the uploaded image
variable_order <- c(
  "Age group: 30-39",
  "Age group: 40-49",
  "Age group: 50-59",
  "Age group: 60+",
  "Sex: Male",
  "Area of residence: Urban",
  "Highest level of education: Primary",
  "Highest level of education: Secondary",
  "Highest level of education: Tertiary",
  "Country: Benin",
  "Country: Botswana",
  "Country: Eswatini",
  "Country: Ethiopia",
  "Country: Kenya",
  "Country: Malawi",
  "Country: Morocco",
  "Country: STP",
  "Country: Sudan",
  "Country: Uganda",
  "Country: Zambia",
  "Having hypertension: Yes",
  "High salt intake: Yes",
  "Low fruit uptake: Yes",
  "Physical activity level: Moderate level",
  "Physical activity level: High level",
  "Smoking history: Never",
  "Smoking history: Previous"
)

plot_data <- plot_data %>%
  mutate(Variable_Level = factor(Variable_Level, levels = rev(variable_order)))

# Create the forest plot
forest_plot <- ggplot(plot_data, aes(x = OR, y = Variable_Level)) +
  # Add confidence intervals
  geom_errorbarh(aes(xmin = CI_lower, xmax = CI_upper), 
                 height = 0.2, size = 0.6, color = "black") +
  # Add points for OR estimates
  geom_point(size = 3, shape = 18, color = "black") +
  # Add reference line at OR = 1
  geom_vline(xintercept = 1, linetype = "dashed", color = "red", size = 0.8) +
  # Use log scale for proper symmetry
  scale_x_log10(
    breaks = c(0.10, 0.25, 0.50, 1.00, 2.00, 4.00, 8.00, 12.00),
    limits = c(0.08, 15),
    expand = expansion(mult = c(0.02, 0.02))
  ) +
  # Add labels and title
  labs(
    title = "Adjusted Odds Ratios: OR (95% CI)",
    x = "OR, 95% CI",
    y = NULL
  ) +
  # Apply minimal theme
  theme_minimal() +
  # Customize theme elements to match the uploaded image
  theme(
    legend.position = "none",
    panel.grid.major = element_blank(),
    panel.grid.minor = element_blank(),
    axis.text.y = element_text(size = 10, hjust = 0, face = "bold"),
    axis.text.x = element_text(size = 10),
    axis.title = element_text(face = "bold", size = 12),
    plot.title = element_text(face = "bold", size = 14, hjust = 0.5),
    panel.border = element_rect(color = "black", fill = NA, size = 0.5),
    plot.margin = margin(1, 1, 1, 1, "cm")
  ) +
  # Add OR values and p-values as text on the right side
  geom_text(aes(label = OR_label, x = 15), 
            hjust = 0, vjust = 0.5, size = 3.5, check_overlap = TRUE) +
  # Add p-values
  geom_text(aes(label = P_label, x = 30), 
            hjust = 0, vjust = 0.5, size = 3.5, check_overlap = TRUE) +
  # Ensure proper scaling
  coord_cartesian(xlim = c(0.08, 40), clip = "off")

# Display the plot
print(forest_plot)

# Save the plot to a file
ggsave("adjusted_odds_ratios_forest_plot.png", 
       forest_plot, 
       width = 12, 
       height = 10, 
       dpi = 300,
       bg = "white")

```

CVD prevention
```{r}
# Load required libraries
library(ggplot2)
library(dplyr)
library(forcats)

# Create the data frame from the provided table
data <- data.frame(
  Variable = c(
    "Age group", "Age group", "Age group",
    "Sex", "Sex",
    "Area of residence", "Area of residence",
    "Highest level of education", "Highest level of education", "Highest level of education", "Highest level of education",
    "Marital status", "Marital status", "Marital status",
    "Country", "Country", "Country", "Country", "Country", "Country", "Country", "Country", "Country", "Country", "Country", "Country",
    "Having hypertension", "Having hypertension",
    "High salt intake", "High salt intake",
    "Low fruit uptake", "Low fruit uptake",
    "Low vegetable uptake", "Low vegetable uptake",
    "Physical activity level", "Physical activity level", "Physical activity level",
    "Smoking history", "Smoking history", "Smoking history",
    "Harmful alcohol use", "Harmful alcohol use",
    "BMI", "BMI", "BMI", "BMI"
  ),
  Level = c(
    "40-49 (Ref)", "50-59", "60+",
    "Female (Ref)", "Male",
    "Rural (Ref)", "Urban",
    "None (Ref)", "Primary", "Secondary", "Tertiary",
    "Currently married (Ref)", "Formerly married", "Never married",
    "Algeria (Ref)", "Benin", "Botswana", "Eswatini", "Ethiopia", "Kenya", "Malawi", "Morocco", "STP", "Sudan", "Uganda", "Zambia",
    "No (Ref)", "Yes",
    "No (Ref)", "Yes",
    "No (Ref)", "Yes",
    "No (Ref)", "Yes",
    "Low Level (Ref)", "Moderate level", "High level",
    "Current (Ref)", "Never", "Previous",
    "No (Ref)", "Yes",
    "<18.5 (Ref)", "18.5-24", "25-29", "30+"
  ),
  OR = c(
    1.00, 1.80, 2.40,
    1.00, 0.73,
    1.00, 1.65,
    1.00, 1.04, 1.23, 1.24,
    1.00, 1.37, 0.70,
    1.00, 0.28, 1.51, 0.47, 0.42, 0.22, 0.34, 0.68, 0.52, 0.98, 0.20, 0.15,
    1.00, 16.65,
    1.00, 0.60,
    1.00, 0.80,
    1.00, 0.78,
    1.00, 1.13, 0.78,
    1.00, 0.88, 1.17,
    1.00, 0.66,
    1.00, 0.92, 1.28, 1.36
  ),
  CI_lower = c(
    NA, 1.47, 1.95,
    NA, 0.58,
    NA, 1.33,
    NA, 0.84, 0.95, 0.91,
    NA, 0.98, 0.53,
    NA, 0.19, 1.09, 0.36, 0.27, 0.14, 0.22, 0.55, 0.36, 0.76, 0.12, 0.10,
    NA, 13.93,
    NA, 0.48,
    NA, 0.66,
    NA, 0.63,
    NA, 0.91, 0.63,
    NA, 0.64, 0.83,
    NA, 0.28,
    NA, 0.70, 0.96, 1.01
  ),
  CI_upper = c(
    NA, 2.20, 2.96,
    NA, 0.92,
    NA, 2.04,
    NA, 1.30, 1.59, 1.69,
    NA, 1.92, 0.93,
    NA, 0.40, 2.09, 0.63, 0.65, 0.35, 0.52, 0.84, 0.75, 1.25, 0.36, 0.23,
    NA, 19.91,
    NA, 0.75,
    NA, 0.98,
    NA, 0.96,
    NA, 1.39, 0.96,
    NA, 1.20, 1.64,
    NA, 1.49,
    NA, 1.21, 1.72, 1.82
  ),
  P_value = c(
    NA, "<0.0001", "<0.0001",
    NA, "<0.0001",
    NA, "<0.0001",
    NA, "<0.0001", "<0.0001", "<0.0001",
    NA, "0.01", "0.01",
    NA, "<0.0001", "<0.0001", "<0.0001", "<0.0001", "<0.0001", "<0.0001", "<0.0001", "<0.0001", "<0.0001", "<0.0001", "<0.0001",
    NA, "<0.0001",
    NA, "<0.0001",
    NA, "0.01",
    NA, "0.004",
    NA, "<0.0001", "<0.0001",
    NA, "0.008", "0.008",
    NA, "0.001",
    NA, "<0.0001", "<0.0001", "<0.0001"
  )
)

# Create formatted labels for the plot
data <- data %>%
  mutate(
    OR_label = ifelse(is.na(CI_lower), 
                     "1.00 (Reference)", 
                     sprintf("%.2f (%.2f-%.2f)", OR, CI_lower, CI_upper)),
    P_label = ifelse(is.na(P_value), "", P_value),
    Variable_Level = paste(Variable, Level, sep = ": ")
  )

# Filter out reference categories for plotting (keep only rows with CI values)
plot_data <- data %>% 
  filter(!is.na(CI_lower))

# Define the order for the plot (grouped by variable type)
variable_order <- c(
  # Age group
  "Age group: 50-59", "Age group: 60+",
  # Sex
  "Sex: Male",
  # Area of residence
  "Area of residence: Urban",
  # Education
  "Highest level of education: Primary", "Highest level of education: Secondary", "Highest level of education: Tertiary",
  # Marital status
  "Marital status: Formerly married", "Marital status: Never married",
  # Country
  "Country: Benin", "Country: Botswana", "Country: Eswatini", "Country: Ethiopia", 
  "Country: Kenya", "Country: Malawi", "Country: Morocco", "Country: STP", 
  "Country: Sudan", "Country: Uganda", "Country: Zambia",
  # Clinical factors
  "Having hypertension: Yes",
  # Lifestyle factors
  "High salt intake: Yes", "Low fruit uptake: Yes", "Low vegetable uptake: Yes",
  "Physical activity level: Moderate level", "Physical activity level: High level",
  "Smoking history: Never", "Smoking history: Previous",
  "Harmful alcohol use: Yes",
  # BMI
  "BMI: 18.5-24", "BMI: 25-29", "BMI: 30+"
)

plot_data <- plot_data %>%
  mutate(Variable_Level = factor(Variable_Level, levels = rev(variable_order)))

# Create the forest plot
forest_plot <- ggplot(plot_data, aes(x = OR, y = Variable_Level)) +
  # Add confidence intervals
  geom_errorbarh(aes(xmin = CI_lower, xmax = CI_upper), 
                 height = 0.2, size = 0.6, color = "black") +
  # Add points for OR estimates
  geom_point(size = 3, shape = 18, color = "black") +
  # Add reference line at OR = 1
  geom_vline(xintercept = 1, linetype = "dashed", color = "red", size = 0.8) +
  # Use log scale for proper symmetry (extended range for hypertension OR)
  scale_x_log10(
    breaks = c(0.10, 0.25, 0.50, 1.00, 2.00, 4.00, 8.00, 16.00),
    limits = c(0.08, 35),
    expand = expansion(mult = c(0.02, 0.02))
  ) +
  # Add labels and title
  labs(
    title = "CVD Prevention: Adjusted Odds Ratios (95% CI)",
    x = "OR, 95% CI",
    y = NULL
  ) +
  # Apply minimal theme
  theme_minimal() +
  # Customize theme elements
  theme(
    legend.position = "none",
    panel.grid.major = element_blank(),
    panel.grid.minor = element_blank(),
    axis.text.y = element_text(size = 9, hjust = 0, face = "bold"),
    axis.text.x = element_text(size = 9),
    axis.title = element_text(face = "bold", size = 12),
    plot.title = element_text(face = "bold", size = 14, hjust = 0.5),
    panel.border = element_rect(color = "black", fill = NA, size = 0.5),
    plot.margin = margin(1, 1, 1, 1, "cm")
  ) +
  # Add OR values as text on the right side
  geom_text(aes(label = OR_label, x = 25), 
            hjust = 0, vjust = 0.5, size = 3.2, check_overlap = TRUE) +
  # Add p-values
  geom_text(aes(label = P_label, x = 50), 
            hjust = 0, vjust = 0.5, size = 3.2, check_overlap = TRUE) +
  # Ensure proper scaling
  coord_cartesian(xlim = c(0.08, 60), clip = "off")

# Display the plot
print(forest_plot)

# Save the plot to a file
ggsave("cvd_prevention_forest_plot.png", 
       forest_plot, 
       width = 14, 
       height = 16, 
       dpi = 300,
       bg = "white")


```

CVD treatment

```{r}

# Load required libraries
library(ggplot2)
library(dplyr)
library(forcats)

# Create the data frame from the provided table
data <- data.frame(
  Variable = c(
    "Age group", "Age group", "Age group", "Age group", "Age group",
    "Sex", "Sex",
    "Area of residence", "Area of residence",
    "Country", "Country", "Country", "Country", "Country", "Country", "Country", "Country", "Country", "Country", "Country", "Country",
    "Having hypertension", "Having hypertension",
    "Low fruit uptake", "Low fruit uptake",
    "Low vegetable uptake", "Low vegetable uptake",
    "Physical activity level", "Physical activity level", "Physical activity level",
    "BMI", "BMI", "BMI", "BMI"
  ),
  Level = c(
    "15-29 (Ref)", "30-39", "40-49", "50-59", "60+",
    "Female (Ref)", "Male",
    "Rural (Ref)", "Urban",
    "Algeria (Ref)", "Benin", "Botswana", "Eswatini", "Ethiopia", "Kenya", "Malawi", "Morocco", "STP", "Sudan", "Uganda", "Zambia",
    "No (Ref)", "Yes",
    "No (Ref)", "Yes",
    "No (Ref)", "Yes",
    "Low Level (Ref)", "Moderate level", "High level",
    "<18.5 (Ref)", "18.5-24", "25-29", "30+"
  ),
  OR = c(
    1.00, 0.56, 1.15, 2.31, 2.90,
    1.00, 1.36,
    1.00, 1.36,
    1.00, 0.84, 0.44, 0.49, 0.63, 0.14, 0.39, 0.65, 0.43, 1.56, 0.41, 0.42,
    1.00, 7.10,
    1.00, 0.78,
    1.00, 0.58,
    1.00, 0.69, 0.74,
    1.00, 0.77, 1.06, 1.58
  ),
  CI_lower = c(
    NA, 0.32, 0.67, 1.39, 1.75,
    NA, 0.97,
    NA, 0.97,
    NA, 0.42, 0.16, 0.25, 0.35, 0.07, 0.22, 0.38, 0.19, 0.89, 0.23, 0.21,
    NA, 5.15,
    NA, 0.54,
    NA, 0.54,
    NA, 0.45, 0.50,
    NA, 0.42, 0.57, 0.86
  ),
  CI_upper = c(
    NA, 0.98, 1.96, 3.84, 4.82,
    NA, 1.89,
    NA, 1.90,
    NA, 1.65, 1.20, 1.00, 1.12, 0.30, 0.69, 1.11, 0.99, 2.75, 0.75, 0.83,
    NA, 9.78,
    NA, 1.13,
    NA, 0.83,
    NA, 1.06, 1.10,
    NA, 1.39, 1.93, 2.89
  ),
  P_value = c(
    NA, "<0.0001", "<0.0001", "<0.0001", "<0.0001",
    NA, "0.83",
    NA, "<0.0001",
    NA, "<0.0001", "<0.0001", "<0.0001", "<0.0001", "<0.0001", "<0.0001", "<0.0001", "<0.0001", "<0.0001", "<0.0001", "<0.0001",
    NA, "<0.0001",
    NA, "0.07",
    NA, "0.004",
    NA, "0.001", "0.001",
    NA, "<0.0001", "<0.0001", "<0.0001"
  )
)

# Create formatted labels for the plot
data <- data %>%
  mutate(
    OR_label = ifelse(is.na(CI_lower), 
                     "1.00 (Reference)", 
                     sprintf("%.2f (%.2f-%.2f)", OR, CI_lower, CI_upper)),
    P_label = ifelse(is.na(P_value), "", P_value),
    Variable_Level = paste(Variable, Level, sep = ": ")
  )

# Filter out reference categories for plotting (keep only rows with CI values)
plot_data <- data %>% 
  filter(!is.na(CI_lower))

# Define the order for the plot (grouped by variable type)
variable_order <- c(
  # Age group
  "Age group: 30-39", "Age group: 40-49", "Age group: 50-59", "Age group: 60+",
  # Sex
  "Sex: Male",
  # Area of residence
  "Area of residence: Urban",
  # Country
  "Country: Benin", "Country: Botswana", "Country: Eswatini", "Country: Ethiopia", 
  "Country: Kenya", "Country: Malawi", "Country: Morocco", "Country: STP", 
  "Country: Sudan", "Country: Uganda", "Country: Zambia",
  # Clinical factors
  "Having hypertension: Yes",
  # Lifestyle factors
  "Low fruit uptake: Yes", "Low vegetable uptake: Yes",
  "Physical activity level: Moderate level", "Physical activity level: High level",
  # BMI
  "BMI: 18.5-24", "BMI: 25-29", "BMI: 30+"
)

plot_data <- plot_data %>%
  mutate(Variable_Level = factor(Variable_Level, levels = rev(variable_order)))

# Create the forest plot
forest_plot <- ggplot(plot_data, aes(x = OR, y = Variable_Level)) +
  # Add confidence intervals
  geom_errorbarh(aes(xmin = CI_lower, xmax = CI_upper), 
                 height = 0.2, size = 0.6, color = "black") +
  # Add points for OR estimates
  geom_point(size = 3, shape = 18, color = "black") +
  # Add reference line at OR = 1
  geom_vline(xintercept = 1, linetype = "dashed", color = "red", size = 0.8) +
  # Use log scale for proper symmetry (extended range for hypertension OR)
  scale_x_log10(
    breaks = c(0.10, 0.25, 0.50, 1.00, 2.00, 4.00, 8.00, 16.00),
    limits = c(0.08, 18),
    expand = expansion(mult = c(0.02, 0.02))
  ) +
  # Add labels and title
  labs(
    title = "CVD Treatment: Adjusted Odds Ratios (95% CI)",
    x = "OR, 95% CI",
    y = NULL
  ) +
  # Apply minimal theme
  theme_minimal() +
  # Customize theme elements
  theme(
    legend.position = "none",
    panel.grid.major = element_blank(),
    panel.grid.minor = element_blank(),
    axis.text.y = element_text(size = 9, hjust = 0, face = "bold"),
    axis.text.x = element_text(size = 9),
    axis.title = element_text(face = "bold", size = 12),
    plot.title = element_text(face = "bold", size = 14, hjust = 0.5),
    panel.border = element_rect(color = "black", fill = NA, size = 0.5),
    plot.margin = margin(1, 1, 1, 1, "cm")
  ) +
  # Add OR values as text on the right side
  geom_text(aes(label = OR_label, x = 18), 
            hjust = 0, vjust = 0.5, size = 3.2, check_overlap = TRUE) +
  # Add p-values
  geom_text(aes(label = P_label, x = 30), 
            hjust = 0, vjust = 0.5, size = 3.2, check_overlap = TRUE) +
  # Ensure proper scaling
  coord_cartesian(xlim = c(0.08, 45), clip = "off")

# Display the plot
print(forest_plot)

# Save the plot to a file
ggsave("cvd_treatment_forest_plot.png", 
       forest_plot, 
       width = 14, 
       height = 12, 
       dpi = 300,
       bg = "white")

```
````
