## Supplementary material for "The Prevalence, Prevention, and Treatment of Cardiovascular Diseases in Twelve African Countries (2014-2019): An Analysis of the World Health Organisation STEPwise Approach to Chronic Disease Risk Factor Surveillance": Codes: 4_CVD sensitivity analysis 2025.R

```
#CVD sensitivity analysis

# COMPREHENSIVE SENSITIVITY ANALYSIS FOR IMPUTATION BIAS ASSESSMENT
# Load required libraries
library(dplyr)
library(tidyr)
library(mice)
library(ggplot2)
library(broom)
library(purrr)
library(readxl)

 

# Define variable set
variable_set <- c("age_cat", "gender", "residence", "bmi_cat", 
                  "edlev"
                  , #"occupation", 
                  "bp", 
                  "highsalt", "lfi",  "pa", "smokehis", 
                  "country")

cvd_africa_2024_clean <- read_excel("cvd_africa_2024_clean.xlsx")


# create cvd variable
cvd_africa_2024_clean$cvd <- ifelse(cvd_africa_2024_clean$cvdcase == "Yes", 1, 0)

# check results
table(cvd_africa_2024_clean$cvd)


cvd_africa_2024_clean$bmi_cat <- cut(
  cvd_africa_2024_clean$bmi,
  breaks = c(-Inf, 18.5, 24.9, 29.9, Inf),   # WHO cutoffs
  labels = c("Underweight", "Normal", "Overweight", "Obese"),
  right = TRUE                                # intervals closed on the right (≤)
)


# Check which variables exist in the dataset
existing_vars <- variable_set[variable_set %in% names(cvd_africa_2024_clean)]


# select specific variables
cvdafro_selected <- cvd_africa_2024_clean %>%
  dplyr::select(cvd, age_cat, gender, residence, bmi_cat,
         bp, edlev,
         highsalt, lfi, pa, smokehis,
         country) 

complete_case_data <- cvdafro_selected %>%
  drop_na()


# check the structure of the new dataset
str(cvdafro_selected)


complete_case_model <- glm(
  as.formula(paste("cvd ~", paste(existing_vars[existing_vars != "cvd"], collapse = " + "))),
  data = complete_case_data,
  family = binomial
)

complete_case_results <- tidy(complete_case_model, conf.int = TRUE) %>%
  mutate(method = "complete_case", analysis = "Complete Case")

# 2. SENSITIVITY ANALYSIS WITH MULTIPLE IMPUTATION METHODS
load(here::here("C:\\Users\\LENOVO\\Documents\\GitHub\\CVD_Prevalence_Prevention_Treatment_Paper\\cvd_africa_2024.RData"))

sensitivity_results_model = glm(
  as.formula(paste("cvd ~", paste(existing_vars[existing_vars != "cvd"], collapse = " + "))),
  data = cvd_africa_2024,
  family = binomial
)

imputed_case_results <- tidy(complete_case_model, conf.int = TRUE) %>%
  mutate(method = "imputed_case", analysis = "Imputed Case")

# Combine with complete case results
all_results <- bind_rows(complete_case_results, imputed_case_results)

# 3. BIAS ASSESSMENT METRICS
reference <- complete_case_results %>%
  dplyr::select(term, estimate_ref = estimate, conf.low_ref = conf.low, conf.high_ref = conf.high)

bias_metrics <- all_results %>%
  filter(method != "complete_case") %>%
  left_join(reference, by = "term") %>%
  mutate(
    bias = estimate - estimate_ref,
    relative_bias = (estimate - estimate_ref) / pmax(abs(estimate_ref), 0.001) * 100,
    ci_overlap = !(conf.high < conf.low_ref | conf.low > conf.high_ref),
    significant_change = sign(estimate) != sign(estimate_ref) & 
      abs(estimate) > 0.1 & abs(estimate_ref) > 0.1
  )

# 4. SUMMARY STATISTICS
bias_summary <- bias_metrics %>%
  group_by(method) %>%
  summarise(
    mean_absolute_bias = mean(abs(bias), na.rm = TRUE),
    median_absolute_bias = median(abs(bias), na.rm = TRUE),
    max_absolute_bias = max(abs(bias), na.rm = TRUE),
    proportion_significant_bias = mean(abs(relative_bias) > 10, na.rm = TRUE),
    ci_overlap_rate = mean(ci_overlap, na.rm = TRUE),
    sign_changes = sum(significant_change, na.rm = TRUE),
    n_variables = n()
  ) %>%
  arrange(mean_absolute_bias)

# 5. STATISTICAL TESTS
bias_test_results <- list()
#bias_test_results$anova <- aov(abs(bias) ~ method, data = bias_metrics)
bias_test_results$pairwise <- pairwise.t.test(abs(bias_metrics$bias), bias_metrics$method, 
                                              p.adjust.method = "bonferroni")

correlation_metrics <- bias_metrics %>%
  group_by(method) %>%
  summarise(
    correlation = cor(estimate, estimate_ref, use = "complete.obs"),
    rmse = sqrt(mean((estimate - estimate_ref)^2, na.rm = TRUE)),
    mae = mean(abs(estimate - estimate_ref), na.rm = TRUE)
  )

# 6. VISUALIZATIONS
# Forest plot
forest_plot <- all_results %>%
  filter(term != "(Intercept)") %>%
  ggplot(aes(x = estimate, y = term, color = method)) +
  geom_point(position = position_dodge(width = 0.6), size = 2) +
  geom_errorbarh(aes(xmin = conf.low, xmax = conf.high), 
                 position = position_dodge(width = 0.6), height = 0.2, size = 0.7) +
  geom_vline(xintercept = 0, linetype = "dashed", alpha = 0.5) +
  labs(title = "Sensitivity Analysis: Comparison of Imputation Methods",
       subtitle = "Point estimates and 95% confidence intervals",
       x = "Coefficient Estimate", y = "Predictor Variables", color = "Method") +
  theme_minimal() +
  theme(legend.position = "bottom", axis.text.y = element_text(size = 8))

# Bias distribution
bias_distribution <- bias_metrics %>%
  ggplot(aes(x = abs(bias), fill = method)) +
  geom_density(alpha = 0.5) +
  labs(title = "Distribution of Absolute Bias", x = "Absolute Bias", y = "Density") +
  facet_wrap(~method, ncol = 2) +
  theme_minimal()

# Method comparison plot
method_comparison <- bias_summary %>%
  ggplot(aes(x = reorder(method, mean_absolute_bias), y = mean_absolute_bias, fill = method)) +
  geom_bar(stat = "identity", alpha = 0.8) +
  geom_text(aes(label = round(mean_absolute_bias, 3)), hjust = -0.1, size = 3.5) +
  coord_flip() +
  labs(title = "Mean Absolute Bias by Imputation Method", 
       x = "Imputation Method", y = "Mean Absolute Bias") +
  theme_minimal() +
  theme(legend.position = "none")

# 7. INTERPRETATION AND REPORTING
cat("SENSITIVITY ANALYSIS RESULTS\n")
cat("=============================\n\n")

cat("Complete cases: ", nrow(complete_case_data), "/", nrow(cvd_africa_2024_clean), 
    " (", round(nrow(complete_case_data)/nrow(cvd_africa_2024_clean)*100, "%)\n\n", sep = "")
    
    cat("BIAS ASSESSMENT SUMMARY:\n")
    dplyr::as_tibble(bias_summary) %>% print(n = Inf)
    cat("\n")
    
    cat("CORRELATION AND ERROR METRICS:\n")
    print(correlation_metrics, n = Inf)
    cat("\n")
    
    cat("STATISTICAL TESTS:\n")
    #cat("ANOVA p-value:", summary(bias_test_results$anova)[[1]]$'Pr(>F)'[1], "\n")
    cat("Pairwise comparisons:\n")
    print(bias_test_results$pairwise$p.value)
    cat("\n")
    
    # Bias interpretation
    problematic_methods <- bias_summary %>% filter(mean_absolute_bias > 0.1 | proportion_significant_bias > 0.3)
    best_method <- bias_summary %>% slice(which.min(mean_absolute_bias))
    
    cat("INTERPRETATION:\n")
    if (nrow(problematic_methods) > 0) {
      cat("⚠️  POTENTIAL BIAS DETECTED in methods:", paste(problematic_methods$method, collapse = ", "), "\n")
    } else {
      cat("✅ No substantial bias detected across methods\n")
    }
    cat("🏆 Best performing method:", best_method$method, "(MAE:", round(best_method$mean_absolute_bias, 4), ")\n")
    
    if (any(correlation_metrics$correlation < 0.7)) {
      cat("⚠️  Low correlation with complete case analysis for some methods\n")
    } else {
      cat("✅ Good correlation with complete case analysis\n")
    }
    
    # 8. SAVE RESULTS
    write.csv(all_results, "sensitivity_analysis_full_results.csv", row.names = FALSE)
    write.csv(bias_metrics, "bias_assessment_detailed.csv", row.names = FALSE)
    write.csv(bias_summary, "bias_summary_statistics.csv", row.names = FALSE)
    write.csv(correlation_metrics, "correlation_metrics.csv", row.names = FALSE)
    
    
    
    
    # Load required libraries
    library(ggplot2)
    library(dplyr)
    library(ggrepel)
    library(patchwork)
    
    # Prepare data for sensitivity comparison
    sensitivity_comparison <- all_results %>%
      filter(term != "(Intercept)") %>%
      select(term, estimate, analysis) %>%
      pivot_wider(names_from = analysis, values_from = estimate) %>%
      mutate(
        diff = `Imputed Case` - `Complete Case`,
        abs_diff = abs(diff),
        variable_type = case_when(
          grepl("age_cat", term) ~ "Age",
          grepl("gender", term) ~ "Sex",
          grepl("residence", term) ~ "Residence",
          grepl("bmi_cat", term) ~ "BMI",
          grepl("edlev", term) ~ "Education",
          grepl("bp", term) ~ "Hypertension",
          grepl("highsalt", term) ~ "Salt Intake",
          grepl("lfi", term) ~ "Fruit Intake",
          grepl("pa", term) ~ "Physical Activity",
          grepl("smokehis", term) ~ "Smoking",
          grepl("country", term) ~ "Country",
          TRUE ~ "Other"
        )
      )
    
    # 1. CORRELATION PLOT
    correlation_plot <- ggplot(sensitivity_comparison, 
                               aes(x = `Complete Case`, y = `Imputed Case`, color = variable_type)) +
      geom_abline(intercept = 0, slope = 1, linetype = "dashed", color = "gray50") +
      geom_point(size = 3, alpha = 0.8) +
      geom_smooth(method = "lm", se = FALSE, color = "black", linetype = "dotted") +
      geom_text_repel(aes(label = ifelse(abs_diff > 0.15, term, "")), 
                      size = 3, max.overlaps = 20) +
      scale_color_brewer(palette = "Set3") +
      labs(title = "Sensitivity Analysis: Complete Case vs Imputed Case",
           subtitle = "Correlation of Coefficient Estimates",
           x = "Complete Case Estimates",
           y = "Imputed Case Estimates",
           color = "Variable Type",
           caption = "Diagonal line represents perfect agreement") +
      theme_minimal() +
      theme(legend.position = "bottom",
            plot.title = element_text(face = "bold", size = 16, hjust = 0.5),
            plot.subtitle = element_text(size = 12, hjust = 0.5))
    
    # 2. BLAND-ALTMAN PLOT (Difference vs Average)
    bland_altman_data <- sensitivity_comparison %>%
      mutate(
        average = (`Complete Case` + `Imputed Case`) / 2,
        difference = `Imputed Case` - `Complete Case`
      )
    
    bland_altman_plot <- ggplot(bland_altman_data, aes(x = average, y = difference, color = variable_type)) +
      geom_hline(yintercept = 0, linetype = "dashed", color = "red") +
      geom_hline(yintercept = mean(bland_altman_data$difference), linetype = "dashed", color = "blue") +
      geom_hline(yintercept = mean(bland_altman_data$difference) + 1.96*sd(bland_altman_data$difference), 
                 linetype = "dotted", color = "gray50") +
      geom_hline(yintercept = mean(bland_altman_data$difference) - 1.96*sd(bland_altman_data$difference), 
                 linetype = "dotted", color = "gray50") +
      geom_point(size = 3, alpha = 0.8) +
      geom_text_repel(aes(label = ifelse(abs(difference) > 0.15, term, "")), 
                      size = 3, max.overlaps = 15) +
      scale_color_brewer(palette = "Set3") +
      labs(title = "Bland-Altman Plot: Agreement Between Methods",
           subtitle = "Difference vs Average of Estimates",
           x = "Average of Complete Case and Imputed Case Estimates",
           y = "Difference (Imputed - Complete Case)",
           color = "Variable Type",
           caption = "Red line: Perfect agreement\nBlue line: Mean difference\nDotted lines: 95% limits of agreement") +
      theme_minimal() +
      theme(legend.position = "bottom")
    
    # 3. DIFFERENCE DISTRIBUTION PLOT 
    difference_plot <- ggplot(sensitivity_comparison, aes(x = diff, fill = variable_type)) +
      geom_histogram(alpha = 0.8, bins = 20, color = "white") +
      geom_vline(xintercept = 0, linetype = "dashed", color = "red") +
      geom_vline(xintercept = mean(sensitivity_comparison$diff), linetype = "dashed", color = "blue") +
      scale_fill_brewer(palette = "Set3") +
      labs(title = "Distribution of Differences",
           subtitle = "Imputed Case - Complete Case Estimates",
           x = "Difference in Coefficient Estimates",
           y = "Frequency",
           fill = "Variable Type") +
      theme_minimal() +
      theme(legend.position = "bottom")
    
    # 4. SUMMARY STATISTICS
    mean_diff <- mean(sensitivity_comparison$diff, na.rm = TRUE)
    sd_diff <- sd(sensitivity_comparison$diff, na.rm = TRUE)
    correlation <- cor(sensitivity_comparison$`Complete Case`, sensitivity_comparison$`Imputed Case`, 
                       use = "complete.obs")
    rmse <- sqrt(mean(sensitivity_comparison$diff^2, na.rm = TRUE))
    
    # Create summary annotation
    summary_text <- paste(
      "Mean difference:", round(mean_diff, 4), "\n",
      "SD of differences:", round(sd_diff, 4), "\n",
      "Correlation:", round(correlation, 4), "\n",
      "RMSE:", round(rmse, 4), "\n",
      "95% Limits of Agreement: [", 
      round(mean_diff - 1.96*sd_diff, 4), ", ", 
      round(mean_diff + 1.96*sd_diff, 4), "]"
    )
    
    # 5. COMBINE PLOTS
    combined_plot <- (correlation_plot + bland_altman_plot) / 
      (difference_plot + plot_annotation(caption = summary_text, theme = theme_minimal())) +
      plot_layout(guides = "collect") &
      theme(legend.position = "bottom")
    
    # Display combined plot
    print(combined_plot)
    
    # Save high-quality version
    ggsave("sensitivity_comparison_combined.png", combined_plot, 
           width = 16, height = 12, dpi = 300, bg = "white")
    
    # 6. QUANTITATIVE ASSESSMENT
    cat("SENSITIVITY ANALYSIS RESULTS\n")
    cat("=============================\n\n")
    cat("Correlation between methods:", round(correlation, 4), "\n")
    cat("Mean difference (Imputed - Complete):", round(mean_diff, 4), "\n")
    cat("Standard deviation of differences:", round(sd_diff, 4), "\n")
    cat("Root Mean Square Error:", round(rmse, 4), "\n")
    cat("95% Limits of Agreement: [", 
        round(mean_diff - 1.96*sd_diff, 4), ", ", 
        round(mean_diff + 1.96*sd_diff, 4), "]\n\n")
    
    # Count variables with substantial differences
    substantial_diff <- sum(abs(sensitivity_comparison$diff) > 0.1, na.rm = TRUE)
    cat("Variables with difference > 0.1:", substantial_diff, "/", nrow(sensitivity_comparison), 
        "(", round(substantial_diff/nrow(sensitivity_comparison)*100, 1), "%)\n")
    
    # Variables with largest differences
    largest_diffs <- sensitivity_comparison %>%
      arrange(desc(abs_diff)) %>%
      head(10) %>%
      select(term, `Complete Case`, `Imputed Case`, diff, abs_diff, variable_type)
    
    cat("\nTop 10 variables with largest differences:\n")
    print(largest_diffs)
    
    # 7. AGREEMENT ASSESSMENT
    agreement_stats <- sensitivity_comparison %>%
      summarise(
        perfect_agreement = sum(abs(diff) < 0.01),
        good_agreement = sum(abs(diff) < 0.05),
        moderate_agreement = sum(abs(diff) < 0.1),
        poor_agreement = sum(abs(diff) >= 0.1),
        total = n()
      )
    
    cat("\nAgreement Assessment:\n")
    cat("Perfect agreement (diff < 0.01):", agreement_stats$perfect_agreement, 
        "(", round(agreement_stats$perfect_agreement/agreement_stats$total*100, 1), "%)\n")
    cat("Good agreement (diff < 0.05):", agreement_stats$good_agreement, 
        "(", round(agreement_stats$good_agreement/agreement_stats$total*100, 1), "%)\n")
    cat("Moderate agreement (diff < 0.1):", agreement_stats$moderate_agreement, 
        "(", round(agreement_stats$moderate_agreement/agreement_stats$total*100, 1), "%)\n")
    cat("Poor agreement (diff ≥ 0.1):", agreement_stats$poor_agreement, 
        "(", round(agreement_stats$poor_agreement/agreement_stats$total*100, 1), "%)\n")
    
```
