## Supplementary material for "The Prevalence, Prevention, and Treatment of Cardiovascular Diseases in Twelve African Countries (2014-2019): An Analysis of the World Health Organisation STEPwise Approach to Chronic Disease Risk Factor Surveillance": Codes: Functions_rmph.R

```
# Functions for Introduction to Regression Methods for Public Health Using R (RMPH)
# Ramzi W. Nahhas, PhD
#

# July 13, 2022

# These functions are provided with no express or implied warranty and
# are licensed under a Creative Commons Attribution-NonCommercial-NoDerivatives 4.0 International License
# (https://creativecommons.org/licenses/by-nc-nd/4.0/).

# Linear regression ####

contplot <- function(PREDICTOR, OUTCOME, DAT, COL = "gray", ...) {
  # The contplot() function is provided with no express or implied warranty.
  
  # Plotting OUTCOME vs. a continuous predictor
  # OUTCOME is a string indicating the outcome
  # PREDICTOR is a string indicating the predictor
  # DAT is the dataframe containing this data
  # ... allows you to pass additional arguments to plot below
  # Assign to y and x
  y <- DAT[[OUTCOME]]
  x <- DAT[[PREDICTOR]]
  # Scatterplot with regression line
  plot(x, y, las = 1, pch = 20, col=COL,
       font.lab = 2, font.axis = 2, ...)
  abline(lm(y ~ x), col = "red", lwd = 2)
}

catplot <- function(PREDICTOR, OUTCOME, DAT, COL = "gray", ...) {
  # The catplot() function is provided with no express or implied warranty.
  
  # Plotting OUTCOME vs. a categorical predictor
  # OUTCOME is a string indicating the outcome
  # PREDICTOR is a string indicating the predictor
  # DAT is the dataframe containing this data
  # ... allows you to pass additional arguments to plot below
  # Assign to y and x
  y <- DAT[[OUTCOME]]
  x <- DAT[[PREDICTOR]]
  # Plot
  plot.default(x, y, las = 1, pch = 20, col=COL,
               font.lab = 2, font.axis = 2, xaxt = "n", ...)
  LEVELS <- levels(x)
  NX     <- length(LEVELS)
  axis(1, at = 1:NX, labels = LEVELS, font = 2)
  # Compute mean y at each level of x
  MEANS <- tapply(y, x, mean, na.rm = T)
  # Add means to the plot
  points(1:NX, MEANS, col = "black", pch = 20, cex = 3)
  lines( 1:NX, MEANS, col = "red")  
}

check_normality <- function(fit, sample.size=T, ylim = NULL, ...) {
  # The check_normality() function is provided with no express or implied warranty.
  
  # If ylim is not supplied, the code will automatically figure out what 
  # it should be

  # Unstandardized residuals
  RESID   <- fit$residuals

  if(sample.size) {
    # Sample size vs. # of predictors
    print(paste("The sample size is",
                length(RESID),
                "and there are",
                length(coef(fit))-1,
                "predictors."))
    print(paste("There are",
                round(length(RESID)/(length(coef(fit))-1), 1),
                "observations per predictor."))
  }
  
  par(mfrow=c(1,2))
  # Histogram of residuals
  HIST    <- hist(RESID, plot = F)$density
  DENSITY <- density(RESID, na.rm=T)
  if (is.null(ylim)) {
    # Automatically compute ylim to include both the histogram
    # and the density curve in the plot
    ylim <- c(0, max(c(HIST, DENSITY$y)))
  }
  hist(RESID, xlab = "Residuals", probability = T, ylim=ylim, ...)
  lines(DENSITY, lwd = 2, col = "red")
  curve(dnorm(x, mean = mean(RESID, na.rm=T), sd = sd(RESID, na.rm=T)),
        lty = 2, lwd = 2, add = TRUE, col = "blue")
  
  # Normal quantile-quantile (QQ) plot
  # qqnorm(as.numeric(RESID), col="red", pch=20)
  qqnorm(rstandard(fit), col="red", pch=20)
  abline(a=0, b=1, col="blue", lty=2, lwd=2)
  par(mfrow=c(1,1))
}
# Example
# check_normality(fit.ex6.1, main="")

# Epidemiology functions ####
# odds and odds ratio (OR)
# logit and inverse logit function
# These epidemiology functions function are provided with no express or implied warranty.
{
  odds       <- function(p)      p/(1-p)
  odds.ratio <- function(p1, p2) odds(p1)/odds(p2)
  logit      <- function(p)      log(p/(1-p))
  ilogit     <- function(x)      exp(x)/(1+exp(x))
  OR2.2  <- function(a,b,c,d){
    # 2x2 table, E=exposure, D=disease
    #     | D+ |  D-
    #  --------------
    #  E+ | a  |  b
    #  --------------
    #  E- | c  |  d
    #  --------------
    (a*d)/(b*c)
  }
  # Relative risk (RR)
  RR     <- function(p1, p2) p1/p2
  RR2.2  <- function(a,b,c,d){
    # 2x2 table, E=exposure, D=disease
    #     | D+ |  D-
    #  --------------
    #  E+ | a  |  b
    #  --------------
    #  E- | c  |  d
    #  --------------
    # RR = risk of D+ among E+ / risk of D+ among E-
    (a/(a+b)) / (c/(c+d))
  }
}

# Logistic regression ####

calibration.plot <- function(fit, g = 10, show.p = T,
                             show.bins = F, show.points = F, silent = T, drop.leading0 = T,
                             zoom = F, zoom.x = zoom, zoom.y = zoom, smooth.df = 5,
                             TITLE = "Calibration Plot") {
  # The calibration.plot() function is provided with no express or implied warranty.
  
  # Plot observed proportions vs. predicted probabilities, binned by predicted probability groups
  # Visualization of Hosmer-Lemeshow test
  
  # Takes same arguments as ResourceSelection::hoslem.test
  # fit = a glm() fit from a logistic regression
  # g = number of bins to use to calculate quantiles
  # Additional arguments
  # show.p = T to show the HL p-value on the plot
  # show.bins = T to add vertical lines at the bin boundaries
  # show.points = T to plot the jittered observed and predicted values
  # silent = F to return the Hosmer-Lemeshow (HL) test
  # drop.leading0 = T to drop the 0 before the decimal in the HL p-value
  # zoom = T to zoom in on the triangles
  # Any of zoom, zoom.x, and zoom.y can be c(a,b) to use a custom range

  # x = a numeric vector of observations, binary (0/1)
  x <- fit$y
  # y = expected values
  y <- fit$fitted.values
  
  # Bin the expected values into g equal size groups
  # using quantiles
  # g + 1 breaks leads to g bins
  BREAKS = quantile(y, probs = seq(0, 1, length = g + 1))
  Y.CUT   = cut(y, breaks = BREAKS, include.lowest = T, ordered_result = T)
  # table(Y.CUT, exclude = NULL)

  # Average estimated proportions in each bin
  P.EXP = tapply(y, list(Y.CUT), mean)
  # Proportion of observed outcome = 1 values in each bin
  P.OBS = tapply(x, list(Y.CUT), mean)
  # Number of observed outcome = 1 values in each bin
  X = as.numeric(tapply(x, list(Y.CUT), sum))
  # Size of each bin
  N  = as.numeric(tapply(x, list(Y.CUT), length))
  # CI for P.OBS
  CI <- vector("list", length(N))
  for(i in 1:length(N)) CI[[i]] <- as.numeric(binom.test(X[i], N[i])$conf.int)
  
  # Hosmer-Lemeshow test
  HL <- ResourceSelection::hoslem.test(x, y, g)
  P.FORMATTED <- format(HL$p.value, digits=3, nsmall=3)
  if(drop.leading0) P.FORMATTED <- substr(P.FORMATTED, 2, nchar(P.FORMATTED))
  P  <- case_when(HL$p.value < .001 ~ "p < .001",
                  TRUE              ~ paste("p =", P.FORMATTED))
  
  # # Checking that numbers being plotted match the HL test
  # all((HL$observed[,2] - X) == 0)
  # all((apply(HL$observed, 1, sum) - N) == 0)
  # all(round(HL$expected[,2] - P.EXP*N, 5) == 0)
  
  # Calibration plot
  XLIM <- YLIM <- c(0, 1)
  OFFSET <- 0.10*range(P.EXP)
  if(is.logical(zoom.x)) {
    if(zoom.x) {
      XLIM <- c(max(0, min(P.EXP               ) - OFFSET[1]),
                min(1, max(P.EXP               ) + OFFSET[2]))
    }
  } else if(length(zoom.x) == 2 & is.numeric(zoom.x)) {
    XLIM <- zoom.x
  }
  if(is.logical(zoom.y)) {
    if(zoom.y) {
      YLIM <- c(max(0, min(c(P.OBS, unlist(CI))) - OFFSET[1]),
                min(1, max(c(P.OBS, unlist(CI))) + OFFSET[2]))
    }
  } else if(length(zoom.y) == 2 & is.numeric(zoom.y)) {
    YLIM <- zoom.y
  }
  
  plot(0, 0, col = "white", xlim = XLIM, ylim = YLIM,
       xlab = "Average predicted probability", ylab = "Observed proportion",
       main = TITLE)

  if(show.points) points(jitter(y, 0.05), jitter(x, 0.05), pch = 20, cex = 0.75, col = "gray")
  
  abline(a = 0, b = 1, lty = 2, lwd = 2, col = "darkgray")
  points(P.EXP, P.OBS, pch = 2)
  for(i in 1:length(P.EXP)){
    lines(rep(P.EXP[i], 2), CI[i][[1]])
  }
  if(show.bins) abline(v = BREAKS, lty = 5, col = "darkgray")
  SUB = !is.na(P.EXP) & !is.na(P.OBS)
  lines(smooth.spline(P.EXP[SUB], P.OBS[SUB], df = smooth.df))
  op = par(usr=c(0,1,0,1))
  legend(0, 1, c("Perfect", "Observed"),
         lwd = c(2, 1), lty = c(2, 1), col = c("darkgray", "black"),
         bty = "n", seg.len = 4)
  if (show.p) text(1, 0.1, P, pos = 2)
  par(op)
  if(!silent) return(HL)
}

# Complex survey analysis ####

# # NOT NEEDED... The error.df option in car::Anova.svyglm allows changing the DF
# Anova_design_df <- function(FIT) {
#   # The Anova_design_df() function is provided with no express or implied warranty.
#   
#   ANOVA <- car::Anova(FIT, type = 3, test.statistic = "F")
#   ANOVA$`Pr(>F)` <- pf(ANOVA$F, ANOVA$Df, degf(FIT$survey.design),
#                        lower.tail = F)
#   rownames(ANOVA)[length(rownames(ANOVA))] <- "Design DF"
#   ANOVA["Design DF", "Df"] <- degf(FIT$survey.design)
#   return(ANOVA)
# }

svycontrast_design_df <- function(FIT, VECTOR, LEVEL=0.95, EXP=F, DESIGN.DF=T) {
  # The svycontrast_design_df() function is provided with no express or implied warranty.
  # Specify DESIGN.DF=F to use R's default DF

  CONTRAST <- svycontrast(FIT, VECTOR)
  
  # Use coef() and vcov() to extract elements from the contrast
  EST <- as.numeric(coef(CONTRAST))
  SE  <- as.numeric(sqrt(vcov(CONTRAST)))
  
  # For svycoxph, the resid df is in degf.resid
  # Create df.residual so the code below will work with a svycoxph object
  if("svycoxph" %in% class(FIT)){
    FIT$df.residual <- FIT$degf.resid
  }

  # Compute the CI
  if(DESIGN.DF) {
    CI  <- EST + c(-1,1)*qt(1 - (1 - LEVEL)/2, degf(FIT$survey.design))*SE
  } else { # Default R DF
    CI  <- EST + c(-1,1)*qt(1 - (1 - LEVEL)/2, FIT$df.residual)*SE
  }
  
  # If you want the est and ci for exp(contrast)
  if(EXP) {
    CI  <- exp(CI)
    EST <- exp(EST)
  }
  return(data.frame(est=EST, lower=CI[1], upper=CI[2]))
}

# Multiple imputation ####

round.summary <- function(FIT.MI, conf.int=T, digits=4, DATA.FRAME=T, ...) {
  # The round.summary() function is provided with no express or implied warranty.
  
  # Function to allow rounding of summary(pool(FIT.MI))
  require(tidyverse)
  # Convert to a data.frame
  DF <- as.data.frame(summary(pool(FIT.MI), conf.int=conf.int, ...))
  # Move term from column to rownames
  rownames(DF) <- DF$term
  DF <- DF %>% 
    select(-term)
  # Convert to matrix, round
  DF <- round(as.matrix(DF), digits)
  # Convert back to a data.frame (if DATA.FRAME=T)
  if (DATA.FRAME) DF <- as.data.frame(DF)
  return(DF)
}

# NO LONGER USED - Outdated. See von Hippel 2020 for better method
# incorporated in Section 9.9 in the text
# nimpute <- function(DAT, method = "any") {
#   # The nimpute() function is provided with no express or implied warranty.
#   
#   # METHOD = "avg" to compute based on the average (over variables) fraction of cases with missing data
#   #                among all variables, regardless of whether or not they have any missing values.
#   # METHOD = "avg_gt_0" to compute based on the average (over variables) fraction of cases with missing data
#   #                     among variables that have any missing values.
#   # METHOD = "any" to compute based on the fraction of cases with a missing value for any variable.
#   if (method == "avg") {
#     p <- mean(sapply(DAT, function(x) mean(is.na(x))))
#   } else if (method == "avg_gt_0") {
#     p <- sapply(DAT, function(x) mean(is.na(x)))
#     p <- mean(p[p > 0])
#   } else if (method == "any") {
#     p <- mean(!complete.cases(DAT))
#   }
#   return(max(5, round(100*p)))
# }

vm <- function(x) {
  # The vm() function is provided with no express or implied warranty.
  
  # Sample variance of the mean
  var(x) / length(x)
}

mi.mean.se.sd <- function(IMP, X) {
  # The mi.mean.se.sd() function is provided with no express or implied warranty.
  
  # Compute mean and standard error of the mean using multiply imputed data
  # For ONE variable at a time
  
  # IMP is a mids object created by mice()
  # X is a character value (e.g., "varname")
  
  # So I can refer to X later
  # Do NOT use include = TRUE here as these computations
  # are just on the imputed data.
  IMPDAT    <- complete(IMP, "long")
  IMPDAT$.x <- IMPDAT[[X]]

  # Compute stats within each imputation
  MEAN     <- tapply(IMPDAT$.x, IMPDAT$.imp, mean)
  VAR.MEAN <- tapply(IMPDAT$.x, IMPDAT$.imp, vm)
  SD       <- tapply(IMPDAT$.x, IMPDAT$.imp, sd)
  
  # Use Rubin's Rules to pool
  POOLED  <- pool.scalar(Q = MEAN,
                         U = VAR.MEAN,
                         n = nrow(IMP$data))
  
  OUT <- data.frame(mean = POOLED$qbar,
                    se   = sqrt(POOLED$t),
                    sd   = mean(SD))
  rownames(OUT) <- X
  return(OUT)
}

mi.n.p <- function(IMP, X) {
  # The mi.n.p() function is provided with no express or implied warranty.
  
  # Compute frequency and proportion using multiply imputed data
  # For ONE variable at a time
  
  # IMP is a mids object created by mice()
  # X is a character value (e.g., "varname")
  
  # So I can refer to X later
  # Must use include = TRUE here or as.mids will not work 
  IMPDAT    <- complete(IMP, "long", include = TRUE)
  IMPDAT$.x <- IMPDAT[[X]]
  IMP       <- as.mids(IMPDAT)
  
  # with() automatically excludes the original data
  # and just does this on the imputed data
  TAB <- matrix(unlist(with(IMP, table(.x))$analyses),
                nrow=IMP$m, byrow = T)
  N <- apply(TAB, 2, mean)
  P <- N/sum(N)
  OUT <- data.frame(n = N, p = P)
  rownames(OUT) <- paste(X, levels(IMPDAT$.x), sep = ": ")
  return(OUT)
}

# Checking that it is just using the imputations, not .imp = 0 (the original data)
# TMP <- array(NA, dim=c(length(levels(IMPDAT[[X]])), IMP$m))
# for(i in 1:IMP$m) {
#   TMP[,i] <- table(IMPDAT[IMPDAT$.imp == i, ".x"])
# }
# apply(TMP, 1, mean)

mi.mean.se.sd.by <- function(IMP, X, BY) {
  # The mi.mean.se.sd.by() function is provided with no express or implied warranty.
  
  # Compute mean and SD using multiply imputed data by another variable
  # For ONE variable at a time
  
  # IMP is a mids object created by mice()
  # X is a character value (e.g., "varname")
  # BY is a character value for a factor variable to stratify by
  
  # So I can refer to X and BY = Z later
  # Do NOT use include = TRUE here as these computations
  # are just on the imputed data.
  IMPDAT    <- complete(IMP, "long")
  IMPDAT$.x <- IMPDAT[[X]]
  IMPDAT$.z <- IMPDAT[[BY]]

  LEVELS <- levels(IMPDAT$.z)
  
  # Get MI-based sample sizes
  N <- mi.n.p(IMP, BY)$n
  
  # Compute statistics in each imputed dataset
  for(i in 1:length(LEVELS)) {
    SUB      <- IMPDAT$.z == LEVELS[i]
    MEAN     <- tapply(IMPDAT$.x[SUB], IMPDAT$.imp[SUB], mean)
    VAR.MEAN <- tapply(IMPDAT$.x[SUB], IMPDAT$.imp[SUB], vm)
    SD       <- tapply(IMPDAT$.x[SUB], IMPDAT$.imp[SUB], sd)
    
    # Use Rubin's Rules to pool
    POOLED    <- pool.scalar(Q = MEAN,
                             U = VAR.MEAN,
                             n = N[i])
    if (i == 1) {
      MEAN.SE.SD <- c(            POOLED$qbar, sqrt(POOLED$t), mean(SD))
    } else {
      MEAN.SE.SD <- c(MEAN.SE.SD, POOLED$qbar, sqrt(POOLED$t), mean(SD))
    }
    rm(SUB, MEAN, VAR.MEAN, SD, POOLED)
  }
  
  OUT <- data.frame(matrix(MEAN.SE.SD, nrow=1))
  names(OUT) <- c(rbind(paste("mean", 1:length(LEVELS), sep="."),
                        paste("se",   1:length(LEVELS), sep="."),
                        paste("sd",   1:length(LEVELS), sep=".")))
  rownames(OUT) <- X
  return(OUT)
}

mi.n.p.by <- function(IMP, X, BY) {
  # The mi.n.p.by() function is provided with no express or implied warranty.
  
  # Compute frequency and proportion using multiply imputed data by another variable
  # For ONE variable at a time
  
  # IMP is a mids object created by mice()
  # X is a character value (e.g., "varname")
  # BY is a character value for a factor variable to stratify by
  
  # So I can refer to X and BY = Z later
  # Must use include = TRUE here or as.mids will not work 
  IMPDAT    <- complete(IMP, "long", include = TRUE)
  IMPDAT$.x <- IMPDAT[[X]]
  IMPDAT$.z <- IMPDAT[[BY]]
  IMP       <- as.mids(IMPDAT)
  
  LEVELS <- levels(IMPDAT$.z)
  
  for(i in 1:length(LEVELS)) {
    # with() automatically excludes the original data
    # and just does this on the imputed data
    TAB <- matrix(unlist(with(IMP, table(.x[.z == levels(.z)[i]]))$analyses),
                  nrow=IMP$m, byrow = T)
    N <- apply(TAB, 2, mean)
    P <- N/sum(N)
    if (i == 1) {
      N.P <- cbind(N,P)
    } else {
      N.P <- cbind(N.P, N, P) 
    }
  }
  
  OUT <- data.frame(N.P)
  names(OUT) <- c(rbind(paste("n", 1:length(LEVELS), sep="."),
                        paste("p", 1:length(LEVELS), sep=".")))
  rownames(OUT) <- paste(X, levels(IMPDAT$.x), sep = ": ")
  return(OUT)
}
```
