## Supplementary material for "The Prevalence, Prevention, and Treatment of Cardiovascular Diseases in Twelve African Countries (2014-2019): An Analysis of the World Health Organisation STEPwise Approach to Chronic Disease Risk Factor Surveillance": Tables and Figures

**Figure 1: Flow diagram of individuals included and excluded in the analysis: 2014-2019**
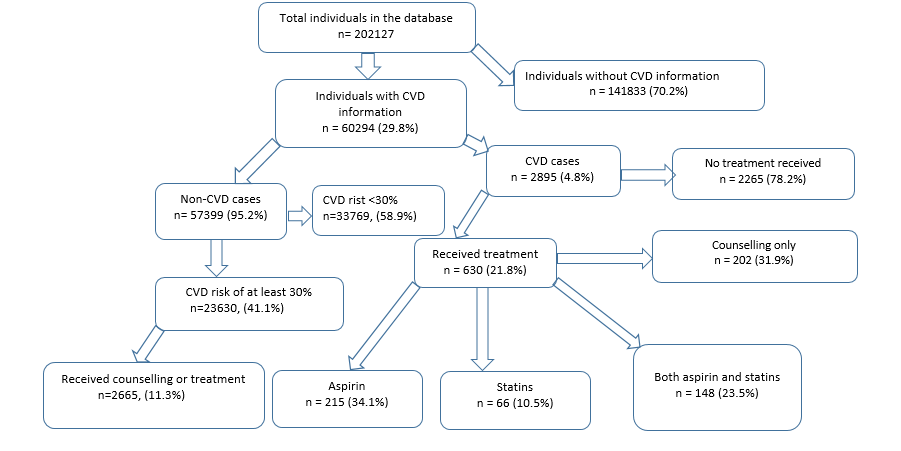

**CVD= cardiovascular diseases**

**Figure 2: Cardiovascular disease care cascade in twelve African countries, 2014–2019**

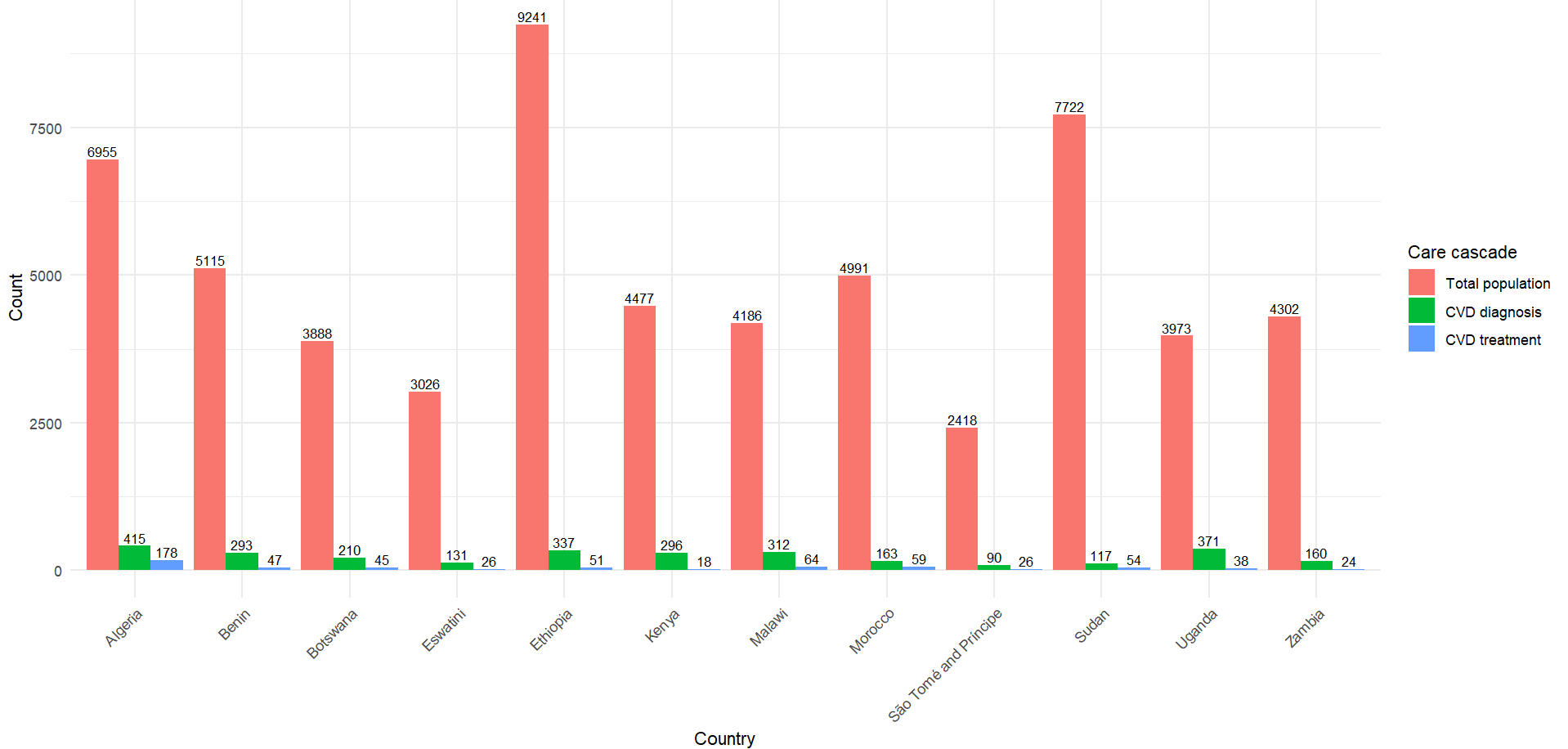

**CVD= Cardiovascular diseases**

**Figure 3: Determinants of cardiovascular disease prevalence in twelve African countries, 2014–2019**

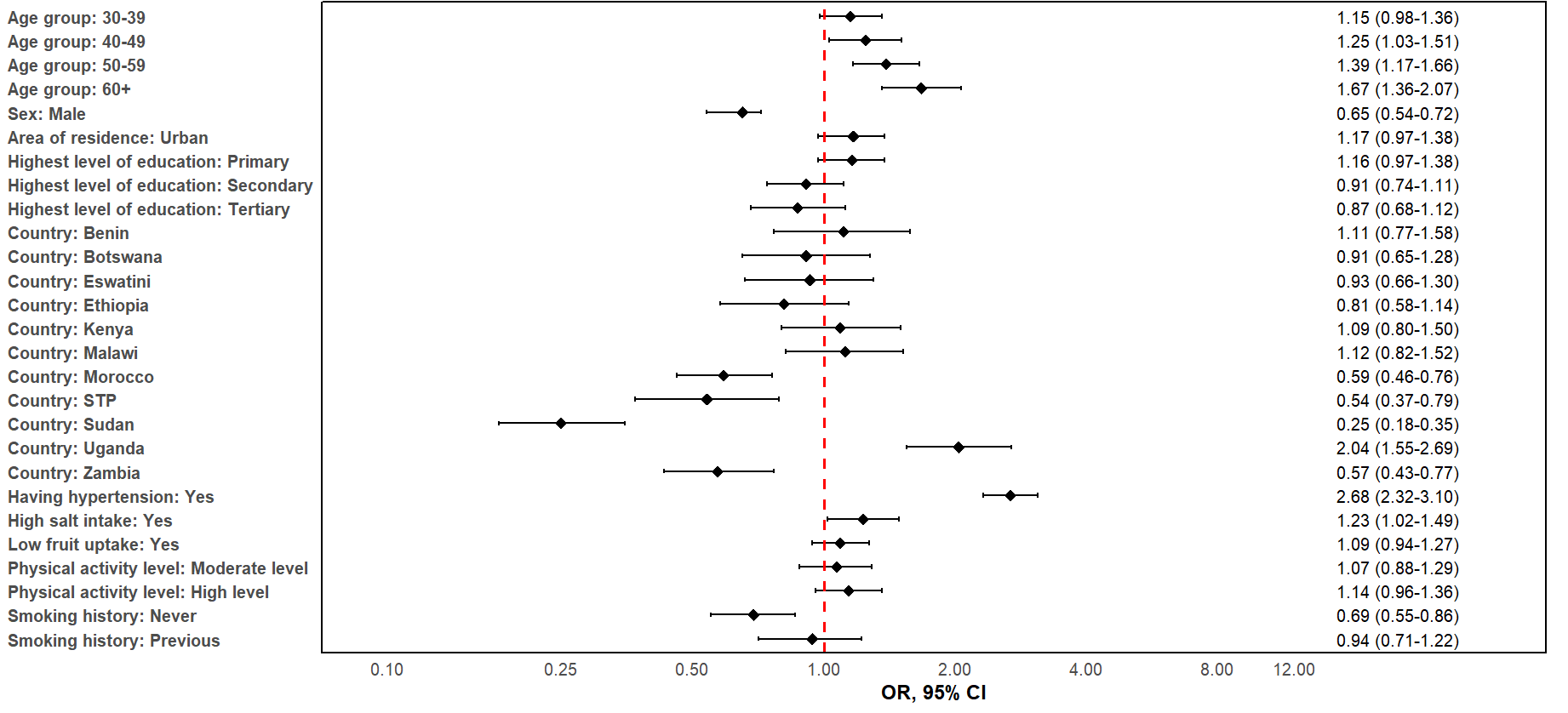

OR= Odds ratio, STP= Sao Tome and Principe

**Figure 4: Factors linked to uptake of preventive cardiovascular disease treatment in twelve African countries, 2014–2019**

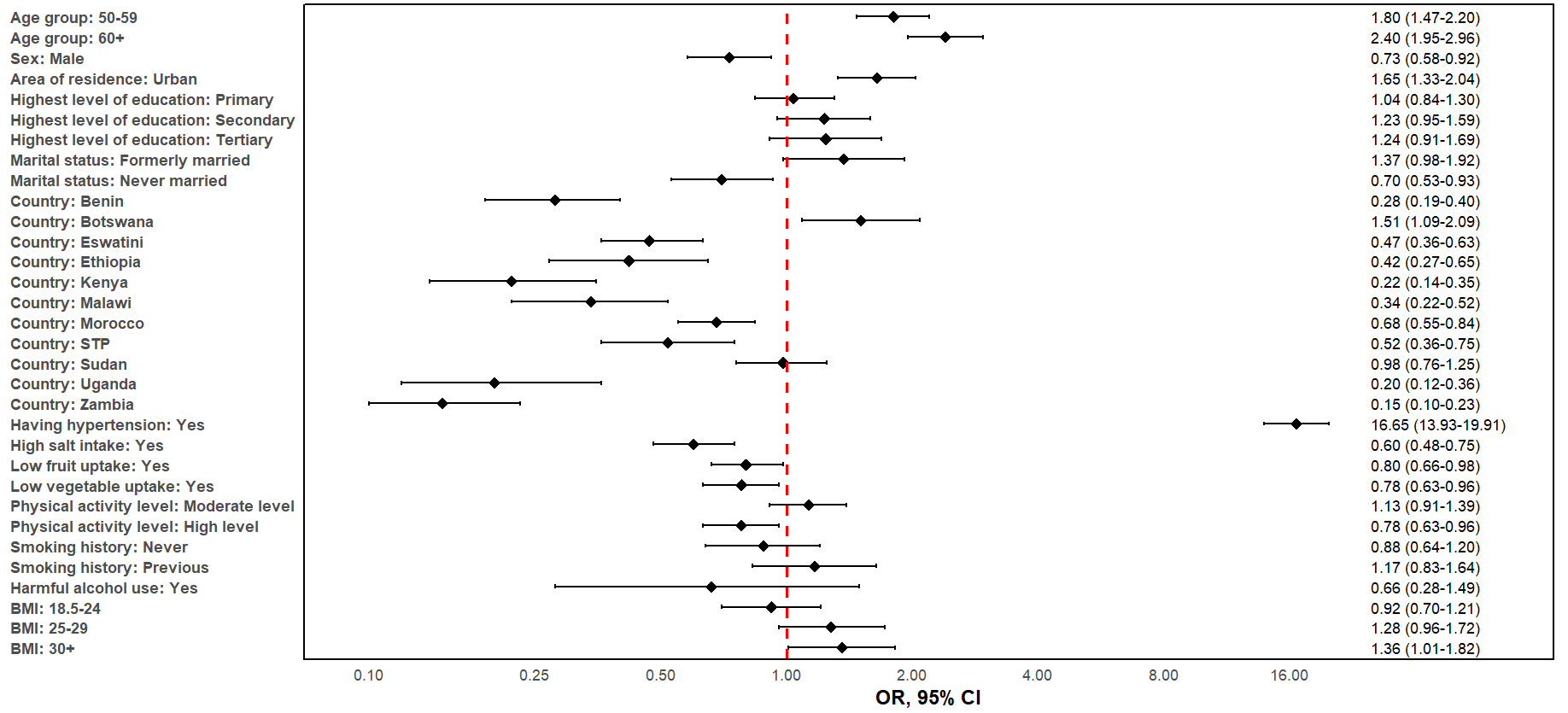

OR= Odds ratio, STP= Sao Tome and Principe

**Figure 5: Factors linked to treatment uptake amongst those diagnosed with cardiovascular disease in twelve African countries, 2014–2019**

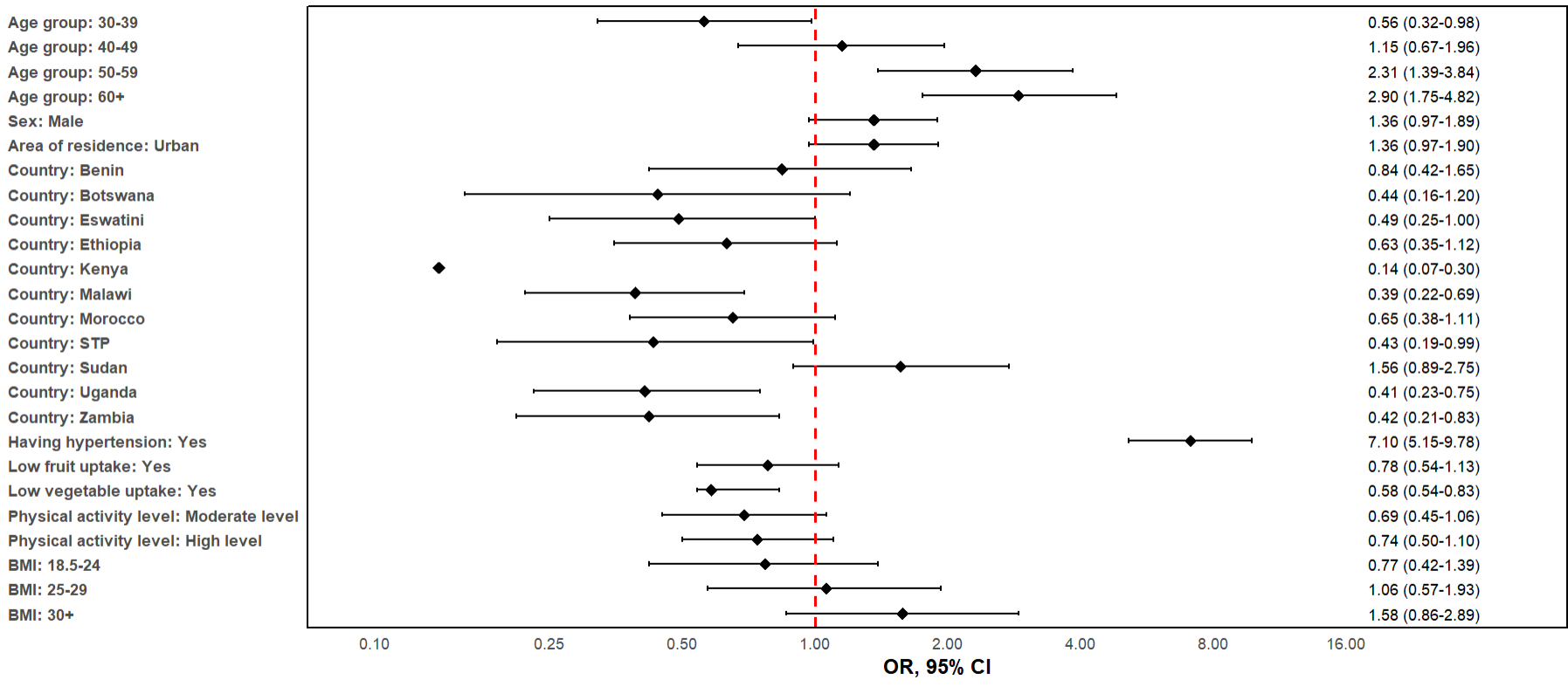

OR= Odds ratio, STP= Sao Tome and Principe

**Box 2: Proportion of missing data across variables used to assess cardiovascular disease prevalence in twelve African countries, 2014–2019**

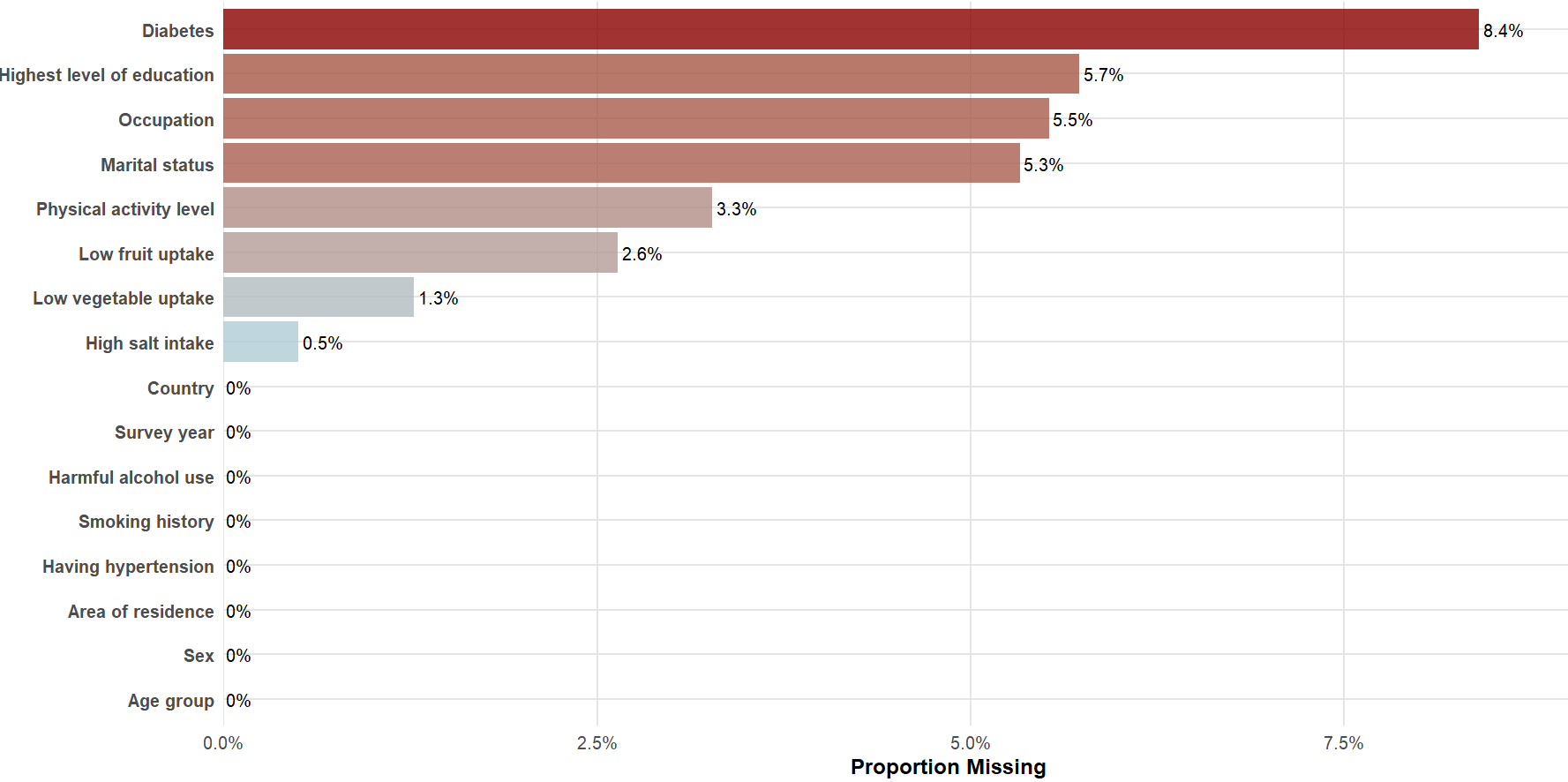

**Box 3: The Cardiovascular disease (CVD) care pathway from WHO STEPWise Surveys conducted across African countries (2014-2019)**

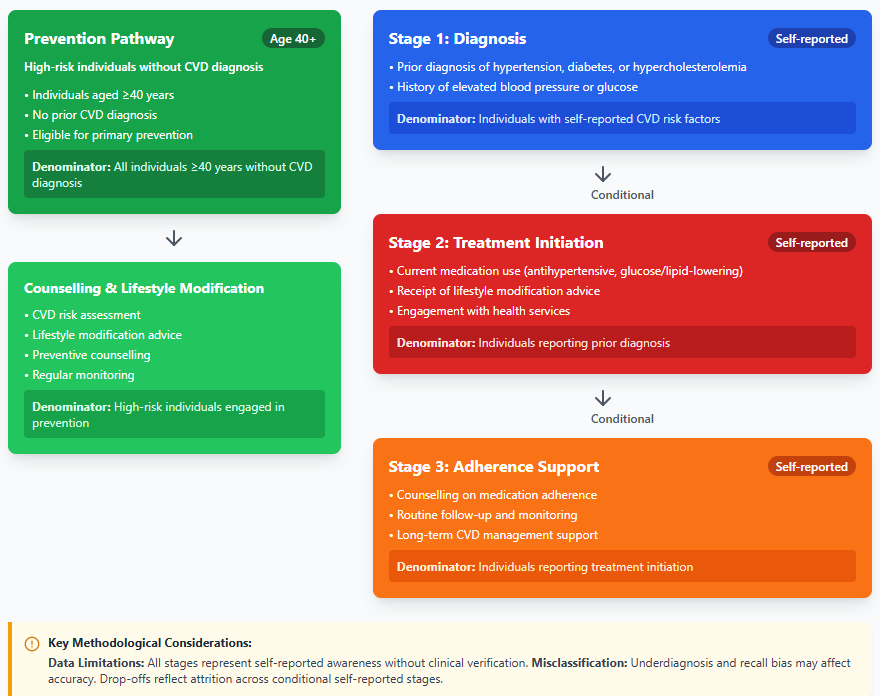

CVD= cardiovascular diseases

**Box 4: Sensitivity analysis of determinant coefficients for cardiovascular disease prevalence, comparing imputation and complete case analysis in twelve African countries, 2014–2019**

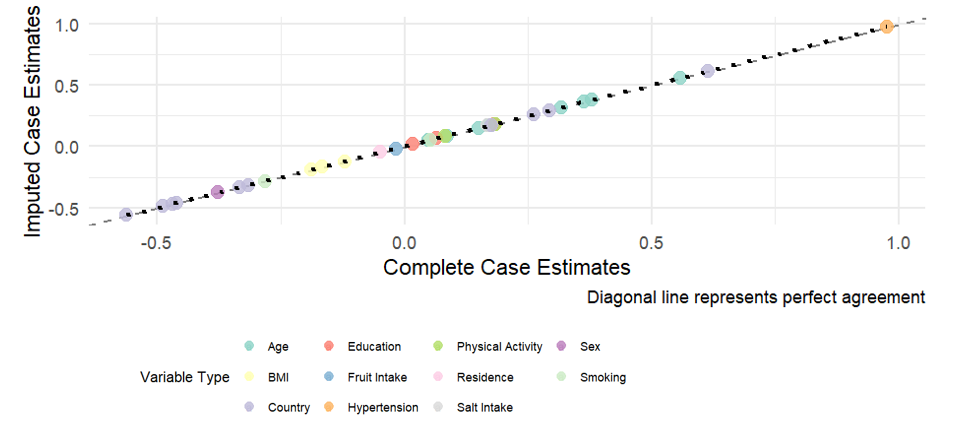
